# Safety of Radiotherapy and Radiosurgery for Optic Pathway-Hypothalamic Glioma: A Systematic Review and Meta-analysis

**DOI:** 10.64898/2026.08.17.26360611

**Authors:** Farzan Fahim, Amirmahdi Mojtahedzadeh, Farnaz Mortezazade, Parvane Tayebzadeh, Narin Biabangard, Mahan Kamali, Maryam Yaftian, Mobina Puraminaie, Hanane Sadat Hashemi, Kowsar Hariri, Borhan Rahimirad, Niloofar Sadeghi, Ava Khalili Dehkordi, Omolbanin Soleymani Pour, Fatemeh Khazaei, Alireza Zali

## Abstract

**Background:** Radiotherapy can provide durable local control for optic pathway–hypothalamic glioma (OPHG), but its use is limited by concern regarding delayed vascular, endocrine, visual, oncological, and neurological toxicities.

**Objective:** To systematically characterize and quantify the safety of radiotherapy and radiosurgery for OPHG and explore clinically relevant modifiers of treatment-related toxicity.

**Methods:** PubMed, Scopus, Web of Science, Embase, Cochrane, Google Scholar, and ClinicalTrials.gov were searched from inception through 1 June 2026. Eligible non-randomized studies reporting safety outcomes after radiotherapy or radiosurgery were included. Random-effects binomial-normal generalized linear mixed-effects models were used to pool proportions, with exact conditional models for sparse comparative analyses.

**Results:** Thirty-five studies were included, of which 31 contributed event-level data to at least one quantitative safety outcome. The pooled incidence of any treatment-related toxicity was 8.46% (95% CI, 1.37–38.01%). Vasculopathy occurred in 9.44% (95% CI, 5.22–16.49%). Secondary neoplasms occurred in 5.41% (95% CI, 2.23–12.53%), decreasing to 2.83% under a strict malignant-event definition. Incident endocrinopathy had the highest pooled estimate at 21.19% (95% CI, 4.72–59.31%) and increased with longer follow-up. Treatment-related visual toxicity was 2.26%, whereas radiation-related mortality was 0.59%. Radiation necrosis, severe toxicity, and treatment-attributed neurocognitive toxicity were sparsely reported.

**Conclusion:** Late toxicity following radiotherapy for OPHG is heterogeneous, with endocrinopathy, vasculopathy, and secondary neoplasms representing the principal quantifiable safety concerns. Treatment decisions should therefore be individualized, with prolonged vascular, endocrine, visual, and oncological surveillance and further prospective evaluation of contemporary radiation techniques.

## Introduction

Optic pathway hypothalamic gliomas (OPHGs) are rare, predominantly low-grade tumors arising along the optic nerves, chiasm, optic tracts, hypothalamus, or contiguous structures. They occur mainly in childhood, often during the first decade of life, with no consistent sex predominance across reported series, and may arise sporadically or in association with neurofibromatosis type 1 (NF1) [1, 2]. Their clinical burden reflects both tumor biology and location within highly eloquent structures. Progressive visual loss, visual-field deficits, strabismus, and ultimately blindness may occur, while hypothalamic involvement can produce endocrine dysfunction, growth abnormalities, obesity, and neurological morbidity [3, 4]. Consequently, preservation of long-term neurological, visual, and endocrine function is central to treatment selection.

Management is individualized according to age, symptoms, tumor behavior, anatomical extent, NF1 status, and prior therapy. Observation may be appropriate for selected clinically stable patients, whereas systemic therapy is frequently used in younger children to delay or avoid irradiation; surgery is generally limited by the close relationship of these tumors to the visual pathways and hypothalamus [1, 5]. Radiotherapy nevertheless remains an important option for progressive, refractory, or otherwise difficult-to-control disease, with conventional fractionated radiotherapy, conformal techniques, stereotactic radiotherapy, and radiosurgery all represented in the literature [6-8]. Its potential benefits must, however, be balanced against late toxicities including vasculopathy, endocrinopathy, visual deterioration, secondary neoplasms, neurocognitive effects, radiation necrosis, and treatment-related mortality [9-11].

Despite decades of clinical experience, the safety evidence remains fragmented. The available literature spans markedly different treatment eras, radiation techniques, follow-up durations, and toxicity definitions, while attribution of late morbidity is complicated by tumor progression, NF1-associated susceptibility, previous surgery, and systemic therapy [2, 11, 12]. Moreover, the 35 studies comprising the present evidence base consist of 27 cohort studies, six case series, and two quasi-experimental studies, with no randomized trials. Many reports are retrospective and small, and several safety domains are inconsistently or incompletely reported, limiting precise estimation of uncommon but clinically important late adverse events.

Accordingly, this systematic review and meta-analysis aimed to comprehensively characterize the safety of radiotherapy and radiosurgery for OPHG, quantify the incidence of major treatment-related adverse outcomes, evaluate clinically relevant sources of heterogeneity, and examine differences across patient and treatment subgroups where data permitted. We additionally sought to identify areas in which the existing evidence remains sparse or structurally fragmented, thereby informing treatment selection, survivorship surveillance, and priorities for future research.

## Methods

### Protocol and reporting

This systematic review and meta-analysis was conducted and reported in accordance with the **Preferred Reporting Items for Systematic Reviews and Meta-Analyses (PRISMA) 2020** statement [13]. The review protocol was registered in the International Prospective Register of Systematic Reviews (**PROSPERO; CRD420261474311**). The completed PRISMA 2020 checklist is provided in **Supplementary File 1**, and the complete search protocol and database-specific search strategies are provided in **Supplementary File 2**.

### Information sources and search strategy

A comprehensive literature search was conducted in **PubMed, Scopus, Web of Science Core Collection, Embase, and the Cochrane Library/CENTRAL** from database inception through **1 June 2026**. Supplementary searches were performed in **Google Scholar** and **ClinicalTrials.gov** to identify potentially missed records, grey literature, and relevant completed or ongoing studies. Reference lists of potentially eligible articles and relevant reviews were also examined for additional reports. No restrictions were imposed on publication date or language. When potentially eligible non-English reports were encountered, AI-assisted translation was used as necessary to facilitate eligibility assessment and data extraction, with the original report retained as the source document for interpretation.

Search strategies combined controlled vocabulary, where available, with free-text terms representing two principal concepts: **optic pathway/hypothalamic glioma** and **radiotherapy/radiosurgery**. Search terminology included variants of *optic pathway glioma*, *optic nerve glioma*, *hypothalamic glioma*, *chiasmatic glioma*, and *OPHG*, combined with terms for *radiotherapy*, *radiation therapy*, *proton therapy*, *stereotactic radiosurgery*, and *intensity-modulated radiotherapy*. Syntax was adapted to the indexing structure and search functions of each database; PubMed incorporated Medical Subject Headings, and Embase incorporated indexed terminology alongside title/abstract terms. Broader supplementary combinations were used in Google Scholar and ClinicalTrials.gov. The full strategies for all sources are reproduced in Supplementary File 2.

The complete PubMed search strategy was:

~~~
(
 (“Optic Nerve Glioma”)
 OR (“optic pathway glioma*“[Title/Abstract] ) OR (“optic nerve glioma*“[Title/Abstract]) OR (“hypothalamic glioma*“[Title/Abstract] ) OR (“chiasmatic glioma*“[Title/Abstract])
 OR (OPHG[Title/Abstract] )
)
AND
(
 (“Radiotherapy”)
 OR (“radiation therap*“[Title/Abstract] ) OR (radiotherap*[Title/Abstract] )
 OR (“proton therap*“[Title/Abstract] )
 OR (“stereotactic radiosurgery“[Title/Abstract] ) OR (IMRT[Title/Abstract] )
)
~~~

### Eligibility criteria

Eligibility was defined using a prespecified **PICOST framework** encompassing population, intervention, comparator, outcomes, study design, and time frame. Studies were eligible if they included patients of any age with optic pathway glioma, hypothalamic glioma, or optic pathway–hypothalamic glioma treated with radiotherapy and/or radiosurgery. Both sporadic and neurofibromatosis type 1 (NF1)-associated tumors were eligible. Diagnosis could be established histopathologically or according to accepted clinical and radiological criteria used by the original investigators. Studies of broader low-grade glioma or intracranial tumor populations were eligible only when data for the relevant OPHG population could be separately identified.

Eligible radiation modalities included conventional external-beam radiotherapy, fractionated radiotherapy, conformal radiotherapy, intensity-modulated radiotherapy, proton therapy, fractionated stereotactic radiotherapy, stereotactic radiosurgery, Gamma Knife or CyberKnife treatment, brachytherapy/interstitial radiotherapy, and other therapeutic radiation techniques applied to the eligible population. Radiotherapy could have been administered as primary, adjuvant, or salvage treatment or following previous surgery or systemic therapy. A comparator was not mandatory; both single-arm and comparative studies were eligible. When available, comparator data could comprise non-irradiated patients, alternative treatment strategies, different radiation techniques, or clinically relevant subgroups such as NF1-associated versus sporadic disease.

Eligible designs comprised original non-randomized cohort studies, non-randomized comparative studies, quasi-experimental studies, and case series containing at least 10 relevant patients. Case reports, case series with fewer than 10 relevant patients, cross-sectional and case-control studies, conference abstracts without sufficient original data, reviews, meta-analyses, editorials, commentaries, book chapters, animal or laboratory studies, and other non-original publications were excluded. Studies were additionally excluded when the eligible OPHG population or RT/SRS-treated subgroup could not be separately extracted, when no eligible radiation intervention was evaluated, when adequate full-text information could not be obtained to establish eligibility, or when no relevant clinical safety or treatment outcome could be extracted. No restrictions were imposed according to age, sex, geographic region, treatment era, or duration of follow-up. As a prespecified exception, Han et al. was retained despite including fewer than 10 patients because of its methodological relevance and uniquely detailed reporting of treatment-related safety outcomes.

The standardized PICOST eligibility framework is provided in **Supplementary File 4**.

### Study selection

All records retrieved from the electronic searches were exported into **EndNote 2025 (Clarivate)**, where duplicate records were identified and removed before screening. Title and abstract screening was performed independently and in duplicate by **NB and HSH**. Both reviewers screened the complete set of deduplicated records using independent copies of a standardized Microsoft Excel screening workbook designed by the senior author (**FF**). The workbook recorded article title, first author, publication year, DOI, and the reason for exclusion. At this stage, exclusions were classified principally as either an ineligible study design or a clearly irrelevant population, topic, or intervention. Disagreements were resolved through discussion and, when consensus could not be reached, adjudication by **AMM**. The finalized title/abstract screening workbook is provided in **Supplementary File 3**.

Articles retained after title and abstract assessment underwent independent full-text review by **AKD and BR**, each of whom assessed the entire set of candidate reports separately. Standardized workbooks prepared by FF were used to document both inclusion and exclusion decisions. For potentially eligible studies, the PICOST workbook recorded the title, first author, publication year, DOI, country, study design, population, intervention, comparator, outcomes, and follow-up time frame. For excluded reports, a separate workbook recorded bibliographic information and a specific reason for full-text exclusion. Disagreements regarding eligibility or the applicable exclusion reason were adjudicated by **AMM**. The finalized PICOST inclusion/eligibility workbook and the full-text exclusion workbook are provided as **Supplementary Files 4 and 5**, respectively. Study selection was subsequently summarized using the PRISMA 2020 flow diagram.

### Data extraction and outcome adjudication

Data extraction was performed independently and in duplicate by **MP and OSP**, who were trained by FF before formal extraction. Both reviewers independently extracted the complete dataset from every included study using separate copies of a standardized extraction workbook developed by FF. Discrepancies or uncertainties were reconciled by comparison with the source publication and, when required, adjudicated by **AMM**.

The extraction framework captured bibliographic and methodological characteristics; study setting and recruitment period; sample size and evaluable populations; demographic characteristics; age; sex; NF1 status; diagnostic and histopathological information; tumor location and extent; baseline visual, endocrine, neurological, and vascular status; previous surgery and systemic therapy; radiation modality, indication, timing, dose, fractionation, target and treatment platform; follow-up duration and completeness; methods of toxicity ascertainment and attribution; and outcome-specific event counts and denominators. Detailed information was extracted for overall treatment-related toxicity, severe toxicity, endocrinopathy, vasculopathy, secondary neoplasms, radiation necrosis, ophthalmic and visual toxicity, neurocognitive and neurological toxicity, other late effects, and mortality. Comparative information, including RT versus non-RT and NF1 versus sporadic subgroups, was extracted when available. The workbook also contained dedicated meta-analysis-ready numerator/denominator fields and documentation explaining why an outcome was not quantitatively synthesizable. The finalized extraction dataset is provided in **Supplementary File 6**.

Safety outcomes were adjudicated at the outcome level. Whenever possible, incident adverse outcomes occurring after radiation exposure were distinguished from abnormalities already present before treatment. Events explicitly attributed to tumor progression, surgery, systemic therapy, or another cause were not classified as radiation-related events unless the original study provided adequate treatment attribution. Similarly, numbers of adverse events were not treated as numbers of affected patients when multiple events could occur within the same individual unless a unique-patient denominator could be established. Study-specific outcome definitions were retained when standardized definitions were unavailable, and clinically incompatible outcomes were not combined solely to increase the number of studies contributing to a meta-analysis. These principles were prespecified in the review protocol.

Potentially overlapping cohorts were evaluated using study center, recruitment period, author group, patient characteristics, sample size, and treatment characteristics. When publications clearly or probably represented overlapping populations, patients were not counted more than once within the same quantitative outcome. The report providing the most complete eligible cohort, clearest outcome definition, most appropriate follow-up, or most complete event-level information was preferentially retained, with alternative choices explored in sensitivity analyses where relevant.

### Risk-of-bias assessment

Risk of bias was assessed independently by **FM and PT**, following training by FF and using separate copies of a standardized appraisal workbook prepared by FF. The appropriate **Joanna Briggs Institute (JBI) critical appraisal tool** was selected according to study design: the revised JBI critical appraisal tool for cohort studies [14], the JBI Critical Appraisal Checklist for Case Series [15], and the revised JBI critical appraisal tool for quasi-experimental studies [16]. The JBI tools are designed to evaluate potential bias in study design, conduct, measurement, confounding, follow-up, and statistical analysis according to the applicable design.

Each applicable signaling item was categorized as **Yes, No, Unclear, or Not applicable**. Items judged not applicable were excluded from the denominator of the descriptive study-level assessment. For transparent graphical presentation, the proportion of applicable items judged “Yes” was used to classify studies according to a prespecified operational rule: ≥80% as low risk, 60–79% as moderate risk, and <60% as high risk. These categories were used solely as an operational study-level summary for this review and should not be interpreted as official JBI thresholds. The two reviewers independently appraised every included study, and disagreements were resolved by **AMM**. The complete item-level assessments and study-level classifications are provided in **Supplementary File 7**.

### Data synthesis and statistical analysis

A narrative synthesis was first undertaken to characterize study populations, radiation techniques, follow-up, outcome definitions, and patterns of safety reporting. Quantitative synthesis was conducted only when sufficiently compatible studies provided an interpretable event count and corresponding denominator for the same outcome. Because meaningful between-study clinical and methodological heterogeneity was anticipated, random-effects models were used for the principal quantitative analyses.

For single-arm binary safety outcomes, pooled proportions were estimated using **binomial-normal generalized linear mixed-effects models (GLMMs)** with a logit link. For study (í), the observed number of events (*X*_í_ ) was modeled as

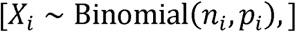

with

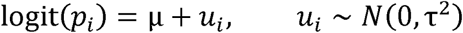

where (n_í_) denotes the eligible denominator, (µ) the pooled log-odds of the outcome, and (τ^2^) the between-study variance. The pooled proportion was obtained by inverse-logit transformation,

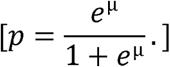

Models were fitted by maximum likelihood using `rma.glmm()` in the **metafor** package with logit-transformed proportions. This likelihood-based approach directly models binomial event data and permits incorporation of zero-event studies without imposing an arbitrary continuity correction, an important consideration for rare adverse events [17, 18].

Pooled results were reported with **95% confidence intervals (CIs)**. Between-study heterogeneity was quantified using (τ^2^) and (/^2^),with (/^2^) interpreted as the proportion of variability attributable to between-study heterogeneity rather than sampling error [19]. Where the data supported meaningful estimation, **95% prediction intervals** were additionally calculated to characterize the range in which the underlying outcome proportion of a comparable future population might be expected to lie [20].

For comparative binary data, effect estimates were expressed primarily as **odds ratios (ORs) with 95% CIs**. Sparse study-level (2 X 2) data were analyzed using exact conditional likelihood models where appropriate. Specifically, conditional GLMMs based on the exact likelihood (`model=“CM.EL“` in `metafor`) were used for rare-event comparisons; this approach conditions on the total number of events within each study and, for odds ratios, employs the corresponding non-central hypergeometric likelihood. Random-effects models were used when between-study heterogeneity could be estimated reliably; an equal-effects exact conditional formulation was used for exceptionally sparse analyses in which random-effects estimation was not supportable. Comparative findings from non-randomized studies were interpreted as associations rather than causal treatment effects.

Prespecified and clinically motivated exploratory analyses considered differences according to **NF1 status, radiation modality/technique, treatment era, follow-up duration, study population purity, toxicity ascertainment, and risk of bias**, where sufficient information was available. Random-effects meta-regression was undertaken only when the number of contributing studies and completeness of study-level covariates allowed a defensible analysis.

For a study-level moderator (*X*_í_ ), the general model was

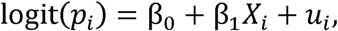

with (u_í_ = N(0, τ^2^)). Meta-regression findings were regarded as exploratory and interpreted at the study level rather than as patient-level associations.

Robustness was evaluated using a common diagnostic framework applied to every meta-analyzed safety outcome when computationally estimable. Primary binomial-normal GLMMs were refitted under outcome-specific eligibility assumptions and after sequential exclusion of each contributing study. Sensitivity restrictions addressed risk of bias, publication era, overlapping cohorts, influential studies, treatment attribution, and clinically plausible alternative outcome definitions; broad and restrictive malignant-event definitions were examined separately for secondary neoplasms. Complementary diagnostics were based on logit-transformed proportions fitted with random-effects REML models and included funnel and regression-based asymmetry visualizations, Baujat plots, Cook’s distance, externally standardized residuals, DFFITS, heterogeneity after study deletion, graphical display of study heterogeneity (GOSH) analyses, radial plots, and profile likelihoods for between-study variance. Formal regression-based asymmetry testing was undertaken only for outcomes with at least 10 contributing studies; for smaller evidence sets, the corresponding displays were descriptive. All asymmetry findings were interpreted cautiously because sparse events, true clinical heterogeneity, and differential ascertainment can produce patterns resembling publication bias. Study-level proportions were also displayed descriptively by risk of bias, radiotherapy technique, and publication era without causal or patient-level inference. Outcomes reported by very few studies or containing extremely few events were summarized with study-specific or aggregate event counts and exact binomial confidence intervals rather than subjected to unstable random-effects pooling.

All statistical analyses were performed in **R version 4.6.0**. The principal meta-analytic analyses used `metafor` (version 5.0-1), with `lme4` (2.0-1), `Matrix` (1.7-5), and `numDeriv` (2016.8-1.1) supporting mixed-model estimation and computation. Data management and workbook handling used `readxl` (1.5.0), `openxlsx` (4.2.8.1), `dplyr` (1.2.1), `tidyr` (1.3.2), `purrr` (1.2.2), and `stringr` (1.6.0). Graphical outputs were generated using `ggplot2` (4.0.3), `ggrepel` (0.9.8), `patchwork` (1.3.2), and `scales` (1.4.0).

### Exploratory evidence-topology analysis

To complement conventional outcome-specific meta-analysis, an exploratory **evidence-topology analysis** was performed to characterize how the included literature contributed across the prespecified safety domains. A binary study-by-outcome matrix was constructed indicating whether each study supplied usable evidence for any treatment-related toxicity, vasculopathy, secondary neoplasms, incident endocrinopathy, treatment-related visual toxicity, radiation-related mortality, all-cause mortality, radiation necrosis, grade ≥3 toxicity, and neurocognitive toxicity. For two outcome domains (A) and (B), shared-study connectivity was quantified using the **Jaccard similarity coefficient**,

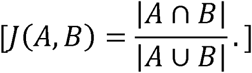

An outcome network was then constructed in which nodes represented safety domains, node size reflected the amount of contributing evidence, and edge weights reflected shared-study Jaccard similarity. Hierarchical clustering was used to explore evidence-domain groupings. Stability was evaluated using **5,000 Bayesian-bootstrap iterations**, with study weights resampled across iterations; edge stability was summarized as the probability that the weighted Jaccard coefficient exceeded 0.10, and consensus co-clustering probabilities were derived across iterations. An exploratory five-cluster solution was used to describe the resulting evidence architecture. This analysis was interpreted strictly as a map of **shared study-level evidence availability** and not as patient-level covariance or co-occurrence of toxicities, biological connectivity between outcomes, or a network meta-analysis. The conceptual framework and these interpretive boundaries were prespecified in the review protocol.

## Results

### Study selection

The systematic search identified **7,055 records** across PubMed (n=261), Scopus (n=2,609), Web of Science (n=420), Embase (n=339), Cochrane (n=5), Google Scholar (n=3,330), and ClinicalTrials.gov (n=91). After removal of **4,452 duplicate records**, 2,603 unique records underwent title and abstract screening. Of these, 2,528 were excluded because they were clearly unrelated to the population, intervention, or review topic (n=1,711) or had an ineligible study design (n=817).

Seventy-five reports proceeded to full-text assessment, all of which were successfully retrieved. Forty reports were subsequently excluded: case reports or case series with fewer than 10 relevant patients (n=11), insufficient reports (n=9), studies in which the relevant OPHG population could not be separately extracted (n=9), studies in which the RT/SRS-treated subgroup could not be separately extracted (n=5), ineligible interventions (n=3), ineligible study designs or publication types (n=2), and absence of an eligible safety or efficacy outcome (n=1). Ultimately, **35 studies** were included in the systematic review, comprising 27 cohort studies, six case series, and two quasi-experimental studies (Fig. 1).

**Figure 1.**
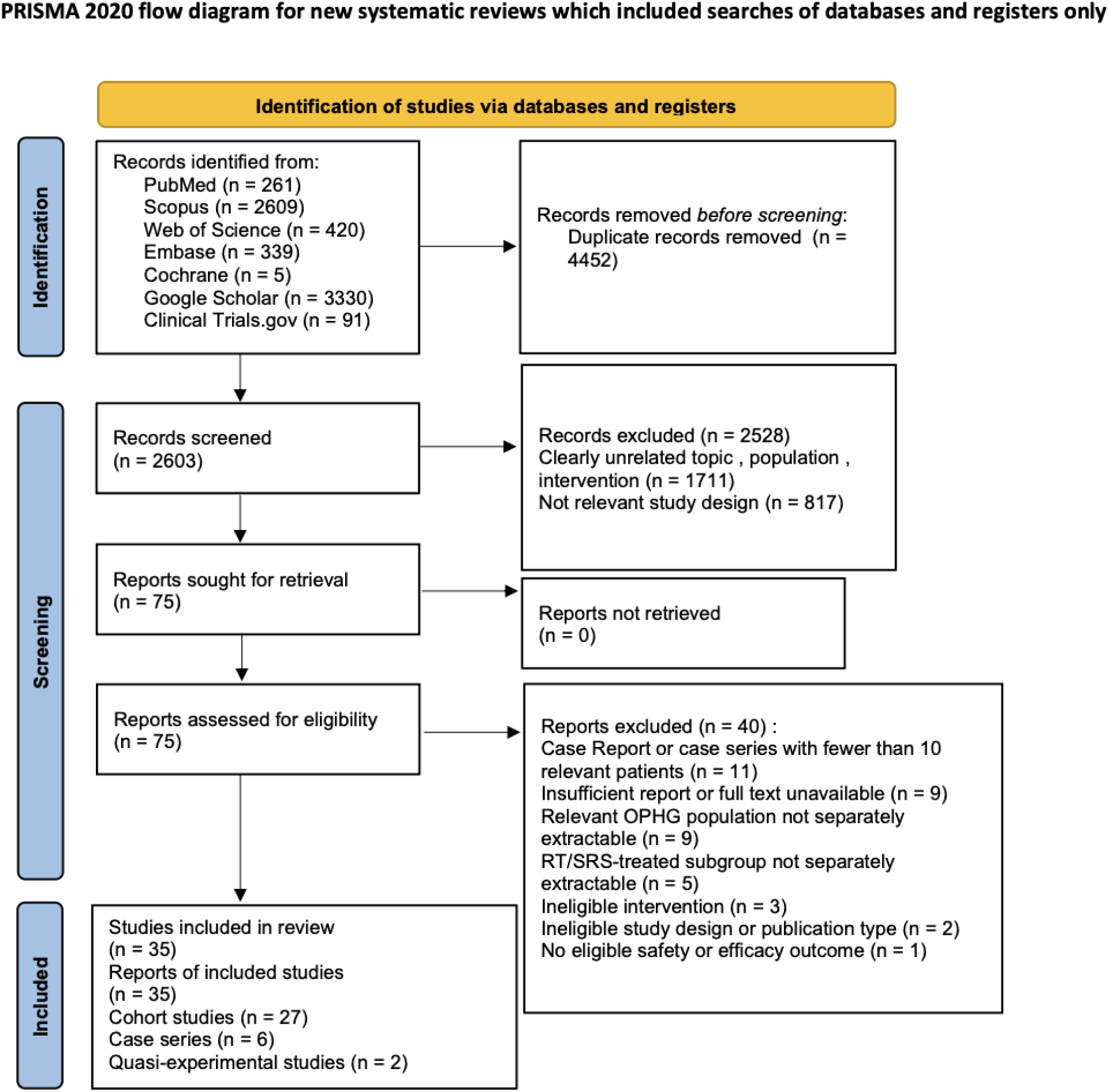
PRISMA 2020 flow diagram of study selection. PRISMA 2020 flow diagram illustrating study identification, screening, eligibility assessment, and final inclusion. The searches identified 7,055 records across the prespecified databases and supplementary sources. After removal of 4,452 duplicates, 2,603 unique records underwent title and abstract screening, 75 reports proceeded to full-text assessment, and 35 studies were ultimately included in the systematic review. The final evidence base comprised 27 cohort studies, six case series, and two quasi-experimental studies.

### Study and patient characteristics

The included studies were published between **1977 and 2024** and represented 13 countries, reflecting several decades of evolution in the management of OPHG. The United States contributed the largest number of studies (n=9), followed by the United Kingdom and Germany (n=5 each) and Canada (n=4); the remaining studies originated from France, Turkey, China, Saudi Arabia, Australia, the Netherlands, South Korea, Egypt, and Russia. The evidence base was predominantly retrospective: 27 studies were retrospective, two were prospective, one incorporated both retrospective and prospective recruitment, and the direction of data collection was not clearly reported in five studies. No randomized trial was identified.

Eligible OPHG cohorts ranged from **4 to 198 patients**, although the number of patients actually contributing to individual safety analyses was frequently smaller because several studies contained mixed treatment groups or reported only selected outcomes for the irradiated subgroup. Most studies involved pediatric or predominantly pediatric populations, whereas several included broader age ranges extending into adolescence or adulthood. Sex was separately reported for the eligible population in 31 studies, and NF1 status was reported in 32. Both sporadic and NF1-associated OPHG were therefore represented throughout the evidence base. Reported follow-up was highly variable; among studies providing a numeric median, follow-up ranged from approximately **30 to 206 months**, while the longest reported follow-up in individual patients extended beyond 50 years.

Radiation exposure was similarly heterogeneous and encompassed historical orthovoltage and cobalt-based external-beam radiotherapy, conventional megavoltage photon radiotherapy, two-and three-dimensional conformal techniques, intensity-modulated radiotherapy, proton therapy, fractionated stereotactic radiotherapy, single-session and fractionated Gamma Knife radiosurgery, CyberKnife treatment, and iodine-125 interstitial brachytherapy. Consequently, the review incorporated both historical treatment paradigms and substantially more conformal contemporary techniques. The principal study, patient, and treatment characteristics are summarized in **Table 1**.

**Table 1.**
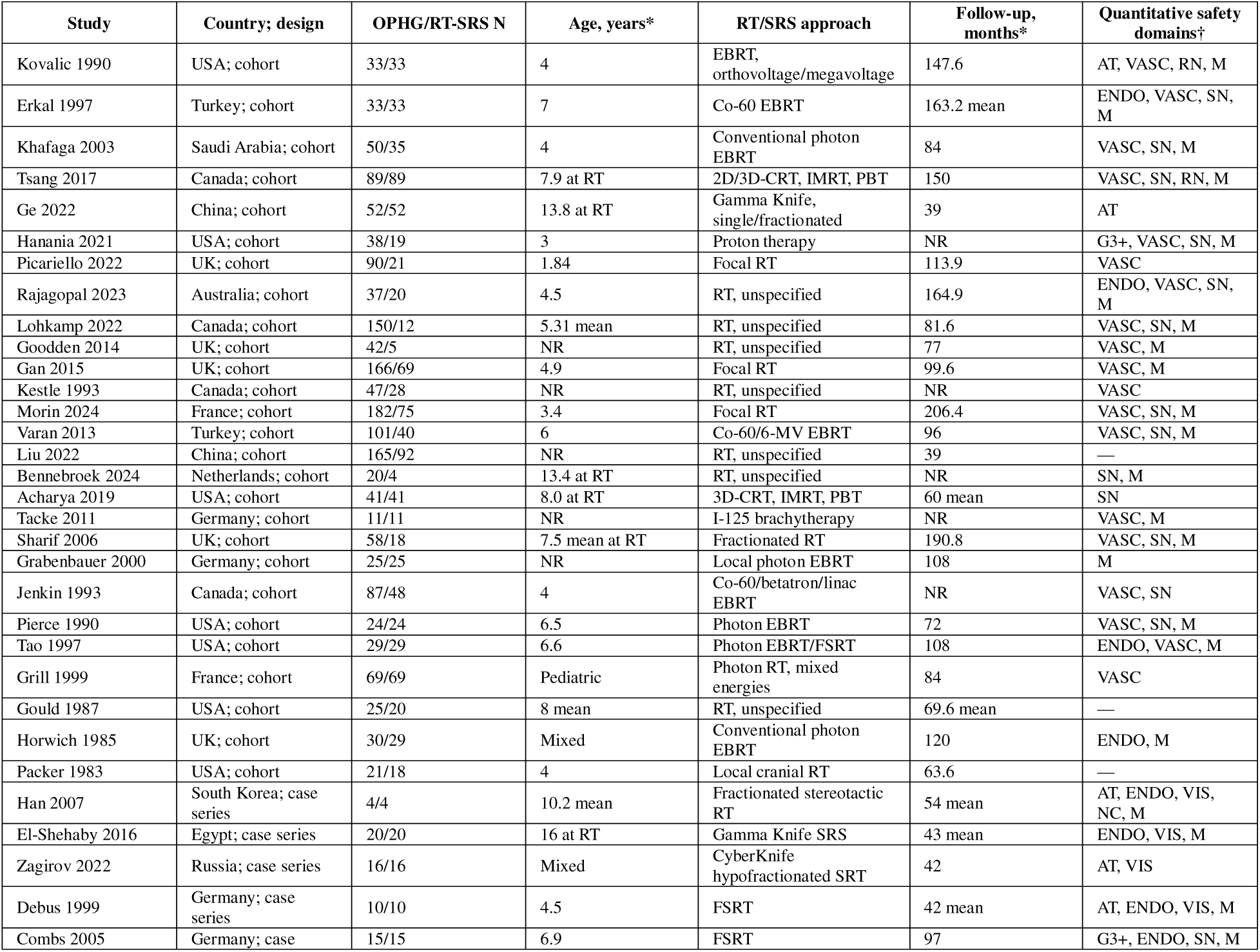
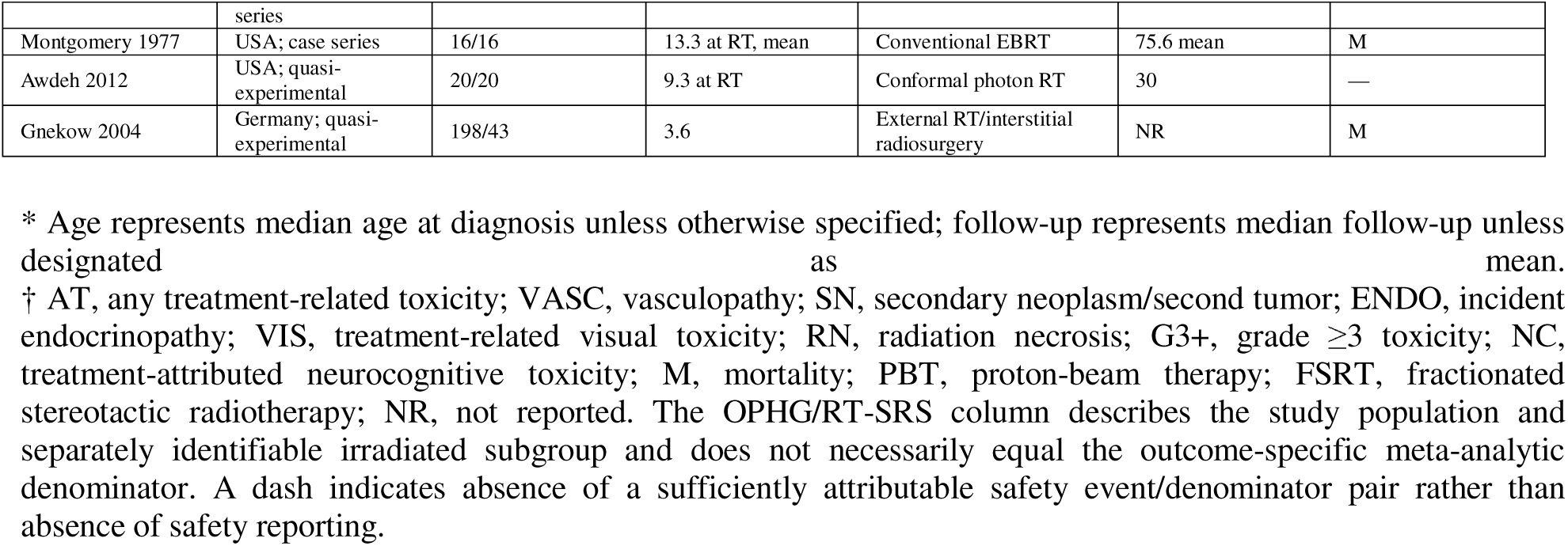
Characteristics of studies included in the systematic review of radiotherapy and radiosurgery safety for optic pathway–hypothalamic glioma. Values represent the separately identifiable OPHG population and, where applicable, the subgroup exposed to radiotherapy or radiosurgery. Age and follow-up are presented as reported by the original studies; medians are shown unless otherwise specified. The number of patients in the RT/SRS subgroup does not necessarily equal the denominator used for an individual quantitative safety outcome because outcome reporting and evaluability varied across studies. AT, any treatment-related toxicity; VASC, vasculopathy; SN, secondary neoplasm/second tumor; ENDO, incident endocrinopathy; VIS, treatment-related visual toxicity; RN, radiation necrosis; G3+, grade ≥3 toxicity; NC, treatment-attributed neurocognitive toxicity; EBRT, external-beam radiotherapy; 3D-CRT, three-dimensional conformal radiotherapy; IMRT, intensity-modulated radiotherapy; PBT, proton-beam therapy; FSRT, fractionated stereotactic radiotherapy; SRS, stereotactic radiosurgery; NF1, neurofibromatosis type 1; NR, not reported.

| Study | Country; design | OPHG/RT-SRS N | Age, years* | RT/SRS approach | Follow-up, months* | Quantitative safety domains† |
| --- | --- | --- | --- | --- | --- | --- |
| Kovalic 1990 | USA; cohort | 33/33 | 4 | EBRT, orthovoltage/megavoltage | 147.6 | AT, VASC, RN, M |
| Erkal 1997 | Turkey; cohort | 33/33 | 7 | Co-60 EBRT | 163.2 mean | ENDO, VASC, SN, M |
| Khafaga 2003 | Saudi Arabia; cohort | 50/35 | 4 | Conventional photon EBRT | 84 | VASC, SN, M |
| Tsang 2017 | Canada; cohort | 89/89 | 7.9 at RT | 2D/3D-CRT, IMRT, PBT | 150 | VASC, SN, RN, M |
| Ge 2022 | China; cohort | 52/52 | 13.8 at RT | Gamma Knife, single/fractionated | 39 | AT |
| Hanania 2021 | USA; cohort | 38/19 | 3 | Proton therapy | NR | G3+, VASC, SN, M |
| Picariello 2022 | UK; cohort | 90/21 | 1.84 | Focal RT | 113.9 | VASC |
| Rajagopal 2023 | Australia; cohort | 37/20 | 4.5 | RT, unspecified | 164.9 | ENDO, VASC, SN, M |
| Lohkamp 2022 | Canada; cohort | 150/12 | 5.31 mean | RT, unspecified | 81.6 | VASC, SN, M |
| Goodden 2014 | UK; cohort | 42/5 | NR | RT, unspecified | 77 | VASC, M |
| Gan 2015 | UK; cohort | 166/69 | 4.9 | Focal RT | 99.6 | VASC, M |
| Kestle 1993 | Canada; cohort | 47/28 | NR | RT, unspecified | NR | VASC |
| Morin 2024 | France; cohort | 182/75 | 3.4 | Focal RT | 206.4 | VASC, SN, M |
| Varan 2013 | Turkey; cohort | 101/40 | 6 | Co-60/6-MV EBRT | 96 | VASC, SN, M |
| Liu 2022 | China; cohort | 165/92 | NR | RT, unspecified | 39 | — |
| Bennebroek 2024 | Netherlands; cohort | 20/4 | 13.4 at RT | RT, unspecified | NR | SN, M |
| Acharya 2019 | USA; cohort | 41/41 | 8.0 at RT | 3D-CRT, IMRT, PBT | 60 mean | SN |
| Tacke 2011 | Germany; cohort | 11/11 | NR | I-125 brachytherapy | NR | VASC, M |
| Sharif 2006 | UK; cohort | 58/18 | 7.5 mean at RT | Fractionated RT | 190.8 | VASC, SN, M |
| Grabenbauer 2000 | Germany; cohort | 25/25 | NR | Local photon EBRT | 108 | M |
| Jenkin 1993 | Canada; cohort | 87/48 | 4 | Co-60/betatron/linac EBRT | NR | VASC, SN |
| Pierce 1990 | USA; cohort | 24/24 | 6.5 | Photon EBRT | 72 | VASC, SN, M |
| Tao 1997 | USA; cohort | 29/29 | 6.6 | Photon EBRT/FSRT | 108 | ENDO, VASC, M |
| Grill 1999 | France; cohort | 69/69 | Pediatric | Photon RT, mixed energies | 84 | VASC |
| Gould 1987 | USA; cohort | 25/20 | 8 mean | RT, unspecified | 69.6 mean | — |
| Horwich 1985 | UK; cohort | 30/29 | Mixed | Conventional photon EBRT | 120 | ENDO, M |
| Packer 1983 | USA; cohort | 21/18 | 4 | Local cranial RT | 63.6 | — |
| Han 2007 | South Korea; case series | 4/4 | 10.2 mean | Fractionated stereotactic RT | 54 mean | AT, ENDO, VIS, NC, M |
| El-Shehaby 2016 | Egypt; case series | 20/20 | 16 at RT | Gamma Knife SRS | 43 mean | ENDO, VIS, M |
| Zagirov 2022 | Russia; case series | 16/16 | Mixed | CyberKnife hypofractionated SRT | 42 | AT, VIS |
| Debus 1999 | Germany; case series | 10/10 | 4.5 | FSRT | 42 mean | AT, ENDO, VIS, M |
| Combs 2005 | Germany; case | 15/15 | 6.9 | FSRT | 97 | G3+, ENDO, SN, M |

|  | series |  |  |  |  |  |
| --- | --- | --- | --- | --- | --- | --- |
| Montgomery 1977 | USA; case series | 16/16 | 13.3 at RT, mean | Conventional EBRT | 75.6 mean | M |
| Awdeh 2012 | USA; quasi-experimental | 20/20 | 9.3 at RT | Conformal photon RT | 30 | — |
| Gnekow 2004 | Germany; quasi-experimental | 198/43 | 3.6 | External RT/interstitial radiosurgery | NR | M |

\* Age represents median age at diagnosis unless otherwise specified; follow-up represents median follow-up unless designated as mean.
† AT, any treatment-related toxicity; VASC, vasculopathy; SN, secondary neoplasm/second tumor; ENDO, incident endocrinopathy; VIS, treatment-related visual toxicity; RN, radiation necrosis; G3+, grade $\geq 3$ toxicity; NC, treatment-attributed neurocognitive toxicity; M, mortality; PBT, proton-beam therapy; FSRT, fractionated stereotactic radiotherapy; NR, not reported. The OPHG/RT-SRS column describes the study population and separately identifiable irradiated subgroup and does not necessarily equal the outcome-specific meta-analytic denominator. A dash indicates absence of a sufficiently attributable safety event/denominator pair rather than absence of safety reporting.

### Availability of safety outcomes

Safety reporting was uneven across the evidence base. **Thirty-one of the 35 included studies provided an interpretable event count and denominator for at least one quantitative safety outcome**, whereas the remaining studies contributed to qualitative characterization, treatment outcomes, or the broader evidence architecture without providing a sufficiently attributable RT/SRS safety numerator and denominator. Accordingly, the number of contributing studies varied substantially across outcomes rather than all 35 studies entering a single pooled analysis.

Vasculopathy and mortality were among the most frequently quantifiable safety domains, whereas treatment-related visual toxicity, radiation necrosis, severe toxicity, and treatment-attributed neurocognitive toxicity were substantially more sparsely reported. This uneven distribution was explicitly retained rather than treating unreported outcomes as zero-event observations.

### Risk of bias

Using the design-specific JBI critical appraisal instruments, **12 studies were classified as low risk of bias, 17 as moderate risk, and six as high risk** according to the prespecified operational study-level interpretation (Fig. 7; Supplementary File 7). The studies classified as low risk were **Erkal 1997, Picariello 2022, Lohkamp 2022, Gan 2015, Kestle 1993, Morin 2024, Acharya 2019, Tao 1997, El-Shehaby 2016, Zagirov 2022, Debus 1999, and Combs 2005. Moderate-**risk studies were **Kovalic 1990, Khafaga 2003, Tsang 2017, Hanania 2021, Rajagopal 2023,Goodden 2014, Bennebroek 2024, Tacke 2011, Sharif 2006, Grabenbauer 2000, Grill 1999, Horwich 1985, Packer 1983, Han 2007, Montgomery 1977, Awdeh 2012, and Gnekow 2004**. The six high-risk studies were **Ge 2022, Varan 2013, Liu 2022, Jenkin 1993, Pierce 1990, and Gould 1987.**

The traffic-light assessment displays item-level JBI judgments separately for cohort studies, case series, and quasi-experimental studies, together with the overall operational classification (Fig. 7).

### Quantitative synthesis

#### Primary outcome: any treatment-related toxicity

Five studies comprising **115 RT/SRS-treated patients and 23 treatment-related toxicity events** provided sufficiently attributable data for the primary outcome. The random-effects binomial-normal GLMM yielded a pooled incidence of **8.46% (95% CI, 1.37–38.01%)**, with substantial between-study heterogeneity (**I^2^=83.2%;** τ**^2^=2.5542**). The 95% prediction interval was correspondingly broad (**0.24–78.22%**) (Fig. 2). Individual observed incidences ranged from 0% to 54.5%, highlighting marked differences in toxicity definition, treatment era, and ascertainment among the contributing studies.

**Figure 2.**
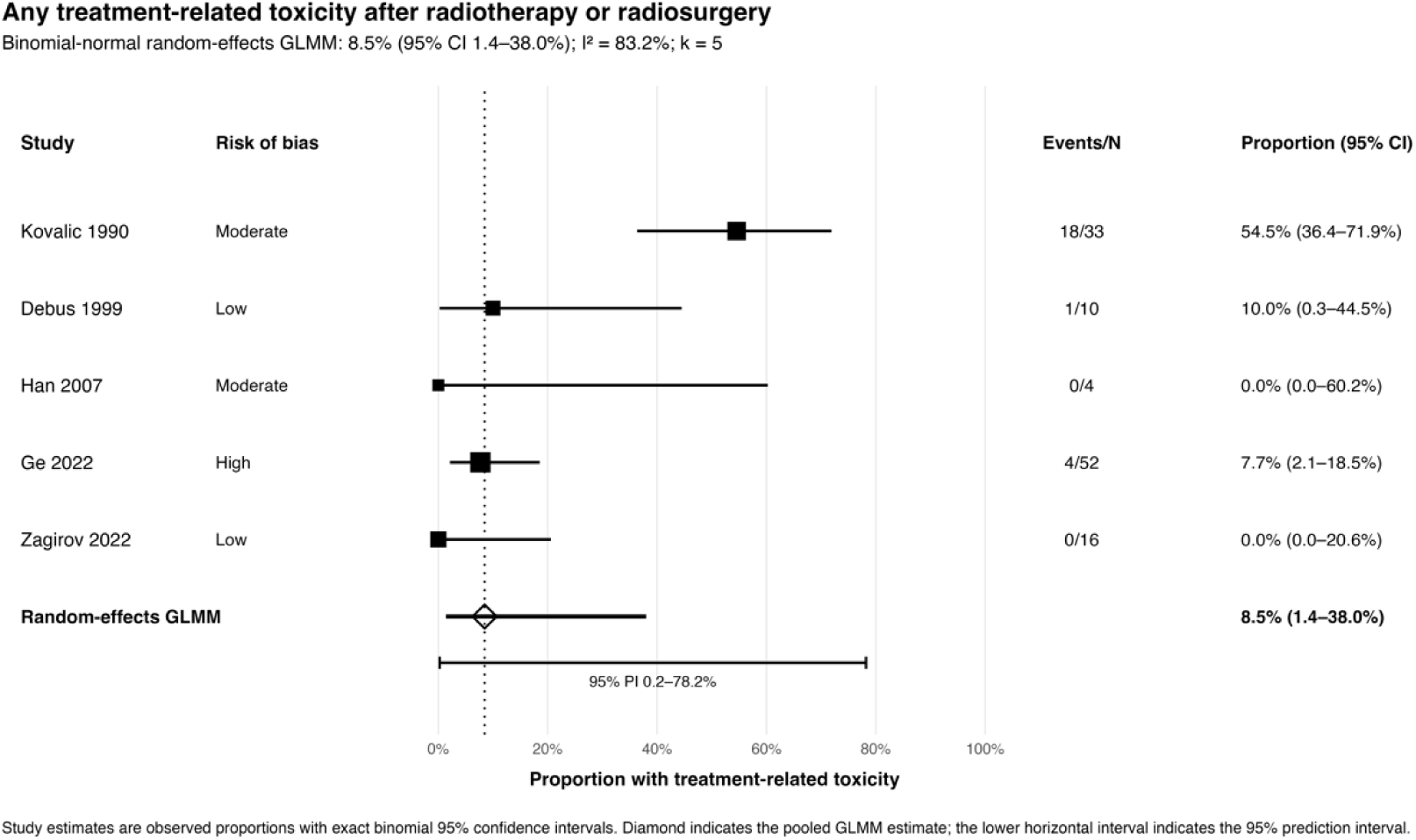
Pooled incidence of any treatment-related toxicity following radiotherapy or radiosurgery for optic pathway–hypothalamic glioma. Forest plot of five studies comprising 115 RT/SRS-treated patients and 23 treatment-related toxicity events. Study-specific proportions are presented with 95% confidence intervals (CIs). The pooled estimate was obtained using a binomial-normal random-effects generalized linear mixed-effects model (GLMM) and was 8.46% (95% CI, 1.37–38.01%). Between-study heterogeneity was substantial (I^2^=83.2%; τ^2^=2.5542), and the 95% prediction interval was 0.24–78.22%.

Sensitivity analyses did not indicate that the overall finding was driven exclusively by a single analytic assumption, although the magnitude decreased when analysis was restricted to more contemporary and/or lower-risk studies. Exclusion of the high-risk study yielded a pooled estimate of 6.7% (95% CI, 0.4–58.3%); a modern-treatment restriction yielded 6.1% (95% CI, 2.6–13.8%), and simultaneously restricting to modern and non-high-risk studies yielded 3.3% (95% CI, 0.5–20.2%) (Supplementary Fig. S1). Sequential study omission yielded pooled estimates from 6.1% to 14.7%. Because only five studies contributed, funnel and regression-based asymmetry displays were treated as descriptive; leave-one-out, influence, and study-level stratification results are presented in Supplementary Figures S2–S4.

### Vasculopathy

After resolving known or probable cohort overlap, 16 studies comprising 564 patients and 67 vasculopathy events contributed to the primary vasculopathy analysis. The pooled incidence was 9.44% (95% CI, 5.22–16.49%), with substantial heterogeneity (I^2^=77.2%; τ^2^=1.1060) and a 95% prediction interval of 1.19–47.43% (Fig. 3).

**Figure 3.**
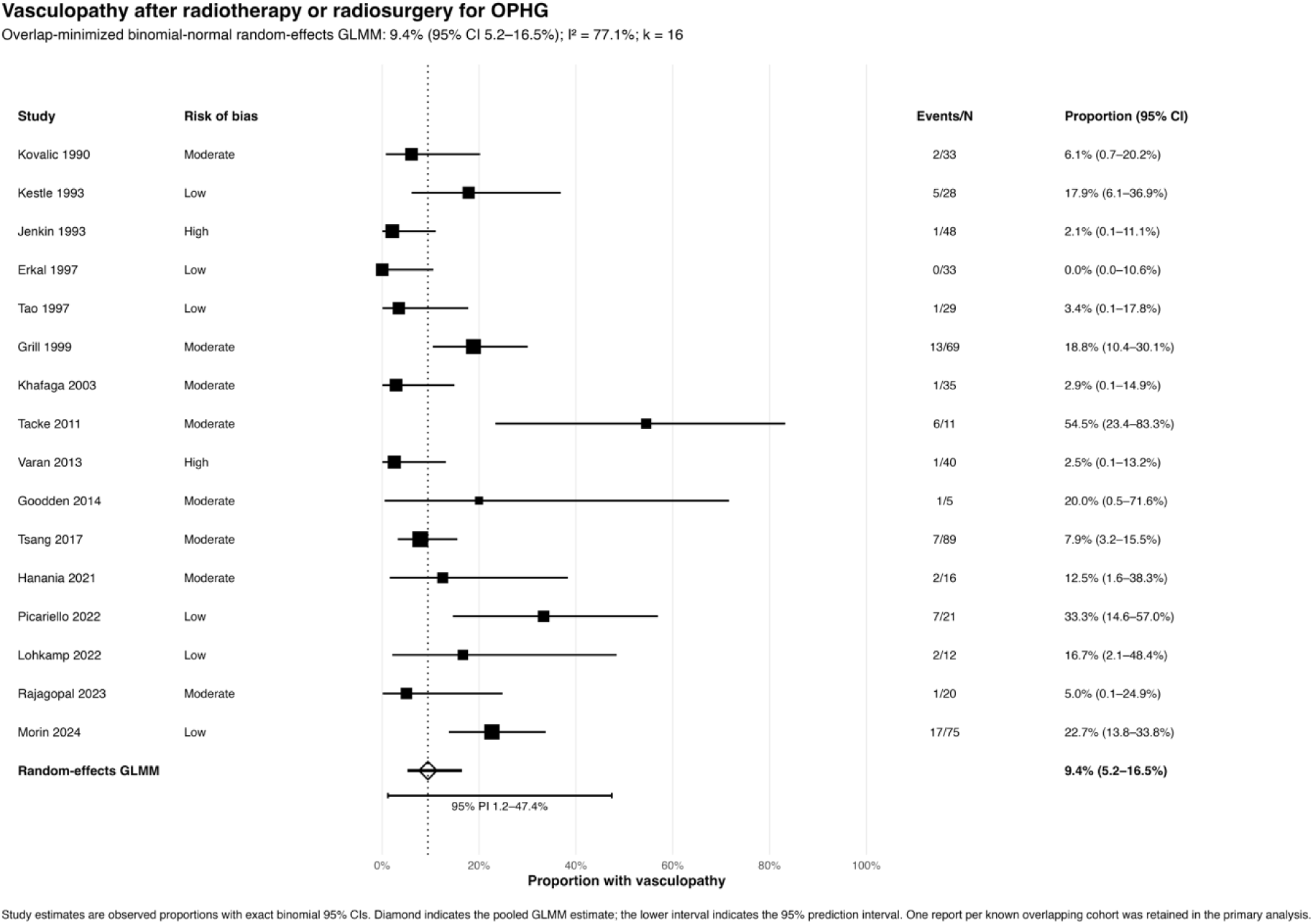
Vasculopathy following radiotherapy or radiosurgery for optic pathway–hypothalamic glioma. Overlap-minimized random-effects GLMM of vasculopathy. Sixteen studies comprising 564 patients and 67 events yielded a pooled incidence of 9.44% (95% CI, 5.22–16.49%), with I^2^=77.2%, τ^2^=1.1060, and a 95% prediction interval of 1.19–47.43%. One report per known overlapping cohort was retained in the primary analysis.

The estimate was comparatively robust to alternative cohort-selection and ascertainment decisions. Inclusion of all eligible reports yielded 9.3% (95% CI, 5.5–15.2%), an alternative resolution of overlapping cohorts yielded 8.9% (95% CI, 5.3–14.8%), exclusion of high-risk-of-bias studies yielded 11.6% (95% CI, 6.6–19.6%), and exclusion of the study using systematic MRA screening yielded 8.4% (95% CI, 4.8–14.2%) (Supplementary Fig. S5). Sequential study omission yielded pooled estimates from 8.4% to 10.7% (Supplementary Fig. S6).

Systematic MRA screening was associated with a notably higher observed event proportion than routine or clinically triggered vascular follow-up, although only one study employed systematic MRA screening and this comparison was therefore descriptive. Complementary diagnostics identified regression-based funnel asymmetry (p=0.023), but this was not interpreted as evidence of publication bias because clinical heterogeneity and differences in vascular surveillance can generate the same pattern (Supplementary Figs. S7–S8).

### Secondary neoplasms and second tumors

Thirteen non-overlapping studies comprising **469 patients and 41 secondary neoplasms or second tumors** contributed to the broad primary analysis. The pooled incidence was **5.41% (95% CI, 2.23–12.53%)**, with substantial heterogeneity (**I^2^=79.7%;** τ**^2^=1.6024**) and a 95% prediction interval of **0.40–44.61%** (Fig. 4A).

**Figure 4.**
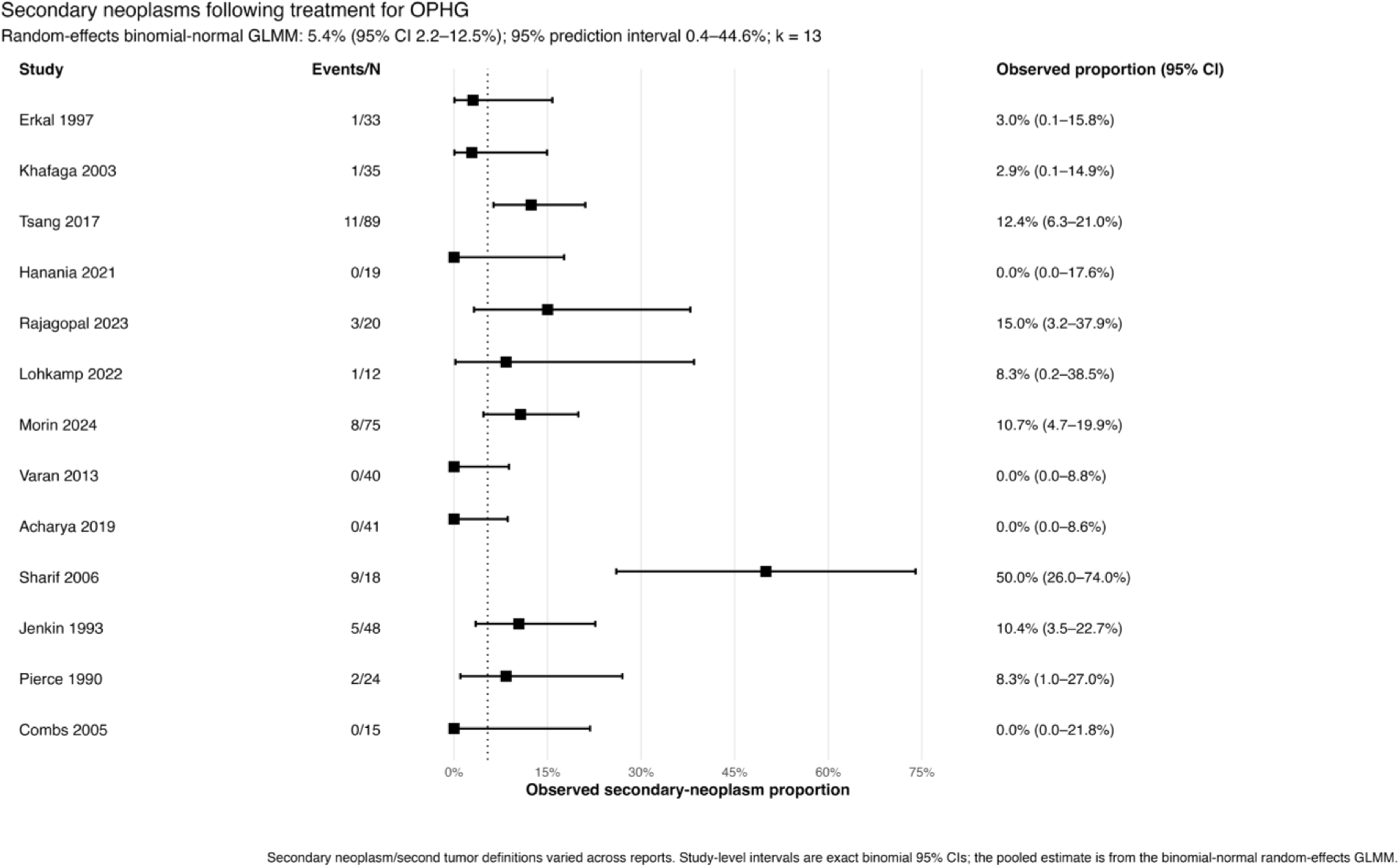
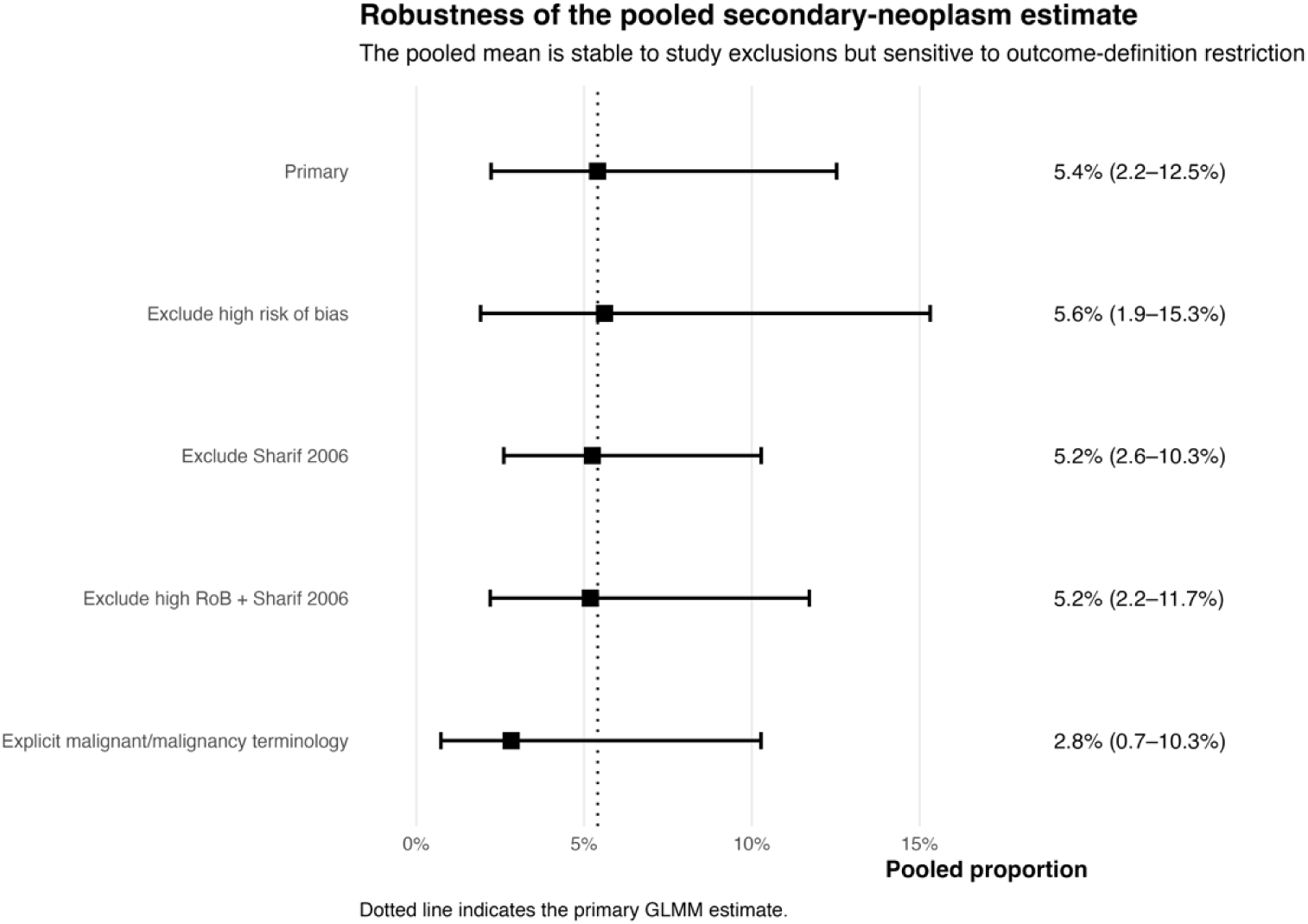
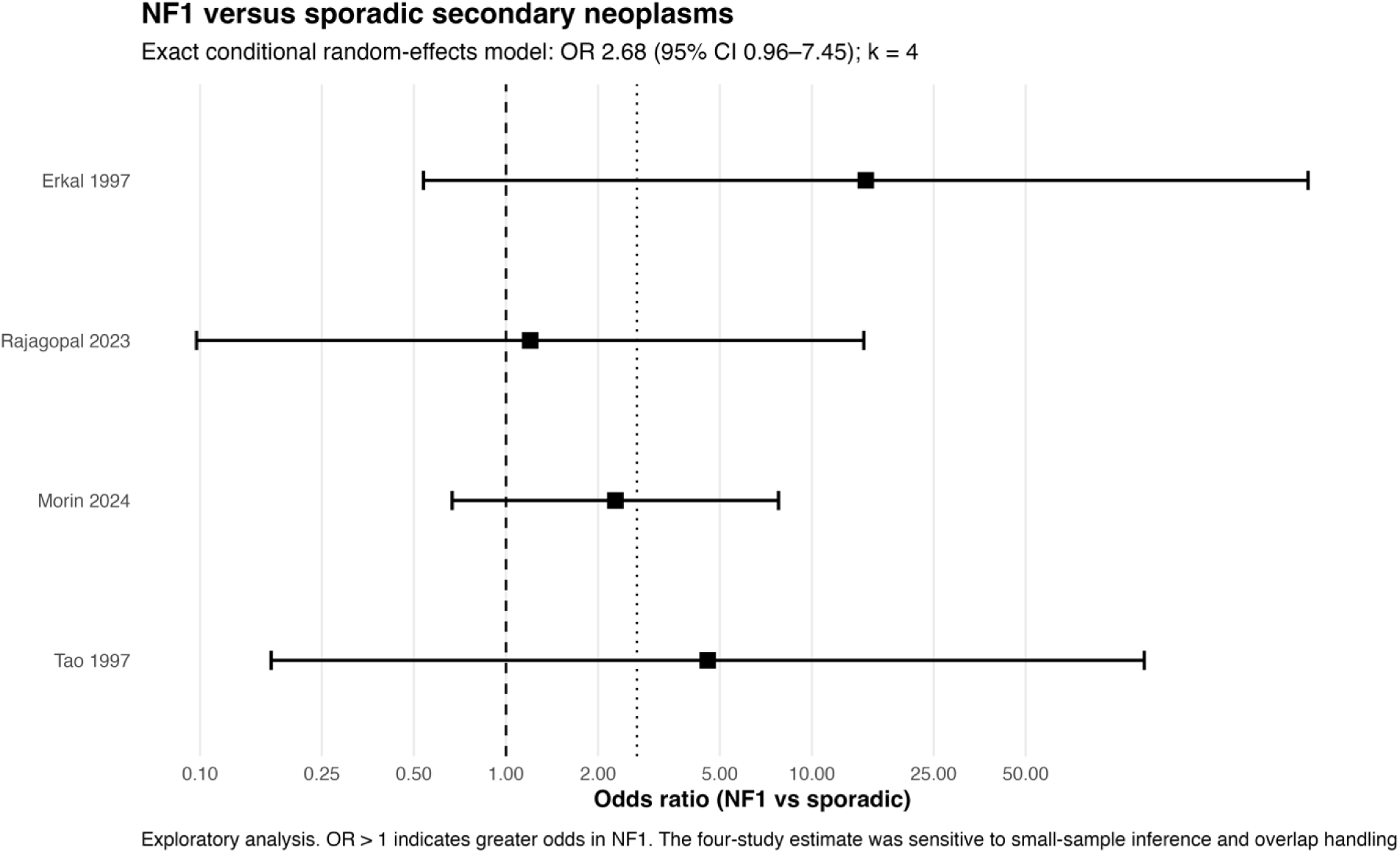
Secondary neoplasms following radiotherapy or radiosurgery for optic pathway–hypothalamic glioma. (A) Primary random-effects analysis of secondary neoplasms or second tumors. Thirteen non-overlapping studies comprising 469 patients and 41 events yielded a pooled incidence of 5.41% (95% CI, 2.23–12.53%), with I^2^=79.7%, τ^2^=1.6024, and a 95% prediction interval of 0.40–44.61%. (B) Sensitivity analyses examining exclusion of high-risk-of-bias studies, exclusion of the influential Sharif 2006 cohort, simultaneous application of both restrictions, and restriction to events explicitly described as malignant. The malignant-event sensitivity analysis yielded a pooled incidence of 2.83% (95% CI, 0.73–10.27%). (C) Exploratory comparison of NF1-associated versus sporadic disease based on four studies using an exact conditional random-effects model (OR, 2.68; 95% CI, 0.96–7.45).

The pooled mean was stable after exclusion of high-risk studies [5.6% (95% CI, 1.9–15.3%)], restriction to studies published from 2000 onward [4.3% (95% CI, 1.1–15.1%)], simultaneous modern-era and risk-of-bias restriction [6.0% (95% CI, 1.9–17.7%)], exclusion of the influential Sharif 2006 cohort [5.2% (95% CI, 2.6–10.3%)], and simultaneous exclusion of high-risk studies and Sharif 2006 [5.2% (95% CI, 2.2–11.7%)] (Fig. 4B; Supplementary Fig. S9). In contrast, restricting the outcome to events explicitly described using malignant or malignancy terminology reduced the estimate to 2.83% (95% CI, 0.73–10.27%) across eight studies and 285 patients, demonstrating sensitivity to outcome definition.

Four studies allowed comparison between NF1-associated and sporadic disease. The exact conditional random-effects estimate suggested greater odds of secondary neoplasms in patients with NF1 (**OR, 2.68; 95% CI, 0.96–7.45; p=0.059**), although the confidence interval included the null and inference remained sensitive to sparse data and overlap handling (Fig. 4C).

An additional exploratory comparison of irradiated versus non-irradiated patients yielded an equal-effects exact conditional **OR of 4.44 (95% CI, 1.86–10.57; p=0.0008)**. Because treatment allocation was non-randomized and potentially confounded by disease severity, age, treatment era, NF1 status, and follow-up, this estimate represents an **unadjusted observational association** rather than a causal effect of radiotherapy.

When analysis was restricted further to secondary neoplasms explicitly attributed to radiotherapy by the original investigators, six studies comprising 216 patients and nine events yielded a pooled incidence of 1.24% (95% CI, 0.09–14.57%), with a 95% prediction interval of 0.02–39.70%, τ^2^=2.307, and I^2^=68.7%. This more restrictive estimate illustrates the distinction between the incidence of second tumors following treatment and events for which radiation attribution was specifically asserted. Leave-one-out estimates ranged from 4.6% to 6.7%. Regression-based funnel asymmetry was detected (p=0.039), but the pattern was regarded as exploratory because it may reflect outcome-definition heterogeneity and sparse-event structure rather than publication bias (Supplementary Figs. S10–S12).

### Incident endocrinopathy

Seven studies comprising **139 patients and 50 incident endocrine events** provided sufficiently separable post-treatment data. The pooled incidence of new endocrinopathy was **21.19% (95% CI, 4.72–59.31%)**, with very substantial heterogeneity (**I^2^=90.9%;** τ**^2^=3.9754**) and a 95% prediction interval of **0.38–95.00%** (Fig. 5A). Study-level estimates ranged from no incident events in some stereotactic series to markedly higher incidences in cohorts with prolonged follow-up.

**Figure 5.**
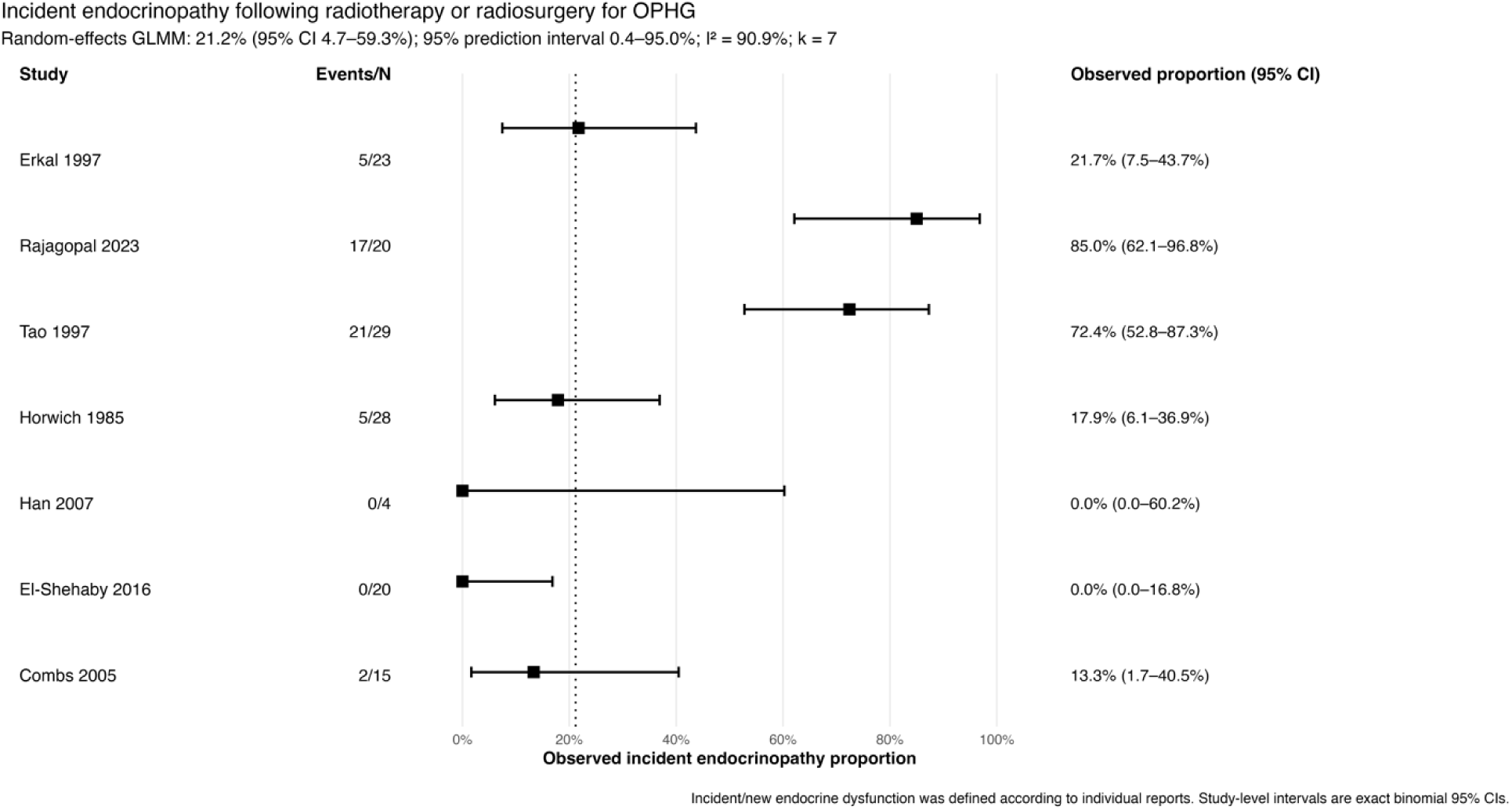
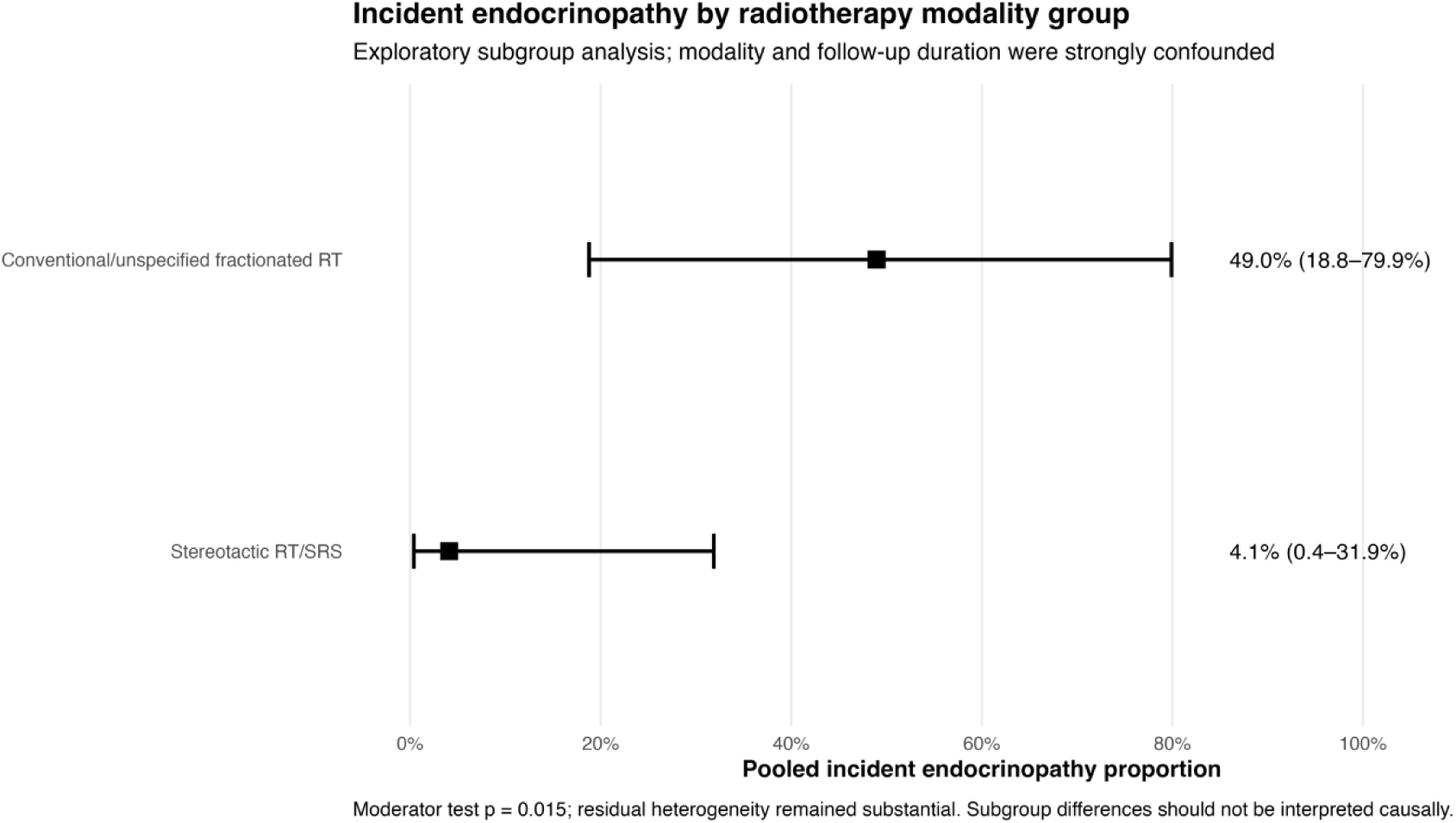
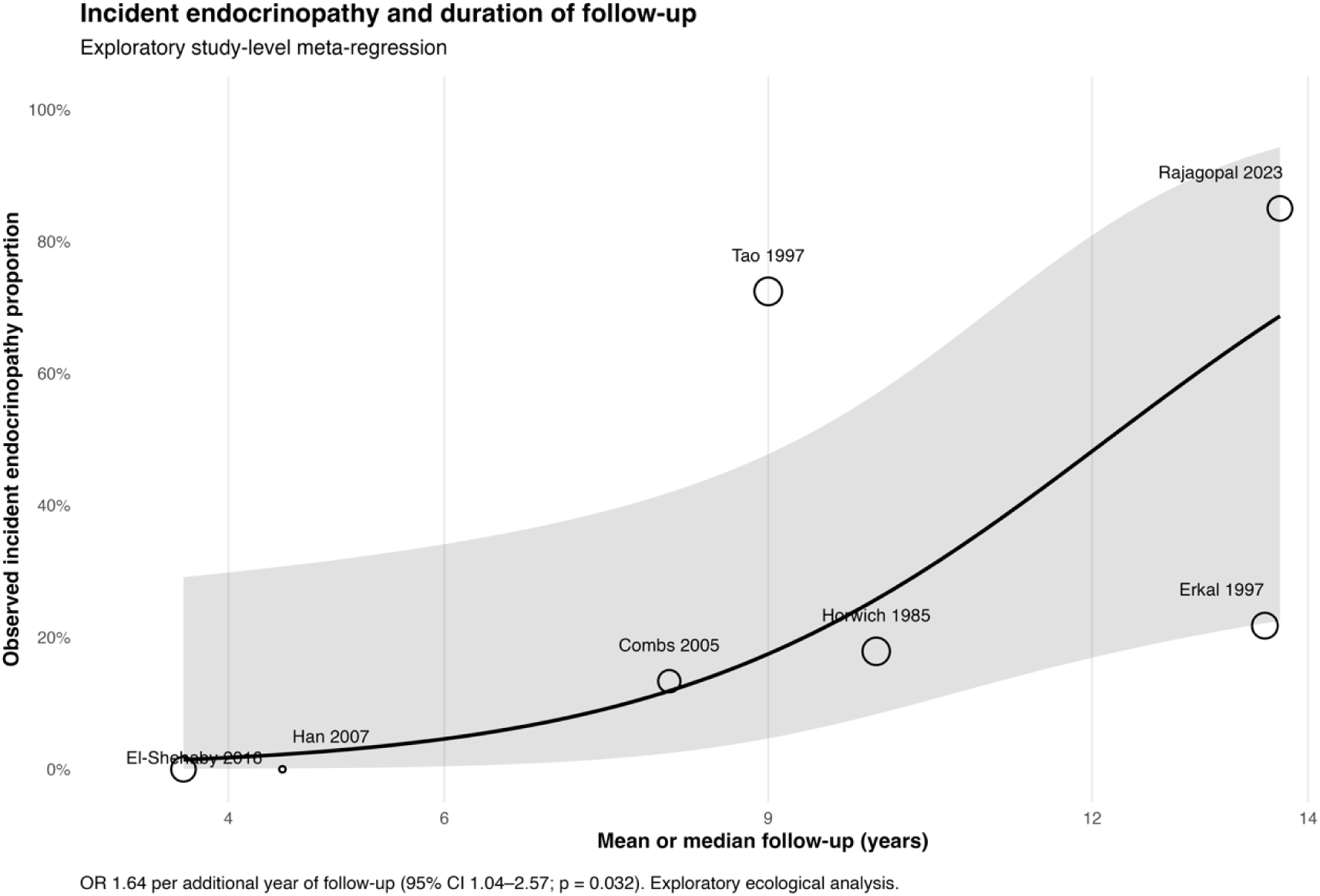
Incident endocrinopathy following radiotherapy or radiosurgery for optic pathway–hypothalamic glioma. (A) Random-effects GLMM of seven studies comprising 139 patients and 50 incident endocrine events. The pooled incidence was 21.19% (95% CI, 4.72–59.31%), with I^2^=90.9%, τ^2^=3.9754, and a 95% prediction interval of 0.38–95.00%. (B) Exploratory analysis according to treatment modality. The pooled incidence was 49.0% (95% CI, 18.8–79.9%) for conventional or unspecified fractionated radiotherapy and 4.1% (95% CI, 0.4–31.9%) for stereotactic RT/SRS; moderator p=0.015. (C) Study-level meta-regression according to mean or median follow-up duration. Longer follow-up was associated with greater reported incidence of endocrinopathy (OR, 1.64 per additional year; 95% CI, 1.04–2.57; p=0.032). Modality and follow-up analyses were exploratory and should not be interpreted as patient-level causal effects.

In an exploratory modality analysis, conventional or unspecified fractionated radiotherapy was associated with a pooled incident endocrinopathy proportion of **49.0% (95% CI, 18.8–79.9%)**, compared with **4.1% (95% CI, 0.4–31.9%)** among stereotactic RT/SRS series (moderator p=0.015) (Fig. 5B). However, modality was strongly confounded by treatment era and follow-up duration, and substantial residual heterogeneity remained; this finding was therefore not interpreted as evidence of a causal modality effect.

Consistent with the latency of endocrine sequelae, exploratory study-level meta-regression demonstrated increasing reported incidence with longer follow-up, corresponding to an OR of 1.64 per additional year of mean or median follow-up (95% CI, 1.04–2.57; p=0.032) (Fig. 5C). This association was ecological and should not be interpreted as a patient-level time-to-event estimate. The pooled estimate was 17.0% (95% CI, 2.6–61.6%) when restricted to low-risk-of-bias studies, 6.9% (95% CI, 0.1–79.9%) in studies published from 2000 onward, and 4.8% (95% CI, 0.6–31.5%) when both restrictions were applied. Leave-one-out estimates ranged from 14.7% to 32.8%. With seven contributing studies, asymmetry assessment was descriptive only (Supplementary Figs. S13–S16).

### Treatment-related visual toxicity

Four studies comprising 49 RT/SRS-treated patients reported sufficiently attributable visual toxicity data. Four treatment-related visual events were identified, producing a pooled incidence of 2.26% (95% CI, 0.03–65.95%), with τ^2^=3.0583 and I^2^=69.6%; the 95% prediction interval was 0.01–86.21% (Fig. 6A). All four events arose in El-Shehaby 2016 and consisted of transient visual worsening associated with post-treatment swelling. No permanent RT/SRS-attributed visual toxicity was identified among the 49 evaluable patients (0/49; exact 95% CI, 0–7.25%). Sensitivity estimates were 3.2% (95% CI, 0.1–62.0%) in low-risk-of-bias studies, 5.1% (95% CI, 0.2–56.7%) in studies published from 2000 onward, and 7.3% (95% CI, 0.6–51.6%) under the combined restriction. Leave-one-out estimates ranged from 3.2% to 10.7%; formal asymmetry testing was not undertaken with four studies (Supplementary Figs. S17–S20).

**Figure 6.**
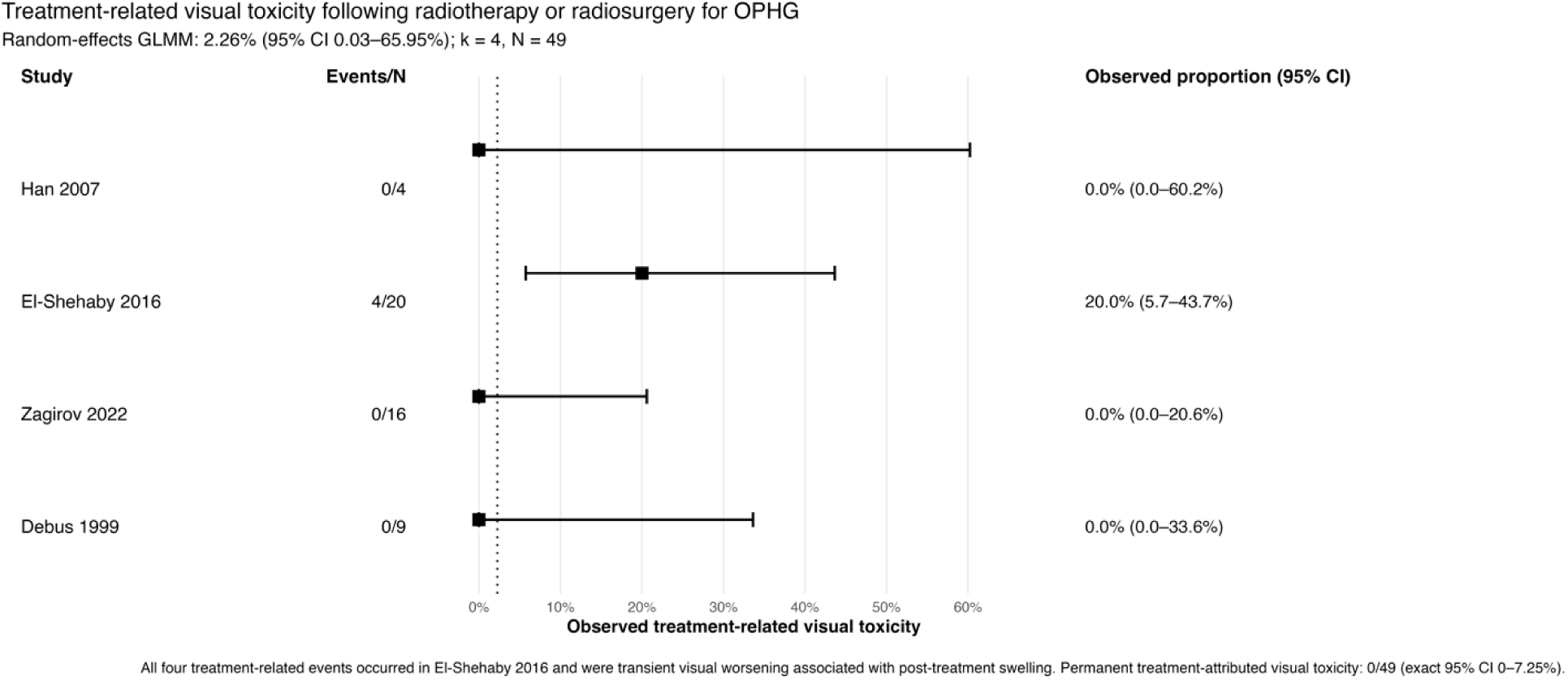
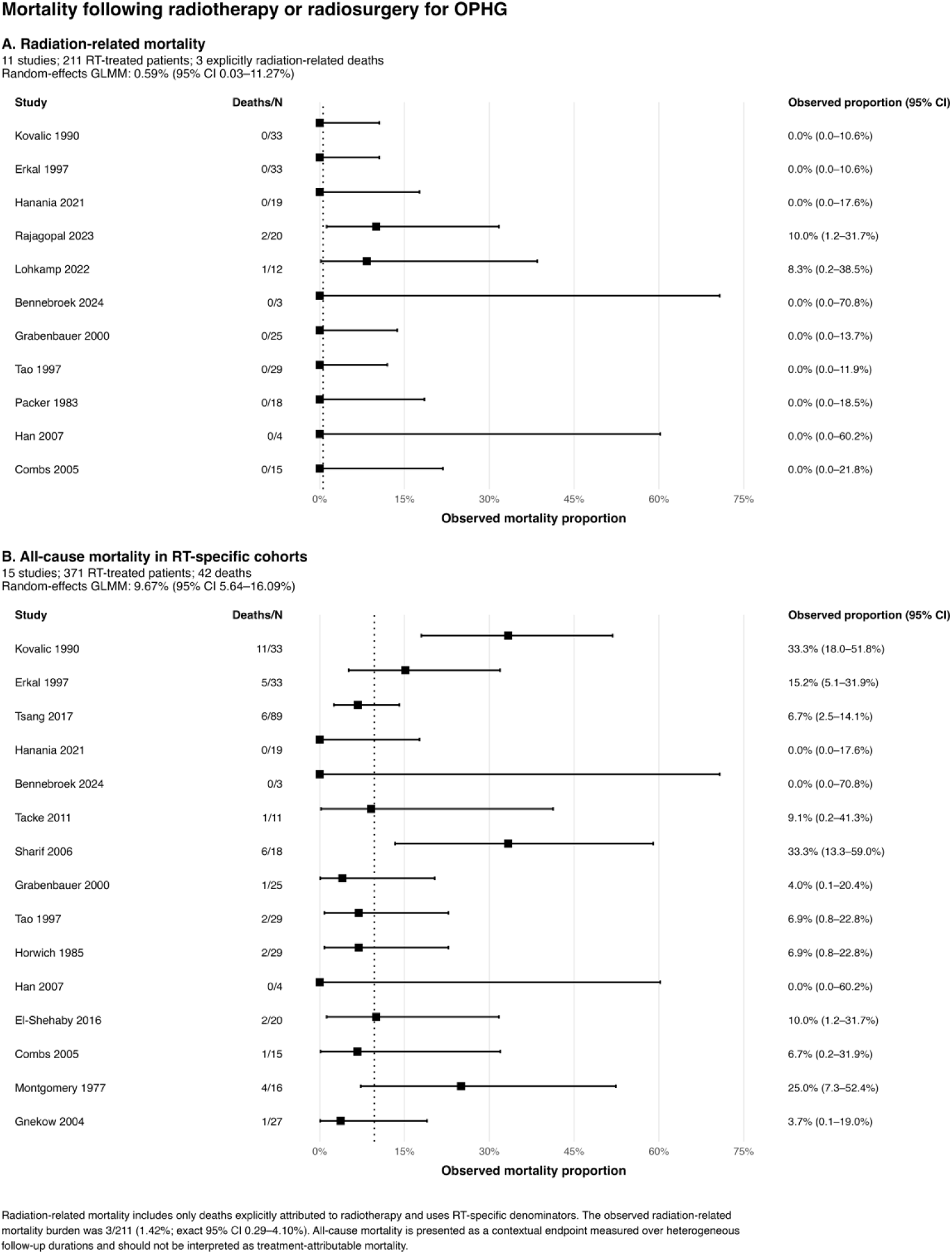
Treatment-related visual toxicity and mortality following radiotherapy or radiosurgery for optic pathway–hypothalamic glioma. (A) Treatment-related visual toxicity. Four studies comprising 49 patients and four attributable events yielded a pooled incidence of 2.26% (95% CI, 0.03–65.95%). All four events represented transient visual worsening associated with post-treatment swelling; no permanent treatment-attributed visual toxicity was identified (0/49; exact 95% CI, 0–7.25%). (B) Radiation-related mortality. Eleven studies comprising 211 RT-treated patients and three explicitly radiation-related deaths yielded a pooled incidence of 0.59% (95% CI, 0.03–11.27%). (C) All-cause mortality in RT-specific cohorts. Across 15 studies comprising 371 patients and 42 deaths, the pooled proportion was 9.67% (95% CI, 5.64–16.09%). All-cause mortality is presented as a contextual outcome and should not be interpreted as treatment-attributable mortality.

**Figure 7.**
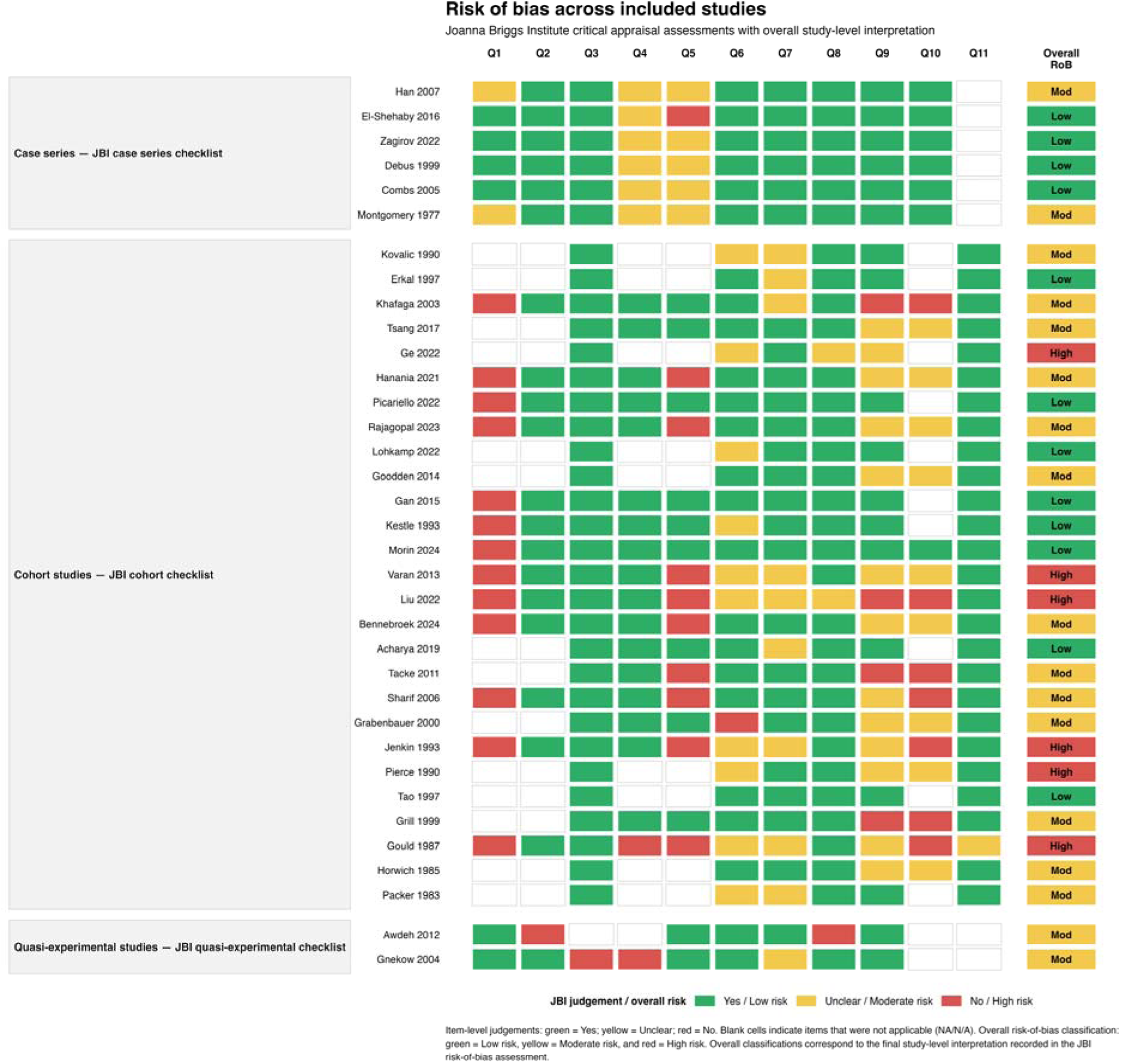
Risk-of-bias assessment of included studies. Traffic-light presentation of Joanna Briggs Institute (JBI) critical appraisal judgments according to study design, using the cohort, case-series, and quasi-experimental instruments as appropriate. Green, yellow, and red cells indicate “Yes,” “Unclear,” and “No” judgments, respectively; blank cells indicate items considered not applicable. The final column presents the prespecified operational study-level interpretation as low, moderate, or high risk of bias. Overall, 12 studies were classified as low risk, 17 as moderate risk, and six as high risk. These categories were used for transparent review-level presentation and do not represent official JBI cutoffs.

### Mortality

Eleven studies comprising 211 RT-treated patients reported mortality with sufficiently explicit attribution to radiotherapy. Three radiation-related deaths were identified. The random-effects pooled incidence was 0.59% (95% CI, 0.03–11.27%), with a 95% prediction interval of 0.01–30.15%, τ^2^=2.3368, and I^2^=59.0% (Fig. 6B). The corresponding unpooled observed proportion was 3/211 (1.42%; exact 95% CI, 0.29–4.10%). Sensitivity estimates were 1.0% (95% CI, 0.0– 27.0%) in low-risk-of-bias studies, 2.6% (95% CI, 0.5–13.6%) in studies published from 2000 onward, and 3.7% (95% CI, 0.5–22.1%) under the combined restriction. Leave-one-out estimates ranged from 0.07% to 0.86%. Regression-based asymmetry was detected (p=0.034), but was interpreted cautiously because the endpoint contained only three deaths and numerous zero-event studies (Supplementary Figs. S21–S24).

All-cause mortality was examined separately as a contextual endpoint. Across 15 RT-specific cohorts comprising 371 patients and 42 deaths, the pooled all-cause mortality proportion was 9.67% (95% CI, 5.64–16.09%), with a 95% prediction interval of 2.12–34.60%, τ^2^=0.5758, and I^2^=55.8% (Fig. 6C). Because deaths were observed across highly heterogeneous follow-up durations and were not necessarily attributable to treatment, this estimate was not interpreted as radiation-related mortality. The corresponding estimates were 10.3% (95% CI, 5.6–18.1%) in low-risk-of-bias studies, 6.8% (95% CI, 3.2–13.9%) in studies published from 2000 onward, and 8.6% (95% CI, 2.8–23.4%) under the combined restriction. Leave-one-out estimates ranged from 8.7% to 10.7%. Regression-based asymmetry was detected (p=0.030), but could not distinguish selective reporting from differences in follow-up, era, and clinical case mix (Supplementary Figs. S25–S28).

### Sparse safety outcomes

Several clinically important outcomes remained too sparsely reported for stable random-effects pooling. **Radiation necrosis** was reported in two studies comprising 122 patients, with no events observed (**0/122; 0%; exact 95% CI, 0–2.98%**). **Grade** ≥**3 toxicity** was available from two studies comprising 31 patients, with three events (**3/31; 9.68%; exact 95% CI, 2.04–25.75%**). Only one study provided an attributable denominator for **radiotherapy-related neurocognitive toxicity**, with no event among four patients (**0/4; exact 95% CI, 0–60.24%**). Neurological, hearing, growth, and metabolic adverse outcomes were additionally described in individual reports, but differences in attribution, definitions, and denominators precluded defensible quantitative pooling. The quantitative safety outcomes are summarized in Table 2.

**Table 2.** Summary of quantitative safety outcomes following radiotherapy or radiosurgery for optic pathway–hypothalamic glioma. Pooled proportions were estimated using binomial-normal random-effects generalized linear mixed-effects models unless otherwise specified. CI, 95% confidence interval; PI, 95% prediction interval; k, number of contributing studies; N, eligible outcome-specific denominator. Radiation necrosis, grade ≥3 toxicity, and treatment-attributed neurocognitive toxicity were too sparsely reported for stable random-effects pooling and are therefore presented descriptively with exact binomial 95% CIs. All-cause mortality is a contextual endpoint and should not be interpreted as treatment-attributable mortality. Estimates across outcomes should not be directly ranked because they derive from different study subsets, outcome definitions, treatment eras, and follow-up durations.

| Outcome | k | N | Events | Pooled estimate, %<br>(95% CI) | 95% PI, % | I<sup>2</sup> |
| --- | --- | --- | --- | --- | --- | --- |
| Any treatment-related toxicity | 5 | 115 | 23 | **8.46 (1.37–38.01)** | 0.24–78.22 | 83.2% |
| Vasculopathy | 16 | 564 | 67 | **9.44 (5.22–16.49)** | 1.19–47.43 | 77.2% |
| Secondary neoplasm/second tumor | 13 | 469 | 41 | **5.41 (2.23–12.53)** | 0.40–44.61 | 79.7% |
| Incident endocrinopathy | 7 | 139 | 50 | **21.19 (4.72–59.31)** | 0.38–95.00 | 90.9% |
| Treatment-related visual toxicity | 4 | 49 | 4 | **2.26 (0.03–65.95)** | 0.01–86.21 | 69.6% |
| Radiation-related mortality | 11 | 211 | 3 | **0.59 (0.03–11.27)** | 0.01–30.15 | 59.0% |
| All-cause mortality‡ | 15 | 371 | 42 | **9.67 (5.64–16.09)** | 2.12–34.60 | 55.8% |
| Radiation necrosis§ | 2 | 122 | 0 | **0.00 (0–2.98)** | — | — |
| Grade ≥3 toxicity§ | 2 | 31 | 3 | **9.68 (2.04–25.75)** | — | — |
| RT-attributed neurocognitive toxicity§ | 1 | 4 | 0 | **0.00 (0–60.24)** | — | — |

Random-effects binomial-normal GLMM estimates are reported for meta-analyzed endpoints.
‡ Contextual outcome; not interpreted as treatment-attributable mortality.
§ Sparse endpoints are reported descriptively with exact binomial 95% CIs rather than as random-effects pooled estimates.

The integrated magnitudes and heterogeneity statistics are also displayed in the final evidence-topology figure.

### Integrated evidence topology

All 35 included studies were subsequently represented in the prespecified study-by-outcome evidence matrix, irrespective of whether they contributed to a conventional pooled estimate. The resulting topology demonstrated a markedly uneven evidence architecture: vasculopathy had the broadest quantitative safety coverage (k=16), followed by all-cause mortality (k=15), secondary neoplasms (k=13), radiation-related mortality (k=11), incident endocrinopathy (k=7), any treatment-related toxicity (k=5), and visual toxicity (k=4), whereas radiation necrosis and grade ≥3 toxicity were each informed by only two studies and treatment-attributed neurocognitive toxicity by one.

Shared-study connectivity was evaluated using Jaccard similarity and stabilized across **5,000 Bayesian-bootstrap reweightings**. The exploratory five-cluster solution showed areas of shared evidence contribution across safety domains, but global community separation was modest. Accordingly, the network should be interpreted as a visualization of **which safety outcomes were investigated together within the same studies**, rather than as evidence that toxicities co-occurred in individual patients or shared biological mechanisms (Fig. 8).

**Figure 8.**
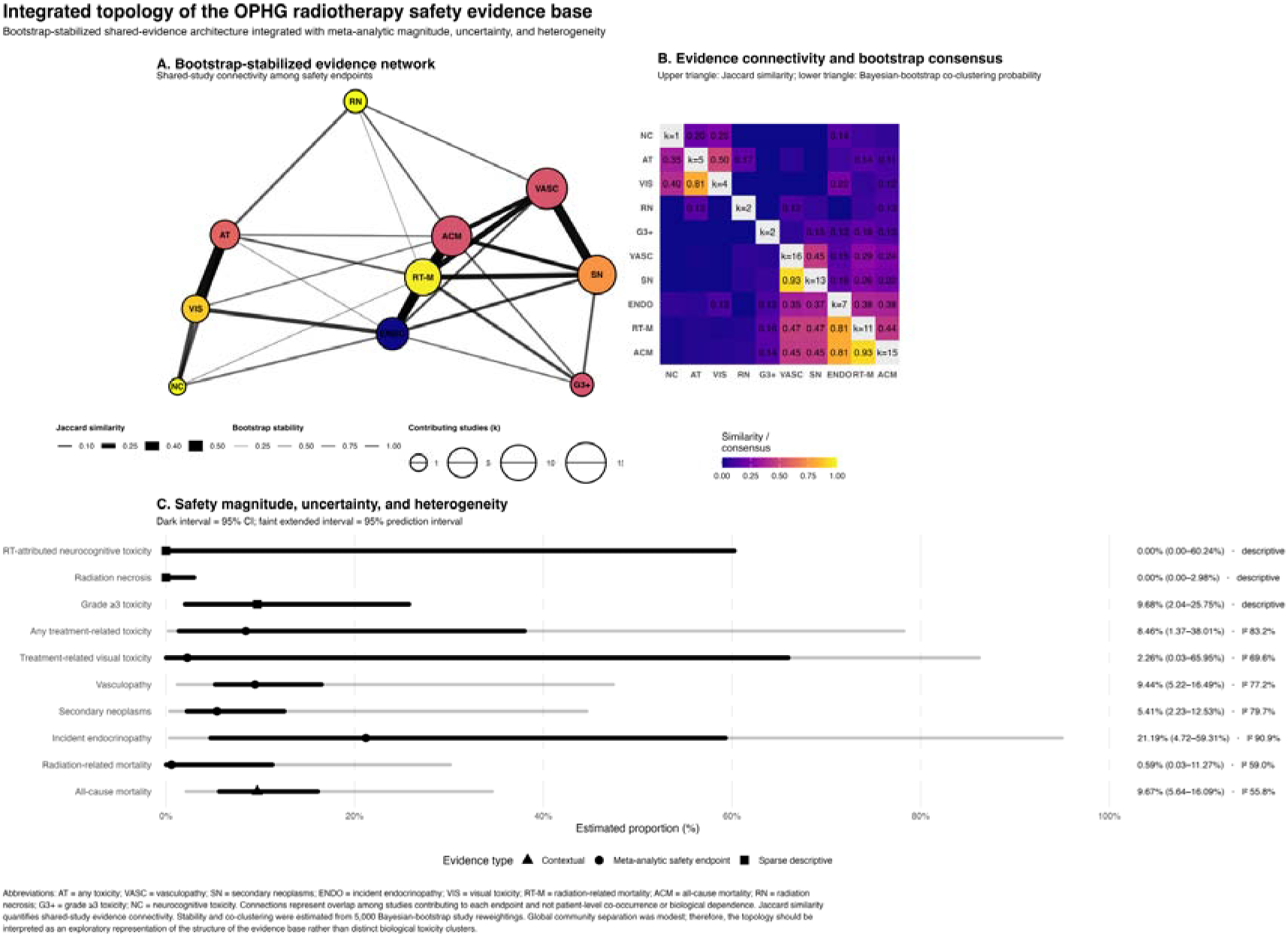
Integrated topology of the OPHG radiotherapy safety evidence base. Exploratory analysis integrating shared-study evidence architecture with meta-analytic magnitude, uncertainty, and heterogeneity across 10 safety domains. (A) Bootstrap-stabilized evidence network in which nodes represent safety outcomes and connections represent overlap among studies contributing evidence to each pair of outcomes. (B) Evidence-connectivity matrix. The upper triangle displays observed Jaccard similarity between outcome pairs, whereas the lower triangle displays Bayesian-bootstrap consensus co-clustering probabilities under the exploratory five-cluster solution. Edge stability represents the proportion of 5,000 Bayesian-bootstrap study reweightings in which the weighted Jaccard similarity exceeded 0.10. (C) Summary of pooled or descriptive safety estimates with 95% CIs, 95% prediction intervals where applicable, and heterogeneity statistics. Connections reflect shared study-level evidence availability and not patient-level co-occurrence, biological dependence among toxicities, or a network meta-analysis. AT, any treatment-related toxicity; VASC, vasculopathy; SN, secondary neoplasms; ENDO, incident endocrinopathy; VIS, treatment-related visual toxicity; RT-M, radiation-related mortality; ACM, all-cause mortality; RN, radiation necrosis; G3+, grade ≥3 toxicity; NC, treatment-attributed neurocognitive toxicity.

Taken together, the quantitative synthesis demonstrated that the largest pooled point estimate was observed for incident endocrinopathy (21.19%), followed by vasculopathy (9.44%), any treatment-related toxicity (8.46%), secondary neoplasms (5.41%), treatment-related visual toxicity (2.26%), and radiation-related mortality (0.59%). However, these estimates arose from different study subsets, outcome definitions, treatment eras, and follow-up durations and were accompanied by substantial heterogeneity and, for several outcomes, very wide prediction intervals. They therefore represent **outcome-specific estimates rather than a directly comparable hierarchy of radiation-related risks**.

## Discussion

This systematic review and meta-analysis provides a comprehensive synthesis of the safety profile of radiotherapy and radiosurgery for optic pathway–hypothalamic glioma (OPHG) across several decades of clinical practice. Several findings are particularly relevant. The pooled incidence of any treatment-related toxicity was 8.46%, although substantial heterogeneity and a wide prediction interval indicated marked variability across study populations and treatment eras. Vasculopathy emerged as one of the most consistently documented late complications, with a pooled incidence of 9.44%. Secondary neoplasms occurred in 5.41% of patients under the broad definition, decreasing to 2.83% when analysis was restricted to explicitly malignant events. Incident endocrinopathy had the highest pooled point estimate, at 21.19%, and its reported frequency increased with longer follow-up. In contrast, permanent treatment-attributed visual toxicity, radiation necrosis, and radiation-related mortality were infrequently documented. Importantly, we adjudicated safety outcomes separately, distinguished incident from pre-existing abnormalities whenever possible, resolved potentially overlapping cohorts before pooling, and avoided interpreting unreported toxicities as zero events. The resulting estimates therefore reflect outcome-specific evidence rather than an artificial composite measure of “radiation toxicity.”

Vasculopathy represents perhaps the clearest long-term safety signal. The pooled incidence of 9.44% is compatible with, but less extreme than, some individual high-risk historical series. Grill et al. reported radiation-associated occlusive vasculopathy in 13 of 69 irradiated children and found a substantially greater frequency among patients with NF1 than among those without NF1 [9]. Kestle et al. similarly described post-radiation moyamoya phenomena, while Pierce et al. reported moyamoya in two irradiated patients with NF1 [21, 22]. The broader literature, however, demonstrates how strongly ascertainment can influence measured incidence. Tacke et al. systematically screened patients with magnetic resonance angiography and identified vascular abnormalities that would largely have remained clinically silent, whereas studies relying primarily on symptomatic events inevitably captured more advanced disease [11]. Picariello et al. likewise documented important cerebrovascular morbidity among young RT-exposed patients during prolonged follow-up, and Gan et al. identified post-radiotherapy moyamoya and stroke within a cohort characterized by substantial long-term hypothalamic and neuroendocrine morbidity [23, 24]. In contrast, Varan et al. reported only one case of moyamoya among 40 irradiated patients, illustrating the variation across cohorts [25]. Despite these differences, our estimate remained relatively stable after alternative handling of overlapping cohorts, exclusion of high-risk studies, and exclusion of the systematically screened MRA cohort, supporting vasculopathy as a reproducible late safety concern.

Secondary neoplasia constitutes another clinically important concern because many patients with OPHG survive for decades. Our broad pooled incidence of 5.41% was relatively robust to exclusion of high-risk and influential studies, whereas restriction to explicitly malignant secondary tumors reduced the estimate to 2.83%. This difference is meaningful because historical publications used heterogeneous terminology encompassing second gliomas, malignant nervous-system tumors, peripheral nerve sheath tumors, and other subsequent neoplasms. Jenkin et al. reported fatal second malignant tumors among irradiated patients and argued for increasingly selective use of irradiation in light of long-term morbidity [26]. Sharif et al. subsequently demonstrated a substantially higher burden of second nervous-system tumors in irradiated patients with NF1 than in their nonirradiated NF1 comparator group [10]. Lohkamp et al., in a more contemporary NF1 cohort, also documented a radiotherapy-induced secondary primitive neuroectodermal tumor together with post-radiation moyamoya, reinforcing the particular concern surrounding irradiation in genetically susceptible patients [27]. Conversely, Varan et al., Acharya et al., and Combs et al. reported no secondary malignancies during their respective follow-up periods [25, 28, 29], demonstrating both the rarity and latency-dependent nature of this outcome.

The exploratory comparison between irradiated and nonirradiated patients suggested greater odds of secondary neoplasia following radiotherapy, but this association cannot establish causality. Patients receiving irradiation commonly have more aggressive or treatment-refractory disease, may survive sufficiently long for delayed neoplasms to emerge, and often originate from older treatment eras with larger radiation fields and less conformal dose distributions. The NF1 comparison carries similar complexity because NF1 itself confers an intrinsic predisposition to multiple neoplasms. Sharif et al. emphasized precisely this problem: irradiation may add to an already elevated background tumor risk, and the longer latency observed for tumors in irradiated patients was compatible with radiation-associated carcinogenesis [10]. Accordingly, our separate analyses of broad second tumors, explicitly malignant neoplasms, and events specifically attributed to radiotherapy are more informative than treating every post-treatment neoplasm as radiation-induced.

Endocrine morbidity was the most frequent specific toxicity quantitatively identified. This is unsurprising given the intimate anatomical relationship among hypothalamic tumors, the hypothalamic–pituitary axis, and historical radiation fields. Earlier reports described considerable endocrine morbidity after irradiation. Horwich et al. documented delayed hypopituitarism appearing many years after treatment, while Pierce et al. observed a high prevalence of growth-hormone deficiency among long-term survivors [21, 30]. Tao et al. reported new hypopituitarism in 21 of 29 irradiated children, emphasizing the considerable endocrine burden detectable with sufficiently mature follow-up [31]. Grabenbauer et al. similarly demonstrated greater hypothalamic–pituitary dysfunction among younger patients and emphasized that long-term observation was essential for assessment of late morbidity [32]. More recent longitudinal evidence has reinforced the complexity of this outcome: Gan et al. demonstrated that hypothalamic–pituitary dysfunction evolves over decades and reflects both tumor location and treatment exposure, while Picariello et al. showed profound evolving neuroendocrine and metabolic morbidity among children diagnosed at very young ages [23, 24].

Our meta-regression provides an additional quantitative perspective: longer study-level follow-up was associated with a greater reported frequency of incident endocrinopathy. Although this is an ecological relationship rather than an individual time-to-event estimate, it illustrates one of the central problems in evaluating late radiation toxicity—**a treatment can appear safer simply because the cohort has not yet been followed long enough**. This issue is especially relevant when historical fractionated radiotherapy is compared with contemporary stereotactic techniques. In our exploratory analysis, conventional or unspecified fractionated RT was associated with a considerably higher pooled endocrinopathy estimate than stereotactic RT/SRS. Modern stereotactic techniques can substantially reduce dose to uninvolved hypothalamic–pituitary tissue, an advantage emphasized in the early fractionated stereotactic experience of Debus et al. and Combs et al. [28, 33]. However, modality, treatment era, age, selection, and follow-up duration were strongly confounded. The apparent difference therefore cannot establish that stereotactic treatment intrinsically carries a lower long-term endocrine risk.

The contemporary stereotactic literature illustrates this tension between encouraging early safety and incomplete late follow-up. Han et al. reported no new endocrine dysfunction, visual deterioration, intellectual deficit, or treatment-related mortality among four children treated with Novalis stereotactic irradiation, but acknowledged the very small cohort and need for longer observation [34]. Ge et al. reported Gamma Knife radiosurgery in 52 patients, with four transient conjunctival reactions and otherwise relatively limited acute toxicity; visual deterioration was observed in some patients but commonly coincided with tumor relapse and could not be confidently attributed to irradiation [35]. Zagirov et al. similarly reported no treatment-related adverse effects or deterioration in visual acuity after hypofractionated CyberKnife treatment in 16 patients [36]. El-Shehaby et al. observed no radiation-induced optic neuropathy or new endocrinopathy after single-session Gamma Knife radiosurgery, although transient tumor swelling produced reversible visual worsening in four patients [37]. These reports are encouraging, but none has the decades-long observation available for some historical cohorts, and therefore absence of late events should not yet be interpreted as proof of absence of late risk.

Visual outcomes require particularly rigorous attribution. OPHG itself damages the optic apparatus, and deterioration after treatment may reflect tumor progression, pre-existing severe visual loss, surgery, cystic change, edema, or radiation injury. Acharya et al. provided detailed longitudinal ophthalmologic assessment after definitive radiotherapy and found that visual-acuity decline occurred predominantly during the first two years, but these events were not equivalent to confirmed radiation optic neuropathy [29]. Awdeh et al., using prospective longitudinal visual assessment after conformal radiation therapy, similarly demonstrated variable post-treatment visual trajectories influenced by previous chemotherapy and surgery, without directly establishing irradiation as the cause of visual decline [38]. Bennebroek et al. reported deterioration among some irradiated children with isolated optic nerve glioma, but again attribution to radiation could not be established and the irradiated subgroup was very small [39]. These studies justify the conservative approach used in our meta-analysis: visual worsening was classified as treatment-related toxicity only when attribution was sufficiently explicit.

The historical literature further demonstrates why this distinction is essential. Gould et al. reported no definite complications that could confidently be attributed to radiotherapy, but explicitly acknowledged the difficulty of distinguishing treatment effects from NF1-related neurological and vascular abnormalities [40]. Montgomery et al. observed preserved or improved vision among surviving irradiated patients and no reported radiation-associated cataract formation, but systematic endocrine, vascular, neurocognitive, and oncological surveillance was not performed [41]. Packer et al. described intellectual abnormalities among irradiated survivors, yet several affected patients also had NF1 or received repeated irradiation, preventing separation of tumor, genetic, and treatment effects [42]. These older studies remain important because of their prolonged follow-up, but their toxicity ascertainment does not meet contemporary standards.

Radiation-related mortality was uncommon in our quantitative synthesis, whereas all-cause mortality was substantially higher. These endpoints should remain conceptually separate. Death following radiotherapy does not imply death caused by radiotherapy: progression, treatment-resistant tumor biology, NF1-associated disease, second tumors, and unrelated medical causes all contribute over long-term follow-up. Historical studies such as Montgomery, Horwich, Packer, Jenkin, and Gould span treatment eras in which both disease management and toxicity attribution differed considerably from contemporary practice [26, 30, 40-42]. Similarly, Gnekow et al. demonstrated the evolving strategy of using chemotherapy to defer irradiation in younger children, but their multicenter treatment study was not designed to systematically quantify late RT toxicity [1]. Liu et al. more recently compared postoperative treatment strategies in sporadic pediatric OPG, but RT-specific adverse events could not be separated from complications of surgery or other adjuvant therapy [43]. These studies illustrate a broader finding of our review: safety evidence is often present in the literature, but not in a form permitting valid radiation-specific quantitative extraction.

This distinction is also important for the interpretation of radiation necrosis, severe toxicity, and neurocognitive injury. No radiation necrosis was identified among the patients with sufficiently extractable data, while grade ≥3 toxicity and explicitly treatment-attributed neurocognitive toxicity were supported by only a handful of observations. These results indicate **sparse reported events**, not demonstrated absence of risk. Neurocognitive morbidity is especially vulnerable to confounding. Older cohorts described learning difficulties, intellectual impairment, and educational support requirements, but many patients had baseline hypothalamic involvement, hydrocephalus, previous surgery, NF1, or neurological deficits before radiation [21, 31, 42]. Without pretreatment neuropsychological testing and longitudinal standardized assessment, attributing later cognitive dysfunction specifically to radiation is inherently uncertain.

Clinically, our findings argue against treating radiotherapy toxicity as a single uniform entity. Risk must be individualized according to patient age, NF1 status, tumor anatomy, existing visual and endocrine deficits, anticipated life expectancy, prior systemic therapy and surgery, radiation dose distribution, and available alternatives. The vascular and oncological concerns documented in NF1-associated cohorts support continued caution when irradiation is contemplated for NF1-associated OPHG. This is consistent with the historical evolution from radiation toward observation and systemic therapy described in cohorts such as Gnekow et al. and Lohkamp et al. [1, 27]. However, these findings should not be interpreted as evidence that radiotherapy should invariably be avoided. Progressive OPHG can itself cause irreversible blindness, endocrine dysfunction, hypothalamic injury, and neurological morbidity. The relevant clinical decision is therefore whether the probability of durable disease and visual-pathway control outweighs the individualized probability and consequence of delayed toxicity.

Treatment era must also be considered when applying these results to present practice. Early series used orthovoltage, cobalt, and large-field conventional photon techniques, whereas contemporary patients may receive highly conformal photon treatment, intensity-modulated radiotherapy, proton therapy, fractionated stereotactic radiotherapy, or carefully selected radiosurgery. Tsang et al. incorporated modern photon and proton approaches within a large pediatric radiation cohort, and Hanania et al. provided contemporary proton-era safety information, whereas Debus and Combs demonstrated the potential of stereotactic fractionation to reduce normal-tissue exposure [7, 8, 28, 33]. Modern conformality should logically reduce unnecessary dose to the pituitary, hypothalamus, cerebral vasculature, temporal structures, and uninvolved brain. Nevertheless, the longest follow-up belongs disproportionately to patients treated with older techniques. The apparent safety of contemporary treatment is therefore partly an issue of **follow-up maturity**, and late vasculopathy, endocrinopathy, or secondary neoplasia may not yet have had sufficient time to manifest.

A further contribution of this review is the demonstration that the evidence base itself is structurally uneven. The integrated topology showed dense shared evidence around vasculopathy, secondary neoplasms, endocrinopathy, and mortality, while severe toxicity, radiation necrosis, and neurocognitive toxicity were represented by very few studies. This explains why some of the most clinically concerning long-term outcomes also have the least precise estimates. Rather than pooling incompatible observations or interpreting non-reporting as absence of events, we retained sparse endpoints descriptively. This approach reduces apparent statistical completeness but provides a more defensible representation of the available evidence.

### Future directions

Future research should prioritize **prospective, multicenter, longitudinal safety cohorts** rather than additional small retrospective series. Standardized definitions are needed for incident endocrinopathy, radiation-associated vasculopathy, radiation optic neuropathy, neurocognitive decline, radiation necrosis, severe toxicity, and subsequent neoplasms. Baseline abnormalities should be documented before treatment and each adverse event should be reported with an explicit numerator, denominator, latency, severity, reversibility, and attribution. This is particularly important in OPHG because the disease itself can produce many of the same endocrine, vascular, neurological, and visual abnormalities attributed to treatment.

Future studies should also incorporate detailed dosimetry. Dose–volume exposure of the optic nerves and chiasm, hypothalamus, pituitary, internal carotid and cerebral arteries, hippocampi, temporal structures, and other organs at risk should be related prospectively to long-term toxicity. Modern photon therapy, proton therapy, fractionated stereotactic treatment, and radiosurgery should be compared using common outcome definitions and sufficiently mature follow-up. Randomized trials are unlikely to be feasible for many of these questions given disease rarity, but prospective registries, multinational individual-participant-data collaborations, propensity-adjusted comparative cohorts, and harmonized survivorship studies could markedly improve inference.

NF1 should be treated as a prespecified biological risk stratum rather than simply a demographic covariate. Future studies should investigate interactions among NF1 status, age at irradiation, radiation dose to large cerebral vessels, treatment technique, and subsequent vascular or oncological events. Lifelong follow-up is likely to be particularly important for second neoplasms because latency may extend well beyond the duration of most contemporary radiation series. Similarly, standardized longitudinal MRA, endocrine testing, neuropsychological assessment, and formal ophthalmological examinations would help distinguish true treatment effects from differences in surveillance intensity.

Finally, a dedicated **core outcome set for OPHG radiation safety** would substantially improve future evidence synthesis. Such a framework should incorporate vascular, endocrine, ophthalmological, neurocognitive, neurological, oncological, severe-toxicity, and mortality outcomes together with functional and patient-reported outcomes. Future investigations should move beyond simply recording whether an adverse event occurred and instead characterize its latency, severity, persistence, reversibility, functional consequences, and relationship to radiation dose. This would transform the field from heterogeneous retrospective toxicity reporting toward genuinely individualized risk prediction.

### Limitations

This study must be interpreted in the context of several limitations. The available evidence was overwhelmingly non-randomized and predominantly retrospective, leaving unavoidable risks of selection bias, confounding by indication, incomplete follow-up, and selective toxicity ascertainment. Studies extended from the 1970s to the contemporary era and therefore combined markedly different radiation technologies, systemic therapies, imaging methods, endocrine surveillance, ophthalmological testing, and survivorship practices. Definitions and attribution were inconsistent: endocrine dysfunction may have preceded irradiation because of hypothalamic disease; visual deterioration could reflect tumor progression rather than radiation injury; vasculopathy was detected using methods ranging from symptom-triggered evaluation to systematic MRA; and subsequent tumors were not uniformly classified as benign, malignant, or radiation-induced. Many late toxicities were reported as crude event proportions rather than person-time incidence or competing-risk estimates, despite profound variation in follow-up duration. Comparative NF1-versus-sporadic and RT-versus-non-RT analyses were observational and could not fully control for age, disease severity, treatment era, radiation dose, prior therapy, follow-up, or surveillance intensity and therefore should not be interpreted causally. Several clinically important outcomes were also extremely sparse, producing wide confidence and prediction intervals; absence of reported radiation necrosis, permanent visual toxicity, or neurocognitive toxicity cannot be equated with absence of risk. Although 31 of the 35 studies supplied a usable event/denominator pair for at least one quantitative safety endpoint, no single meta-analysis incorporated all included studies because outcomes were inconsistently reported and some cohorts overlapped. Finally, the evidence-topology analysis was exploratory and describes similarity in **study-level outcome coverage**, not patient-level co-occurrence, biological relationships among toxicities, or a network meta-analysis.

## Conclusion

Radiotherapy and radiosurgery remain important local treatment options for appropriately selected patients with optic pathway–hypothalamic glioma, but their long-term safety profile is heterogeneous and incompletely characterized. Incident endocrinopathy and vasculopathy were the most prominent quantifiable late toxicities, while secondary neoplasms constituted an important long-term concern. Genetically susceptible populations, including patients with NF1, remain an important priority for long-term study. In contrast, permanent treatment-attributed visual toxicity, radiation necrosis, severe toxicity, and radiation-related mortality were infrequently documented, but these findings were based on substantially sparser evidence and should not be interpreted as proof of negligible risk.

The clinical implication is therefore not that radiotherapy is uniformly safe or unsafe, but that its use should be **individualized according to disease threat, age, NF1 status, expected survival, prior therapy, treatment technique, and the long-term consequences most relevant to each patient**. Modern conformal and stereotactic techniques offer the potential to reduce unnecessary normal-tissue exposure, but their true late safety advantage requires much longer observation. Lifelong vascular, endocrine, visual, and oncological surveillance remains particularly important for irradiated survivors. Prospective multicenter cohorts with standardized toxicity definitions, detailed dosimetry, structured surveillance, and long-term follow-up are now needed to determine which patients can obtain durable benefit from radiotherapy while minimizing its lifelong cost.

## Supplementary Figure Legends

**Supplementary Figure S1.**
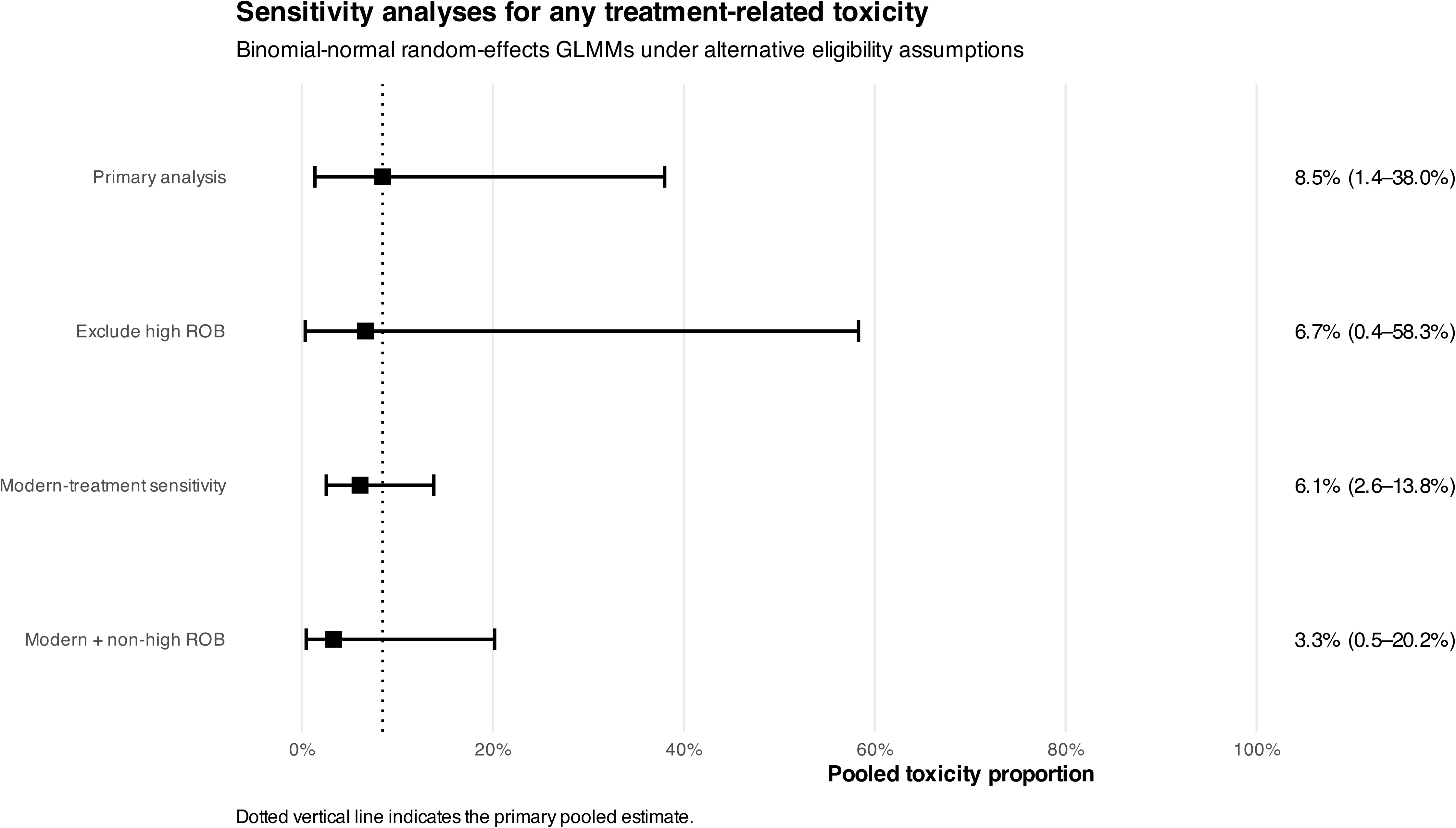
Sensitivity analyses for any treatment-related toxicity. Comparison of the primary random-effects GLMM estimate with analyses excluding studies at high risk of bias, restricting the evidence base to contemporary treatment series, and simultaneously restricting the analysis to contemporary and non-high-risk studies. The pooled estimates were 8.5% in the primary analysis, 6.7% after exclusion of high-risk studies, 6.1% in the modern-treatment sensitivity analysis, and 3.3% when both restrictions were applied.

**Supplementary Figure S2.**
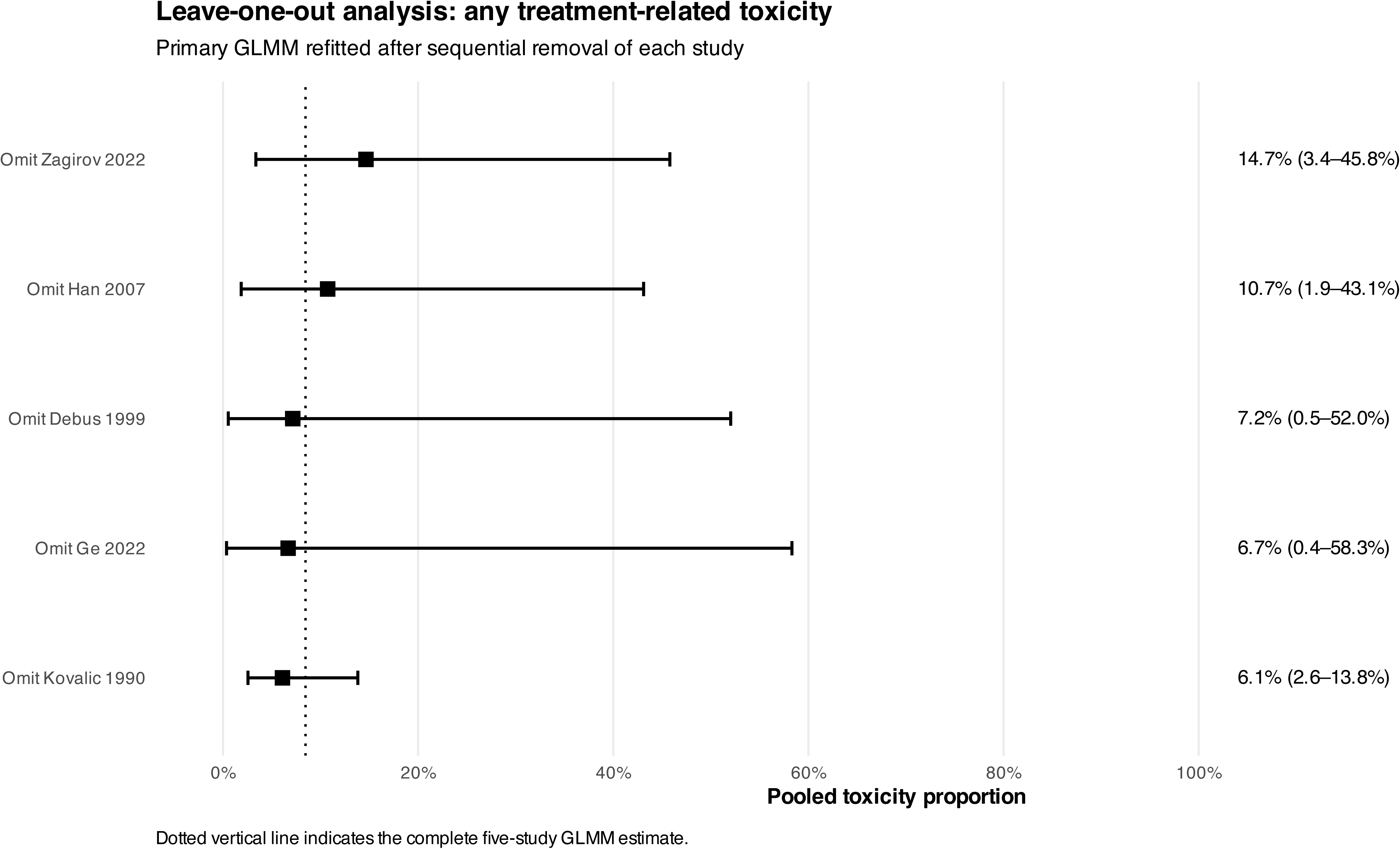
Leave-one-out analysis of any treatment-related toxicity. The random-effects binomial-normal GLMM was sequentially refitted after exclusion of each contributing study. The complete-analysis pooled estimate is displayed as a reference line to assess whether the primary result was disproportionately influenced by an individual study.

**Supplementary Figure S3.**
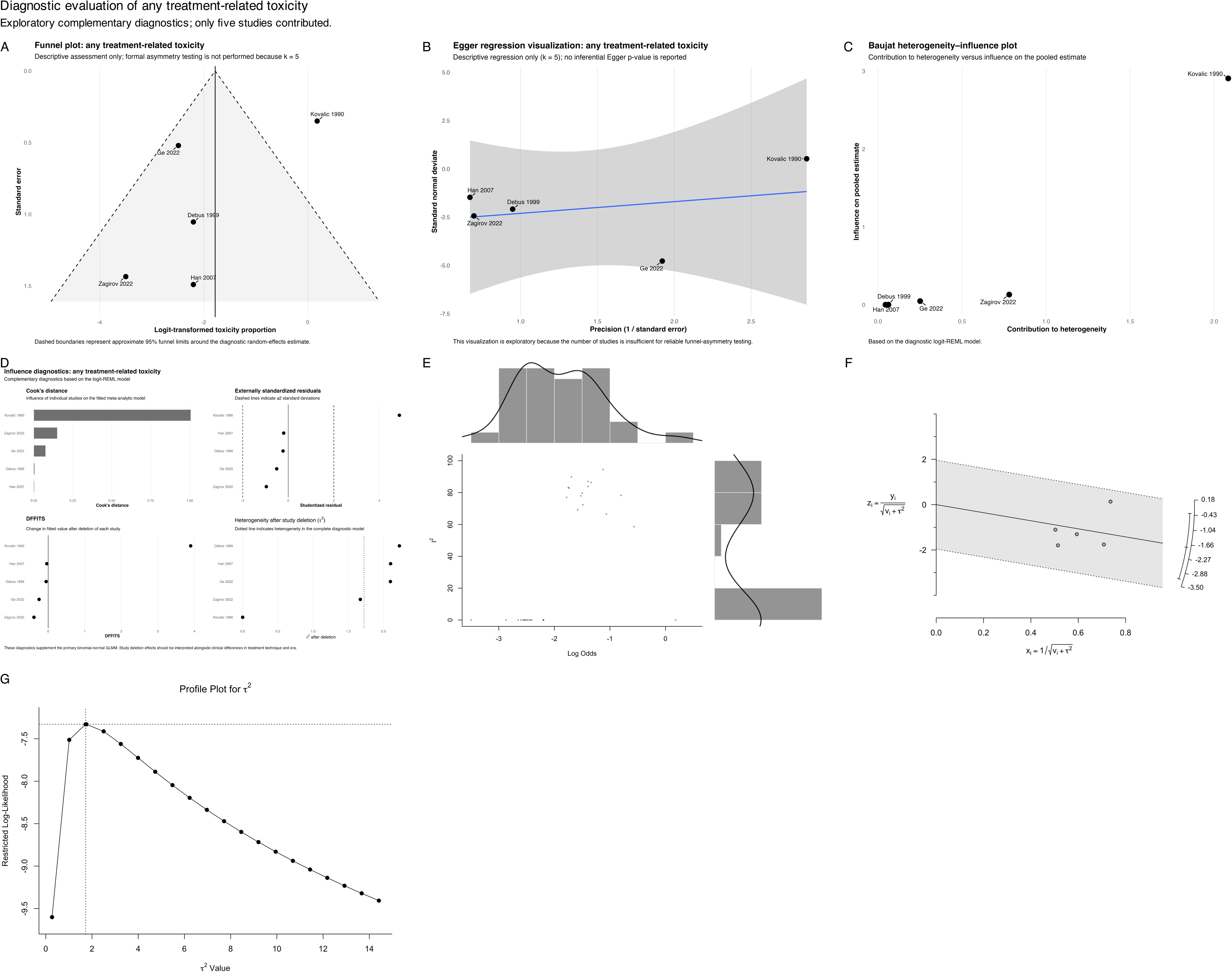
Diagnostic evaluation of the any-treatment-related-toxicity meta-analysis. Multipanel presentation of the funnel plot, regression-based asymmetry assessment, Baujat plot, influence diagnostics, graphical display of study heterogeneity, radial plot, and profile likelihood for between-study variance (τ^2^). Because only five studies contributed to the primary analysis, small-study-effect and asymmetry assessments are exploratory and should not be interpreted as formal evidence for or against publication bias.

**Supplementary Figure S4.**
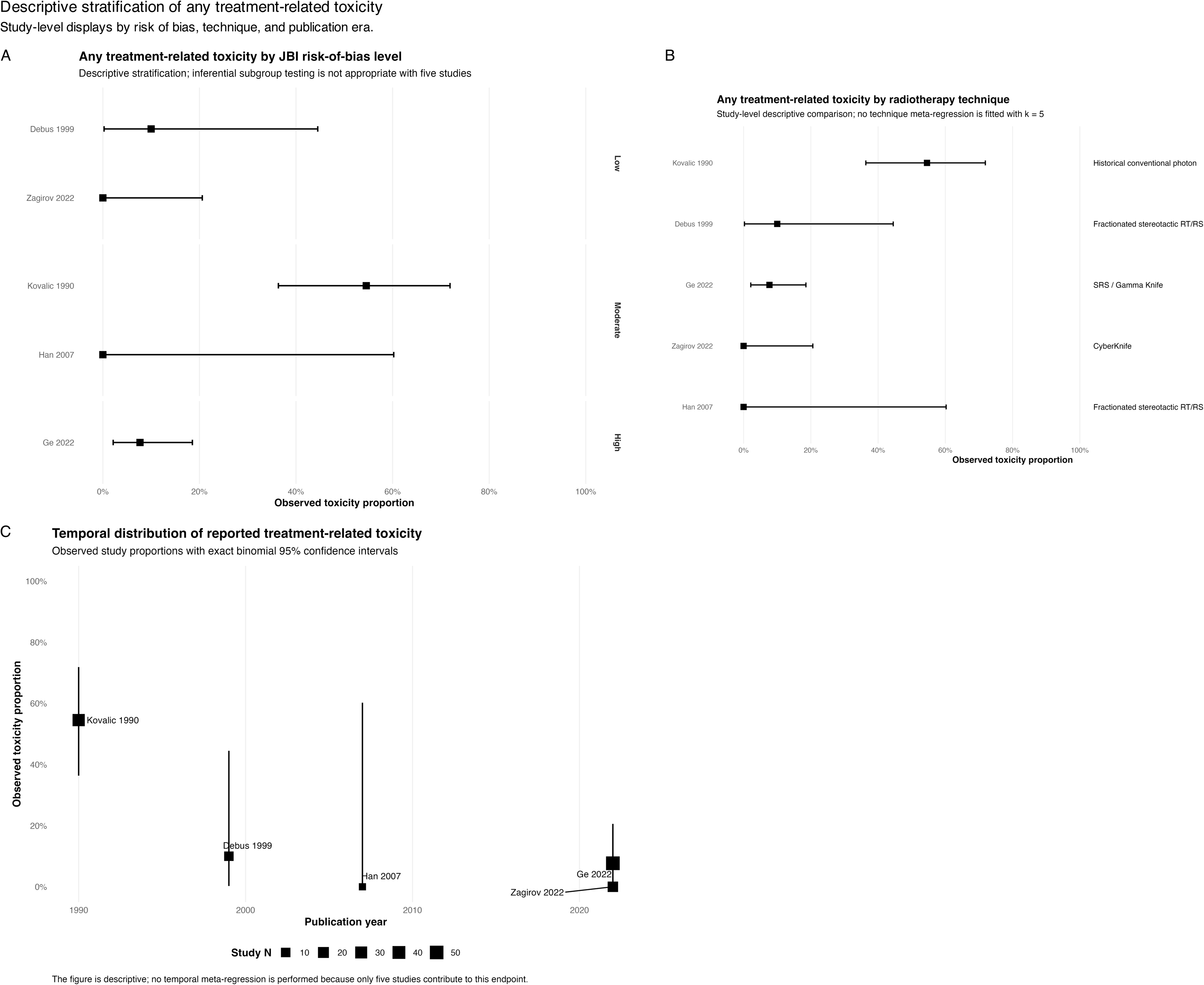
Descriptive stratification of any treatment-related toxicity. Observed study-level toxicity proportions stratified according to JBI risk-of-bias classification, radiotherapy/radiosurgery technique, and publication era. These analyses are descriptive because of the small number of studies contributing to the primary outcome.

**Supplementary Figure S5.**
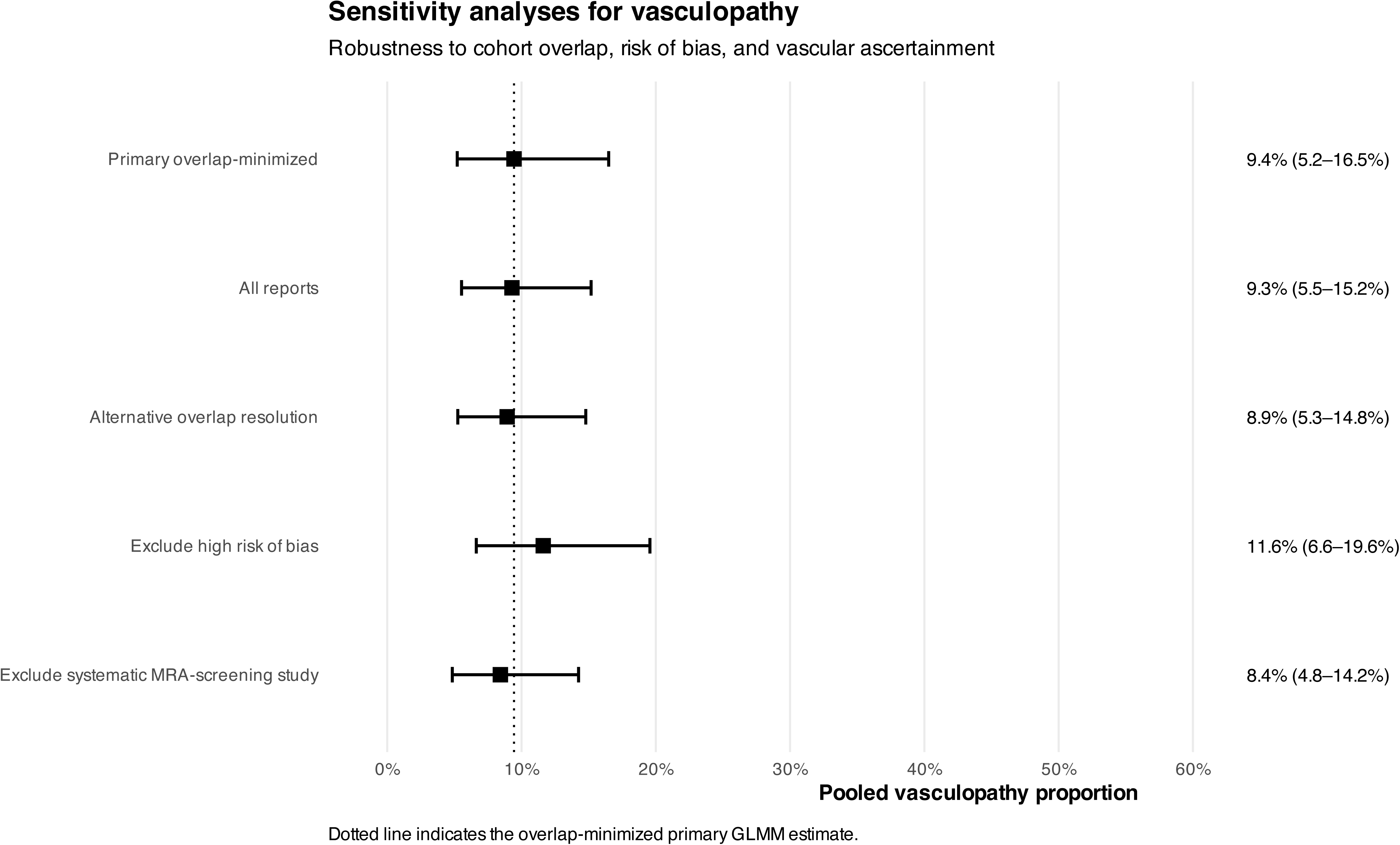
Sensitivity analyses for vasculopathy following radiotherapy or radiosurgery. The overlap-minimized primary estimate is compared with analyses including all eligible reports, using an alternative resolution of potentially overlapping cohorts, excluding studies at high risk of bias, and excluding the study using systematic magnetic resonance angiographic screening.

**Supplementary Figure S6.**
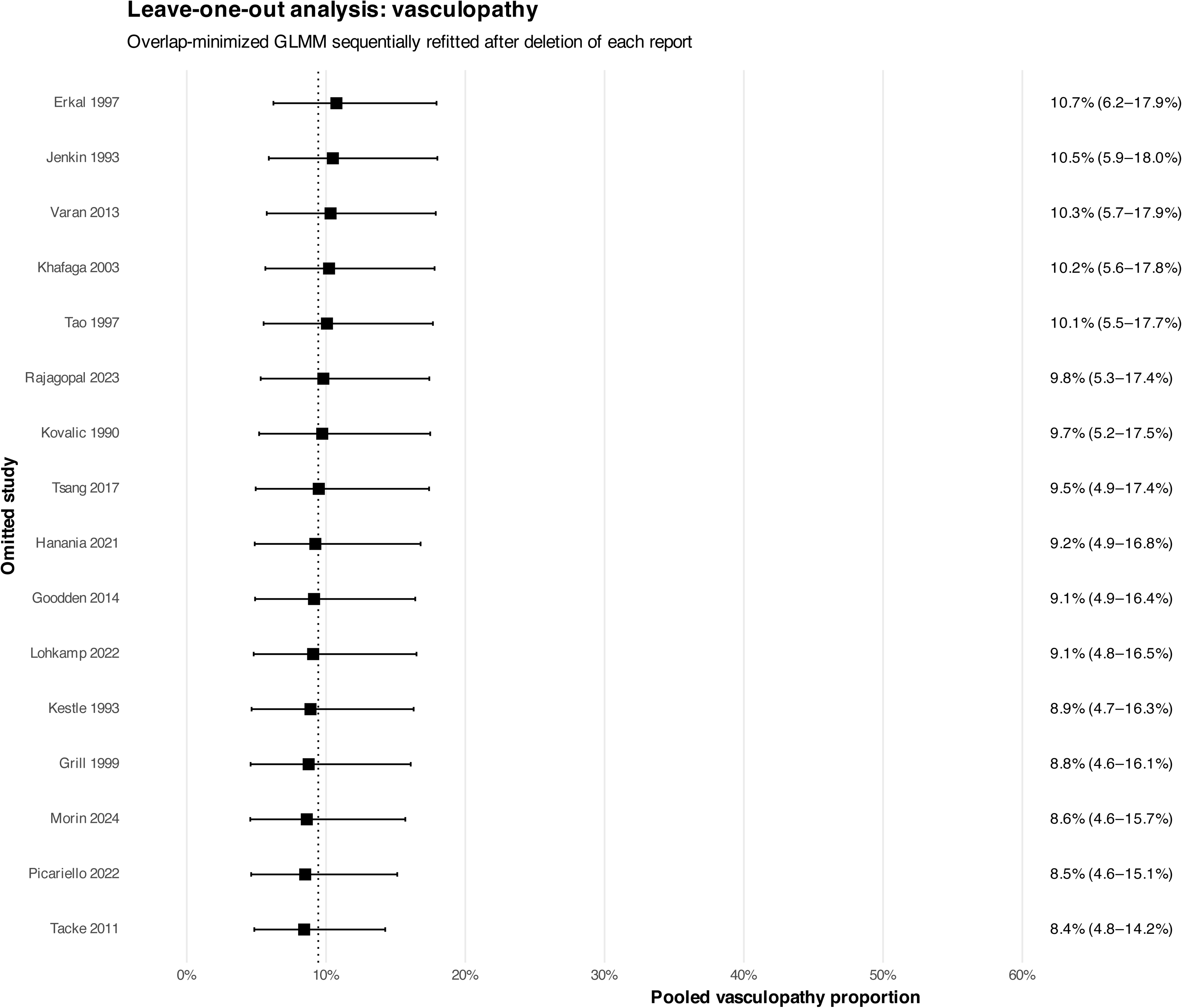
Leave-one-out analysis of vasculopathy. The overlap-minimized random-effects GLMM was sequentially refitted after removal of each individual study. The complete-analysis pooled estimate is displayed as a reference line to assess the influence of individual studies.

**Supplementary Figure S7.**
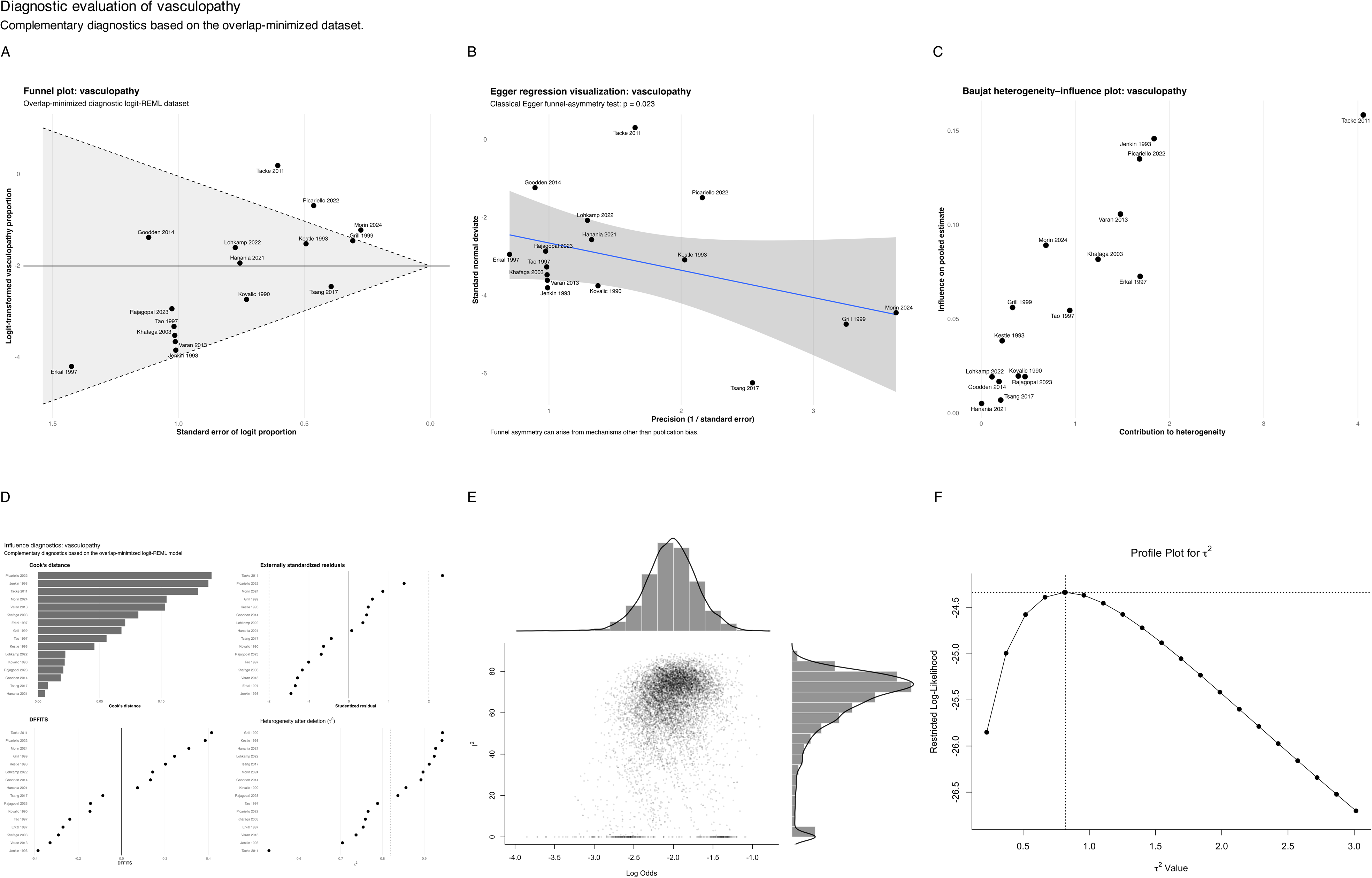
Diagnostic evaluation of the vasculopathy meta-analysis. Multipanel presentation of the funnel plot, regression-based asymmetry assessment, Baujat plot, influence diagnostics, graphical display of study heterogeneity, and profile likelihood for τ^2^. Regression-based asymmetry was detected (p=0.023), but cannot distinguish selective reporting from true clinical heterogeneity or differential vascular ascertainment.

**Supplementary Figure S8.**
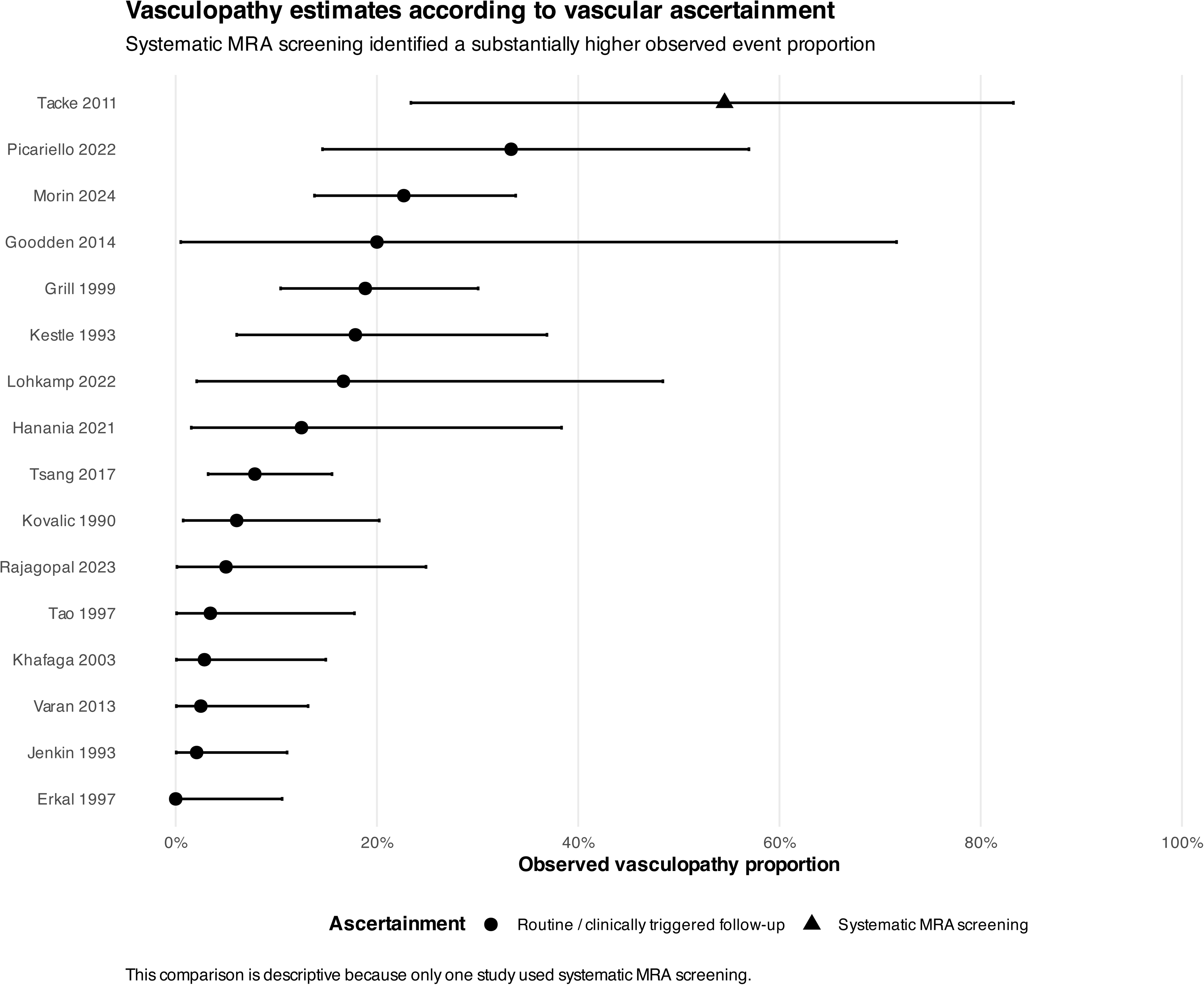
Vasculopathy according to method of vascular ascertainment. Observed vasculopathy proportions according to routine or clinically triggered surveillance versus systematic magnetic resonance angiographic screening. Systematic MRA screening identified a greater observed event proportion; however, only one contributing study used systematic MRA surveillance, and the comparison is therefore descriptive rather than inferential.

**Supplementary Figure S9.**
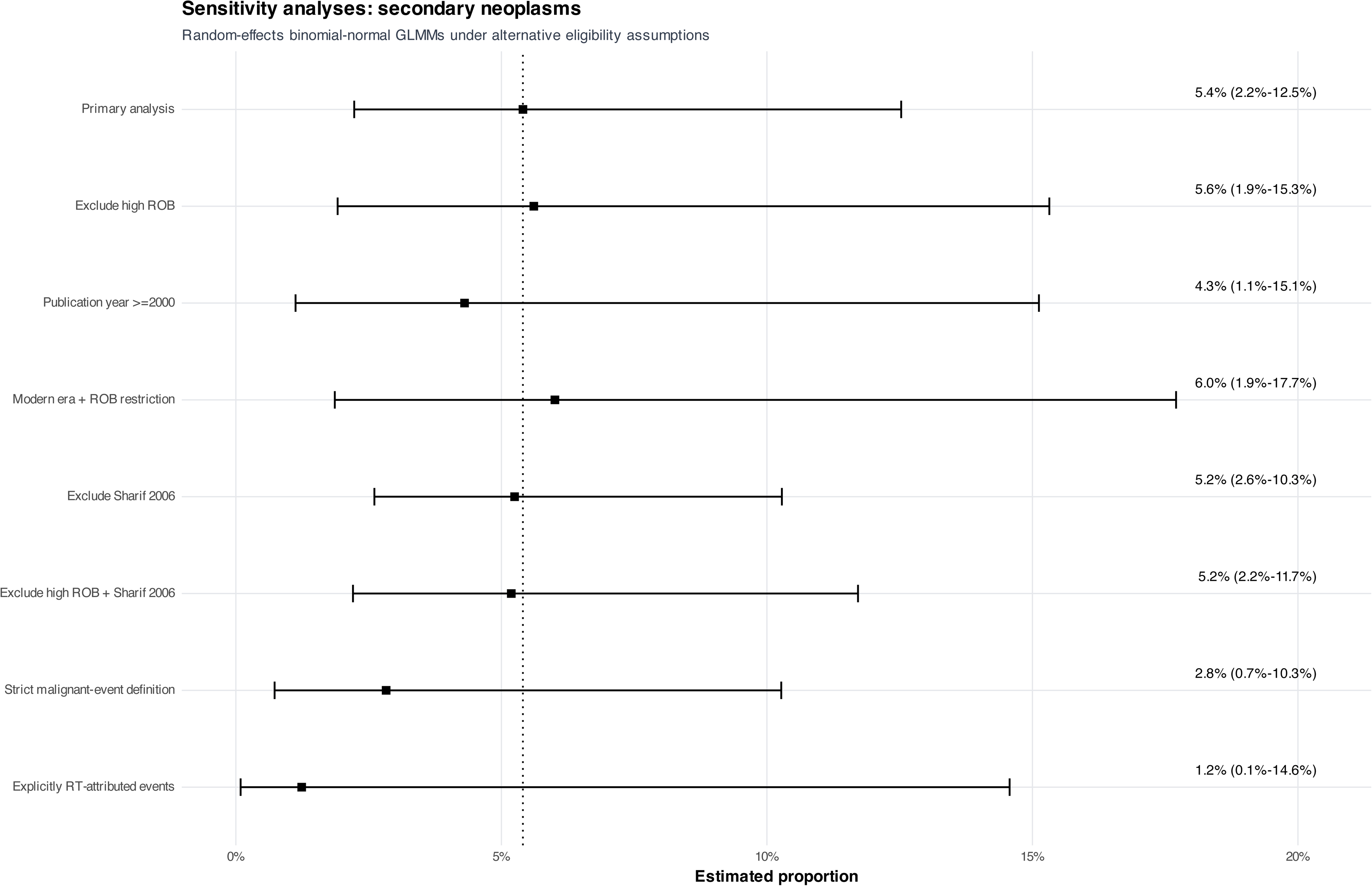
Sensitivity analyses for secondary neoplasms. Comparison of the primary random-effects GLMM with risk-of-bias, publication-era, combined modern-era/risk-of-bias, influential-study, strict malignant-event, and explicit radiotherapy-attribution restrictions. The broad pooled estimate was stable to study restrictions but decreased under narrower outcome definitions.

**Supplementary Figure S10.**
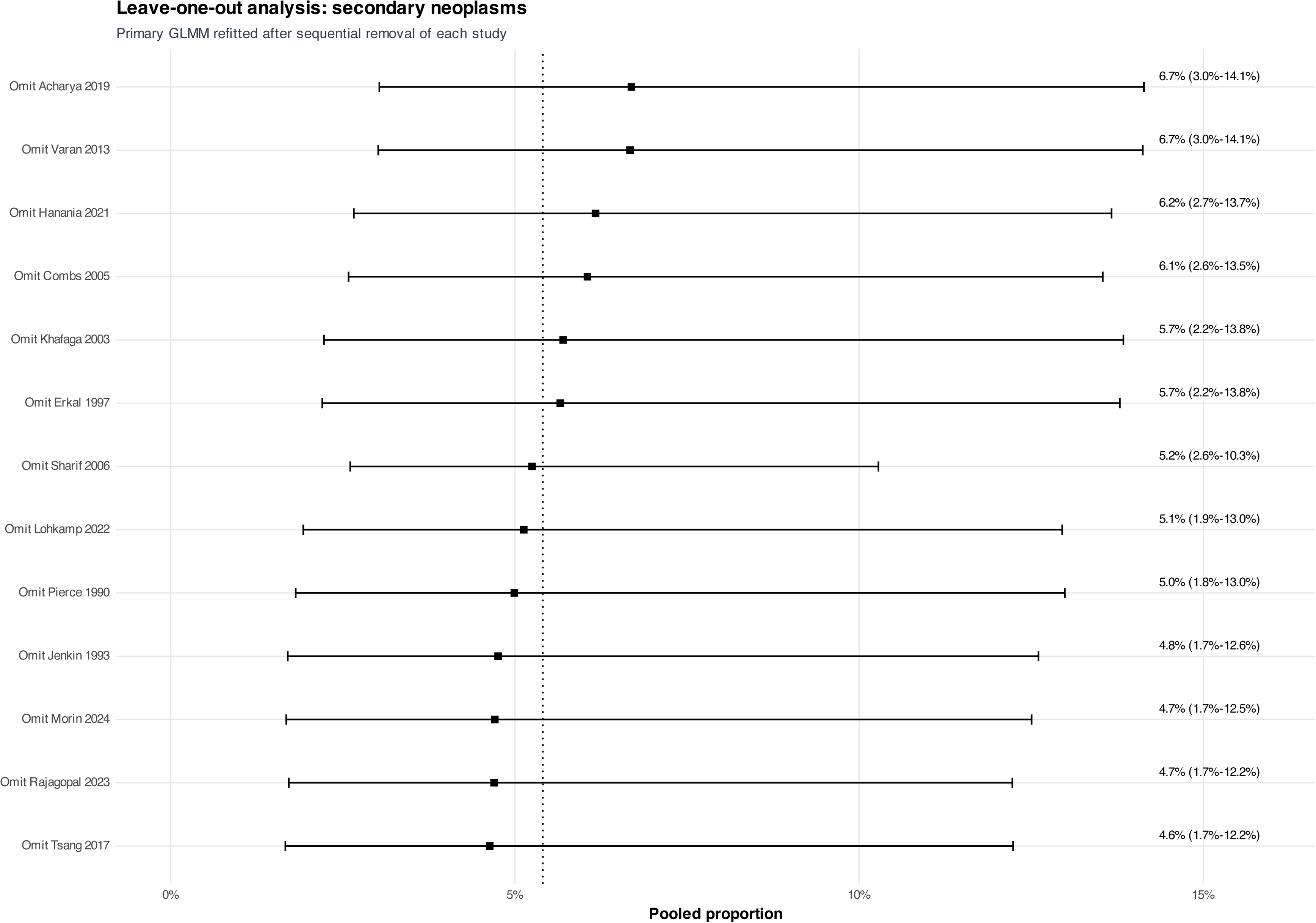
Leave-one-out analysis of secondary neoplasms. The primary GLMM was sequentially refitted after removal of each contributing study. Pooled estimates ranged from 4.6% to 6.7%, indicating that no single study fully determined the broad secondary-neoplasm estimate.

**Supplementary Figure S11.**
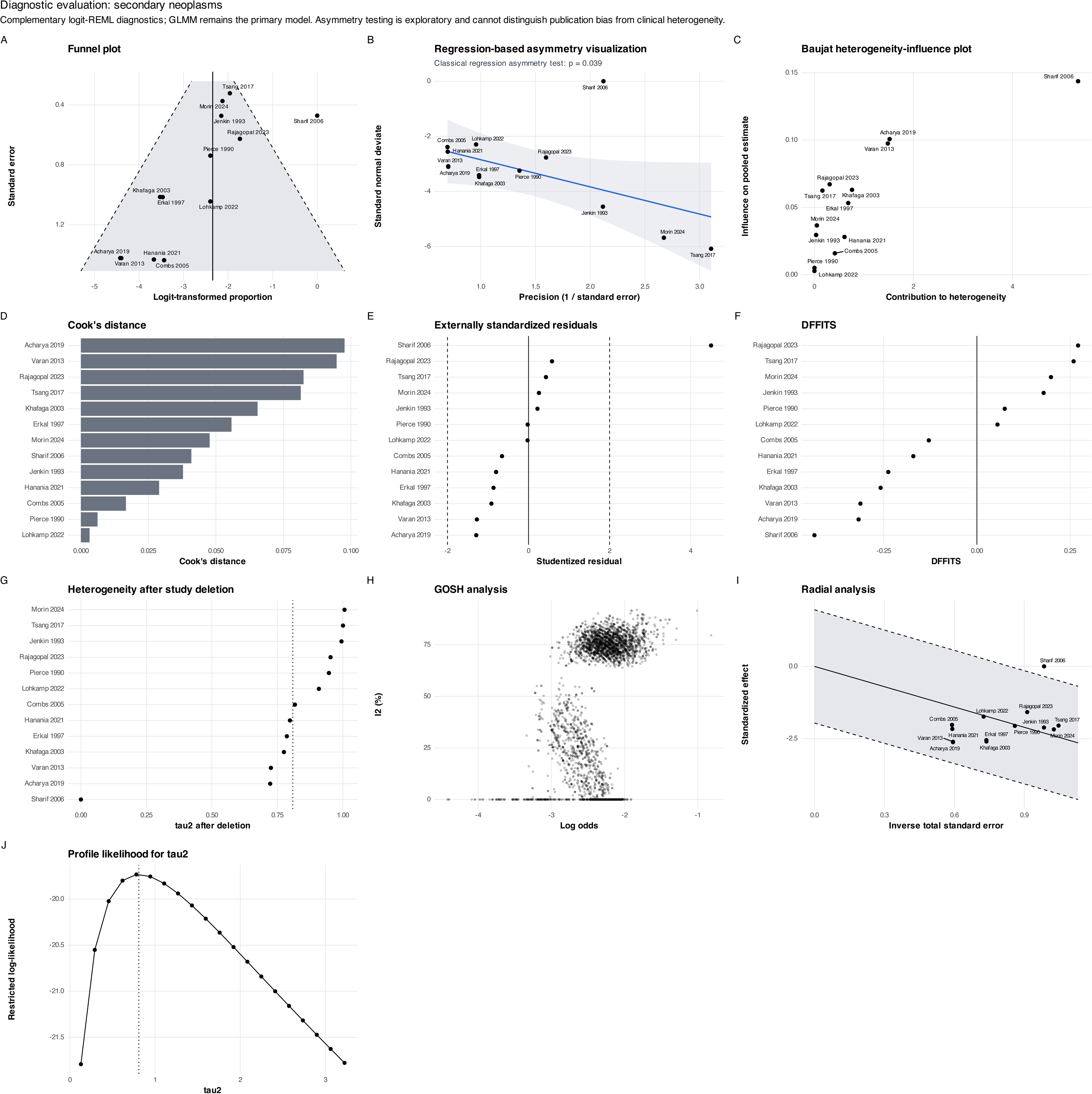
Diagnostic evaluation of the secondary-neoplasm meta-analysis. Funnel and regression-based asymmetry displays, Baujat plot, influence statistics, heterogeneity after study deletion, GOSH analysis, radial analysis, and profile likelihood for τ^2^. Regression-based asymmetry was detected (p=0.039), but cannot distinguish selective reporting from outcome-definition heterogeneity or sparse-event structure.

**Supplementary Figure S12.**
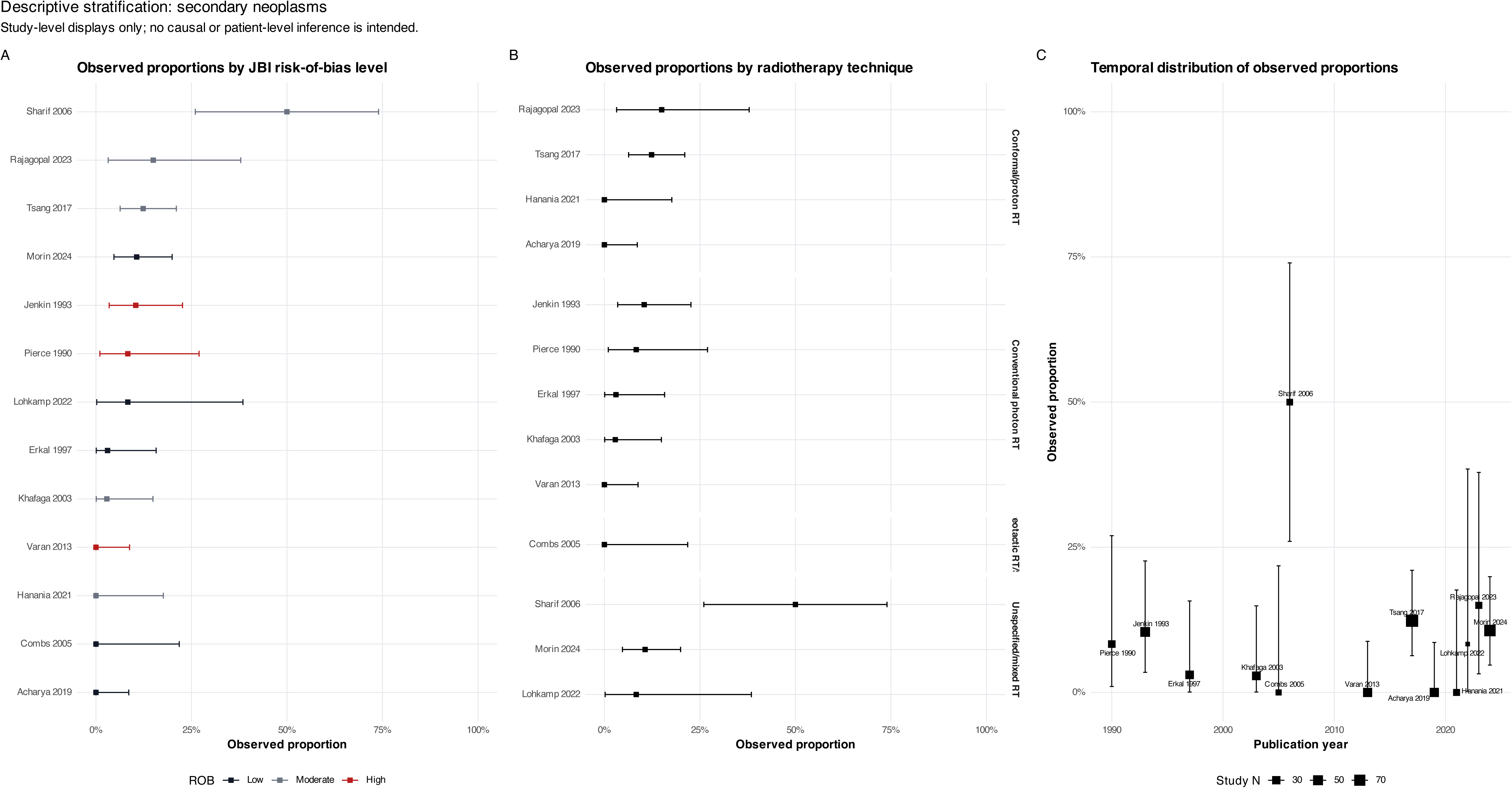
Descriptive stratification of secondary neoplasms. Observed study-level proportions displayed according to JBI risk-of-bias level, radiotherapy technique, and publication year. These plots are descriptive and do not imply causal or patient-level subgroup effects.

**Supplementary Figure S13.**
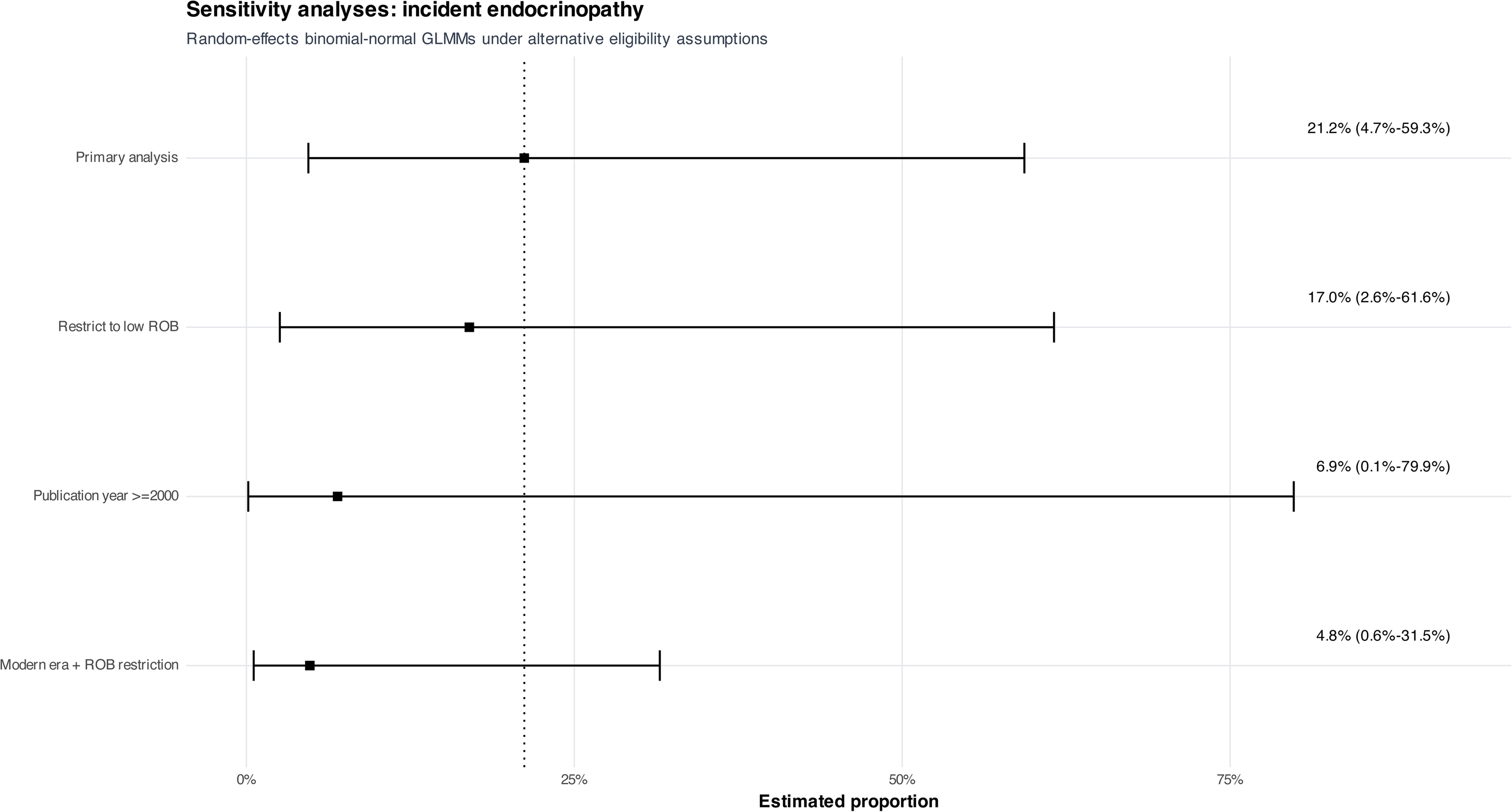
Sensitivity analyses for incident endocrinopathy. Comparison of the primary GLMM with low-risk-of-bias, publication-from-2000, and combined modern-era/risk-of-bias restrictions. Estimates decreased under contemporary restrictions but remained imprecise because few heterogeneous studies contributed.

**Supplementary Figure S14.**
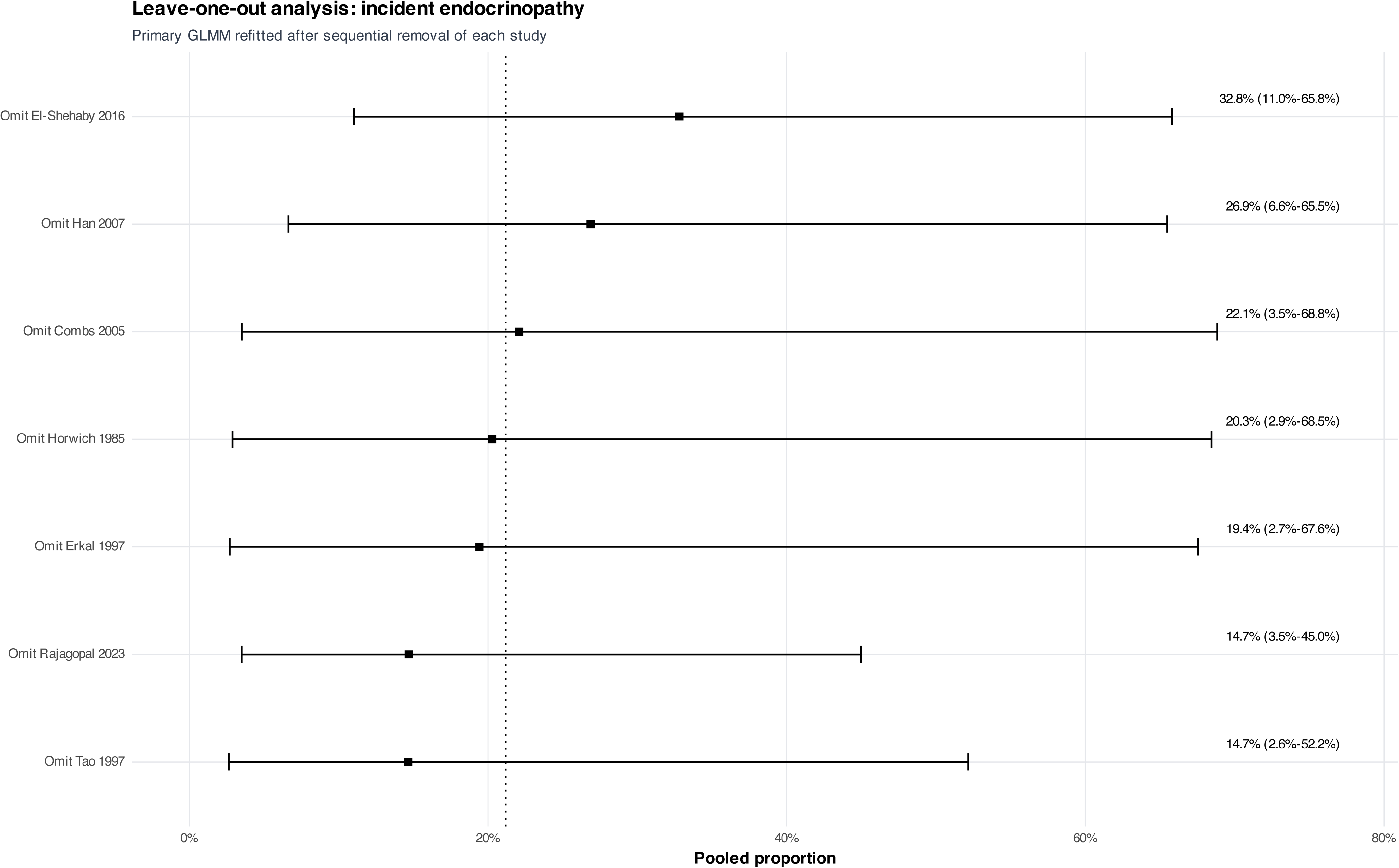
Leave-one-out analysis of incident endocrinopathy. Sequential study omission yielded pooled estimates from 14.7% to 32.8%, illustrating the influence of individual cohorts within a seven-study evidence base.

**Supplementary Figure S15.**
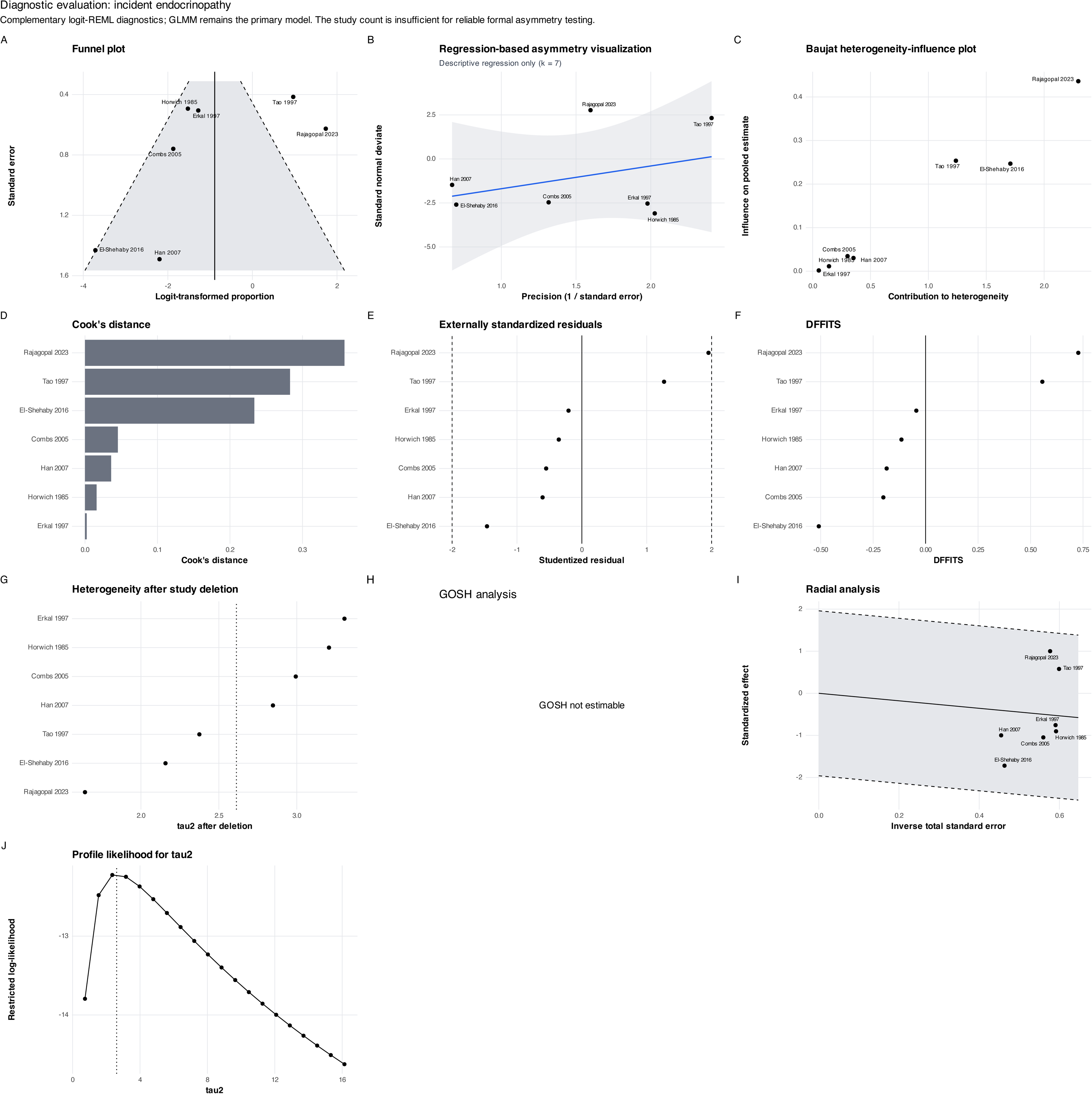
Diagnostic evaluation of the incident-endocrinopathy meta-analysis. Funnel and regression displays, Baujat plot, influence statistics, heterogeneity after deletion, radial analysis, and profile likelihood for τ^2^. GOSH analysis was not estimable, and asymmetry assessment was descriptive because only seven studies contributed.

**Supplementary Figure S16.**
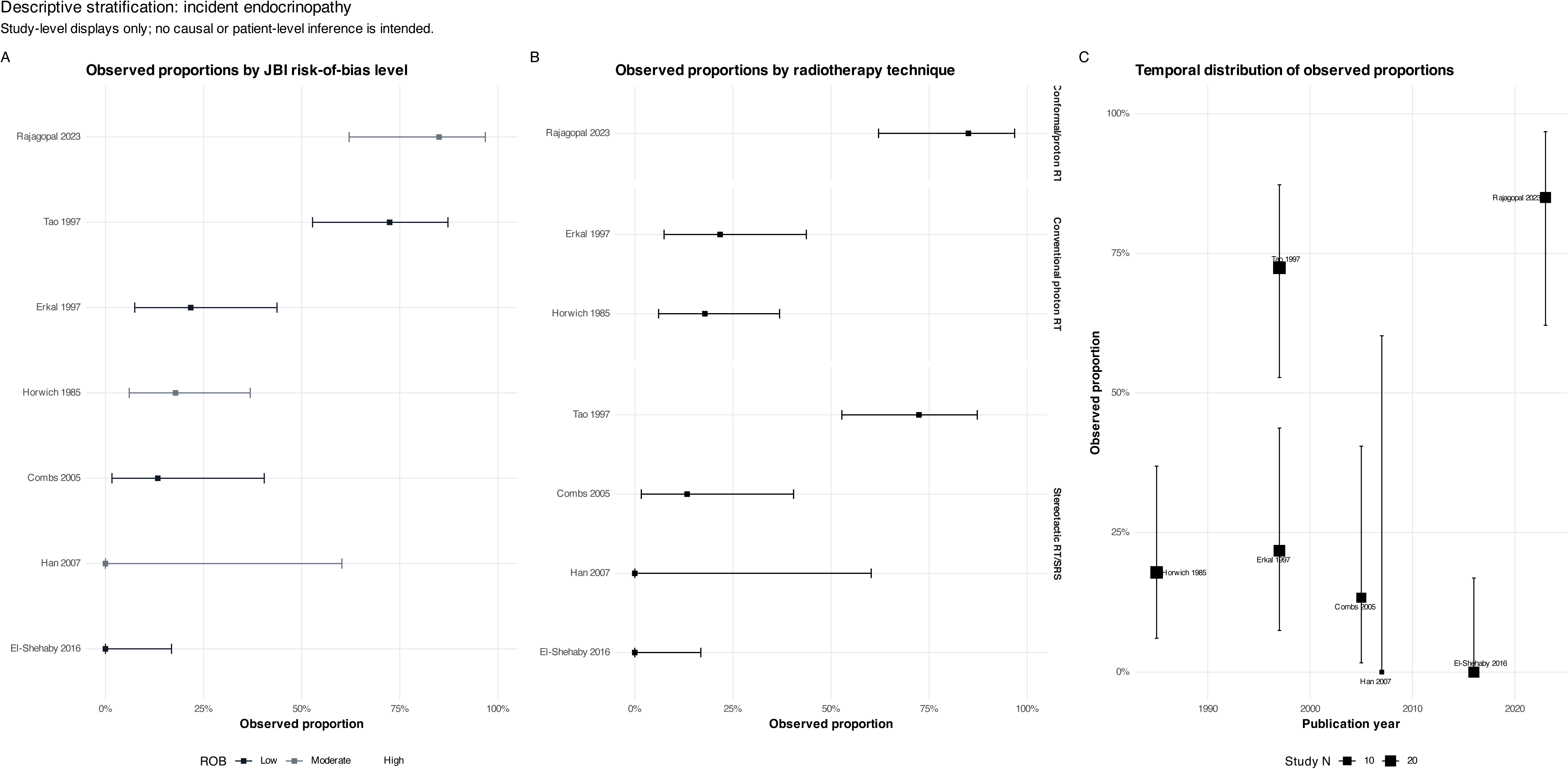
Descriptive stratification of incident endocrinopathy. Observed study-level proportions displayed according to JBI risk-of-bias level, radiotherapy technique, and publication year. These plots supplement, but do not replace, the exploratory modality and follow-up analyses reported in the main text.

**Supplementary Figure S17.**
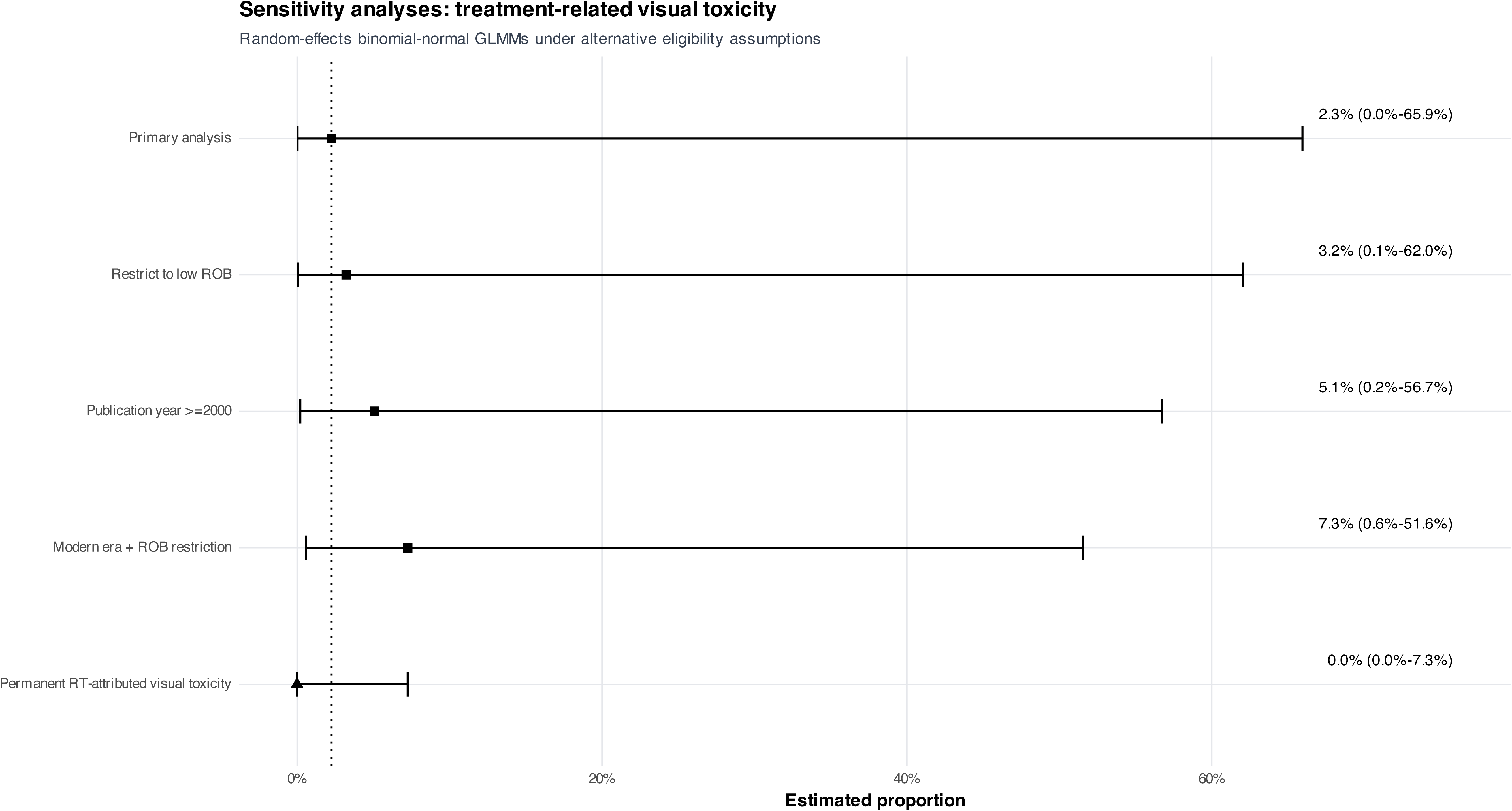
Sensitivity analyses for treatment-related visual toxicity. Comparison of the primary GLMM with low-risk-of-bias, publication-from-2000, combined restriction, and permanent-treatment-attributed event definitions. No permanent treatment-attributed visual toxicity was identified (0/49; exact 95% CI, 0–7.25%).

**Supplementary Figure S18.**
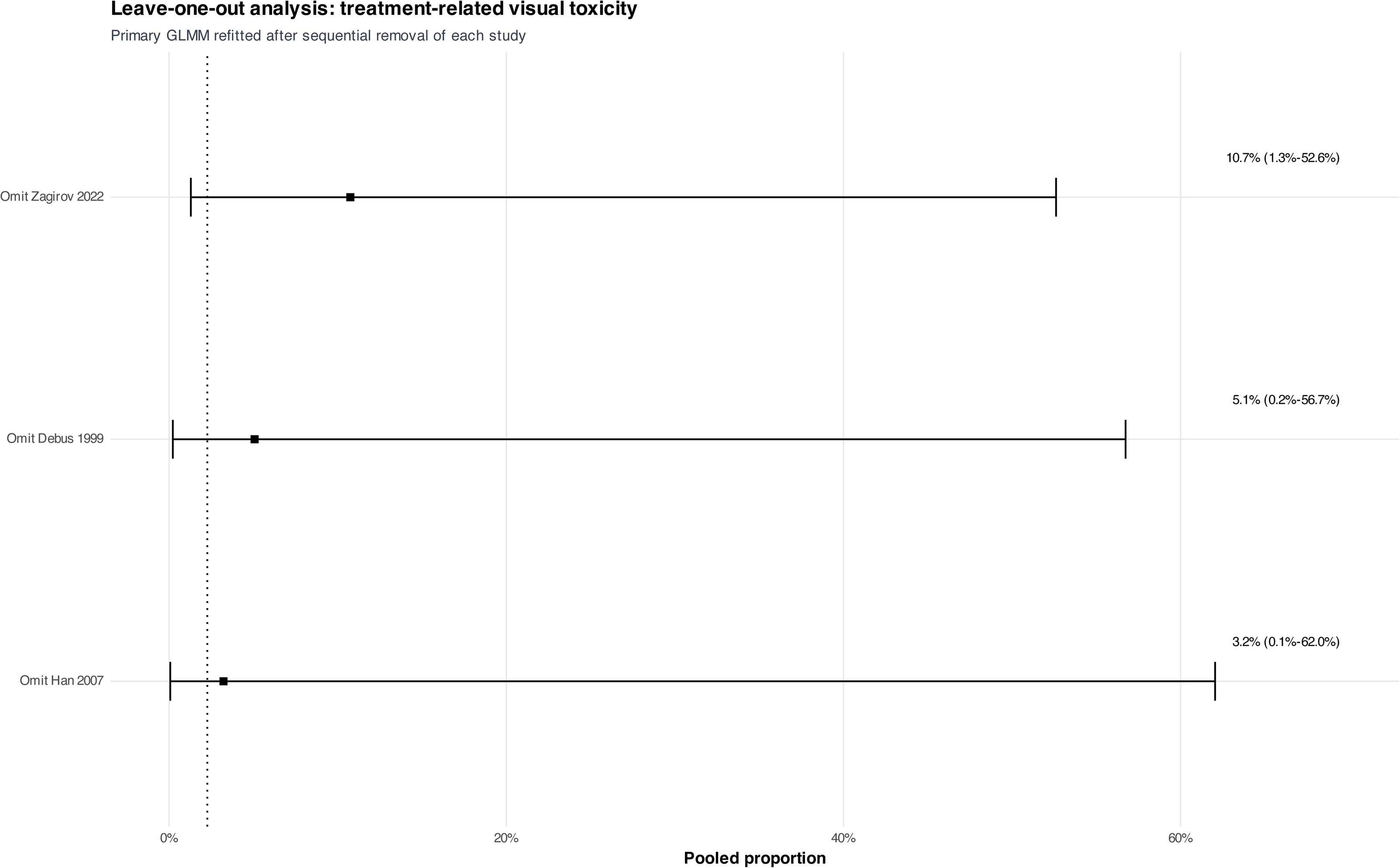
Leave-one-out analysis of treatment-related visual toxicity. The four-study GLMM was sequentially refitted after each study was removed. Estimates remained highly imprecise and ranged from 3.2% to 10.7%.

**Supplementary Figure S19.**
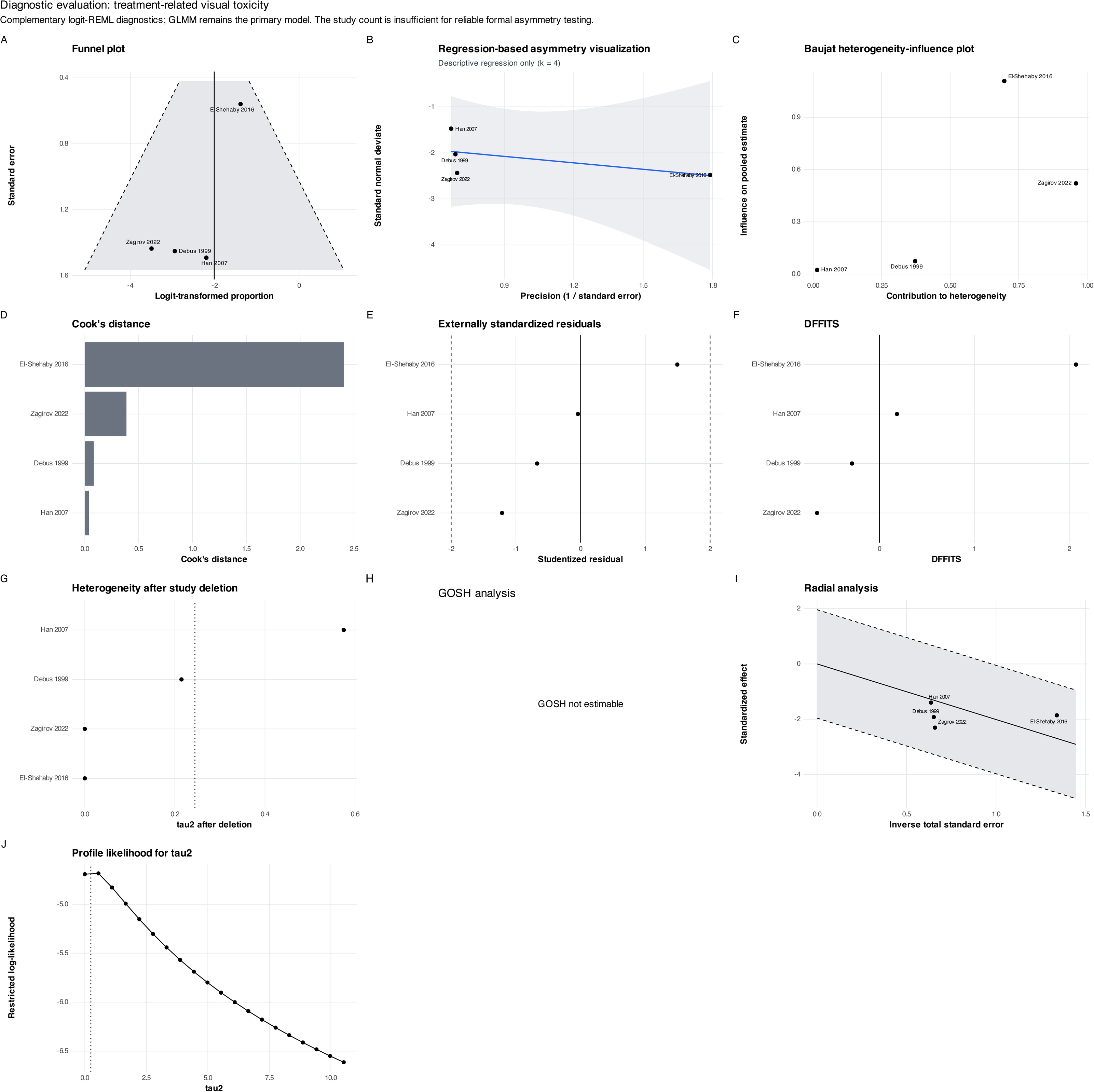
Diagnostic evaluation of the visual-toxicity meta-analysis. Funnel and regression displays, Baujat plot, influence statistics, heterogeneity after deletion, radial analysis, and profile likelihood for τ^2^. GOSH analysis was not estimable, and no formal asymmetry test was performed because only four studies contributed.

**Supplementary Figure S20.**
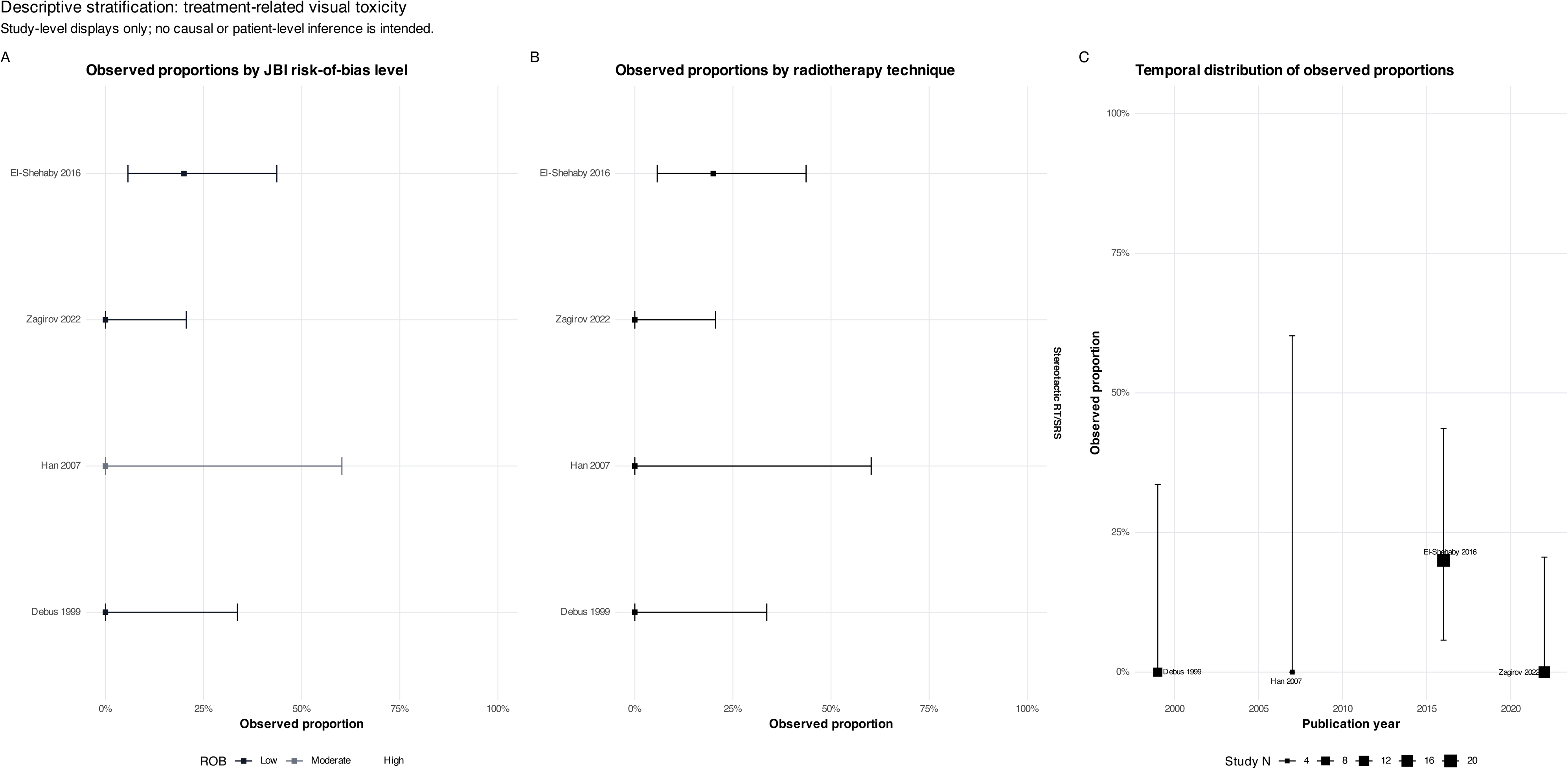
Descriptive stratification of treatment-related visual toxicity. Observed study-level proportions displayed according to JBI risk-of-bias level, radiotherapy technique, and publication year. These analyses are descriptive because all four attributable events occurred in one study.

**Supplementary Figure S21.**
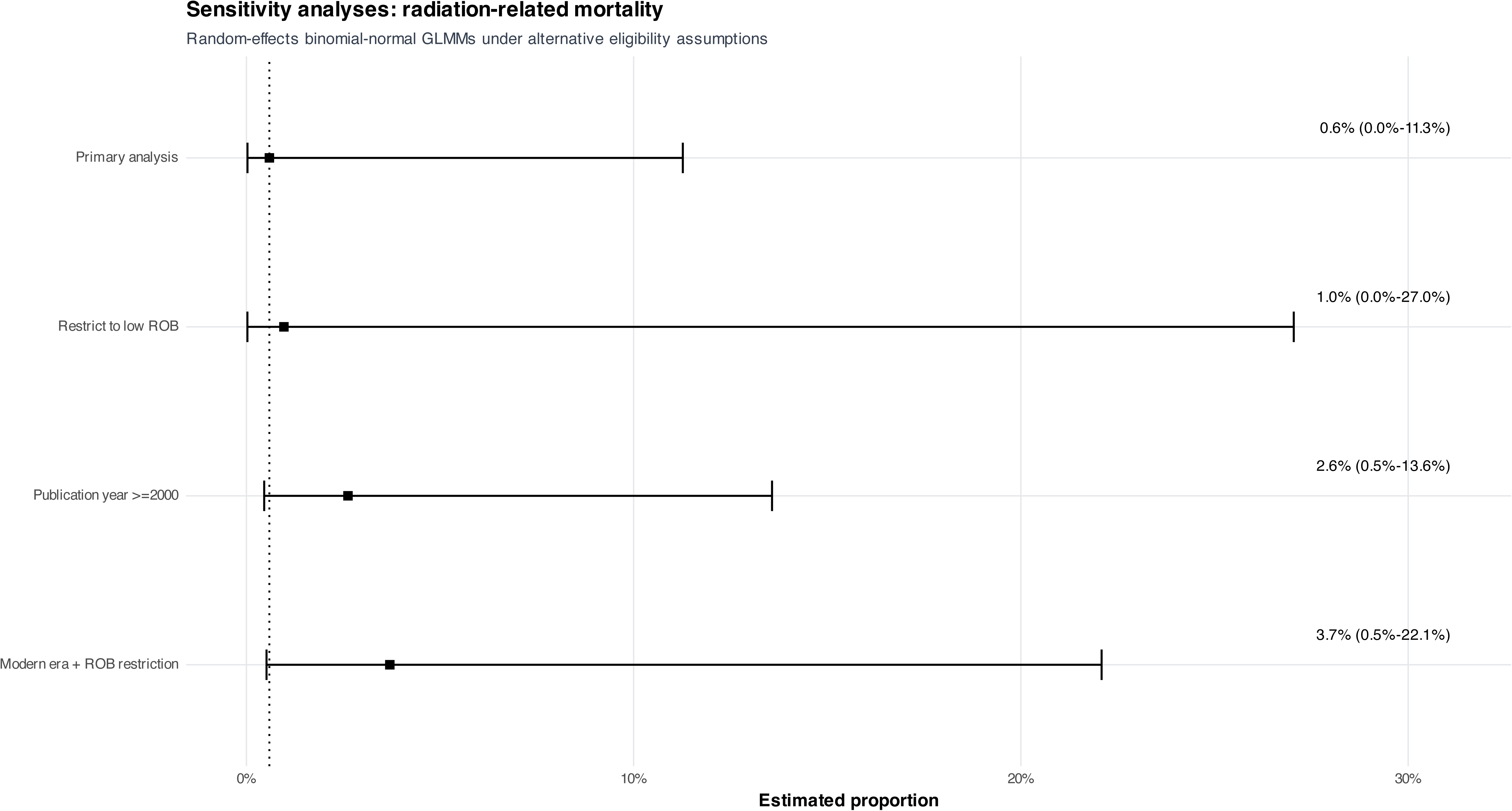
Sensitivity analyses for radiation-related mortality. Comparison of the primary GLMM with low-risk-of-bias, publication-from-2000, and combined modern-era/risk-of-bias restrictions. Estimates remained low but imprecise under all assumptions.

**Supplementary Figure S22.**
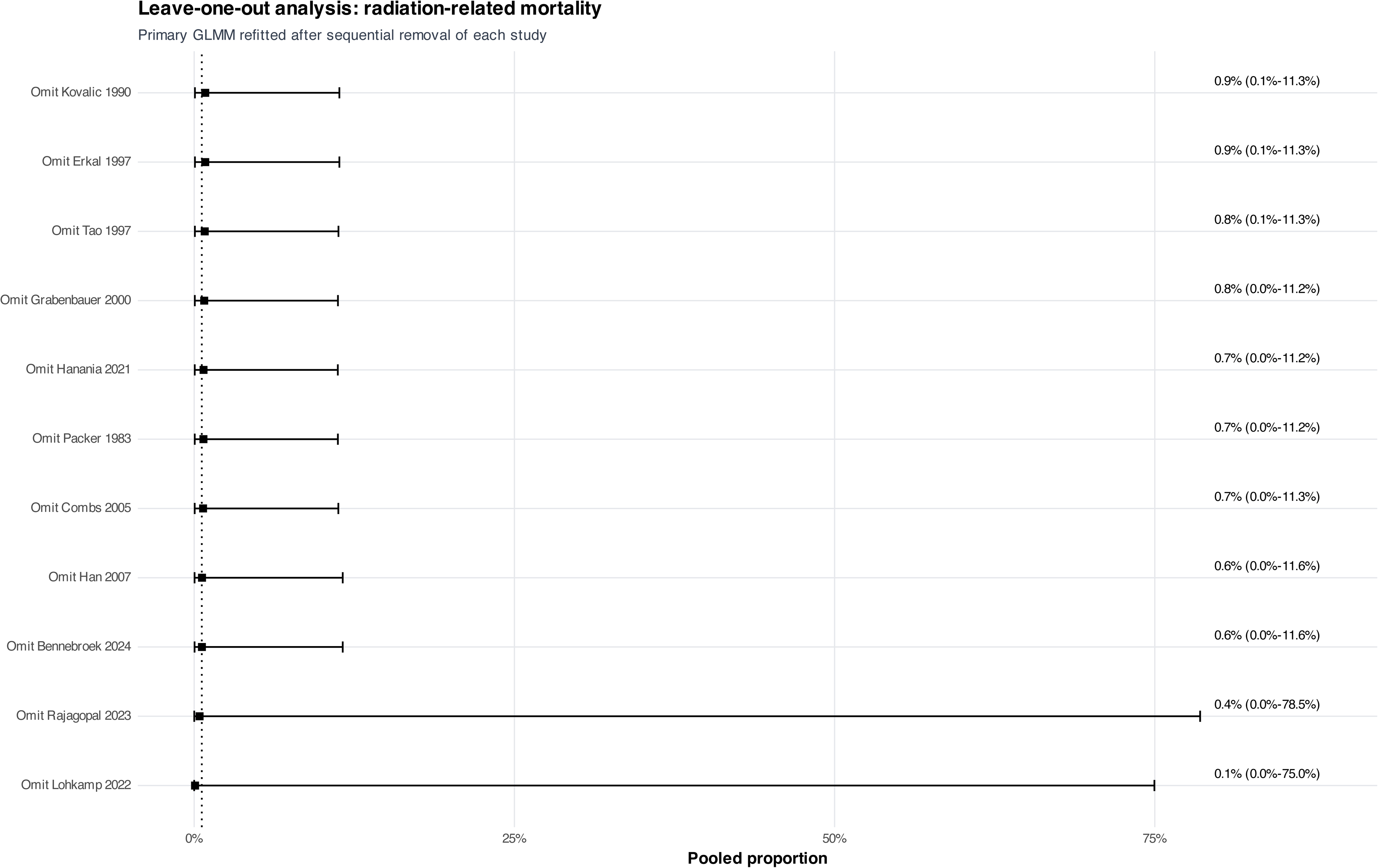
Leave-one-out analysis of radiation-related mortality. Sequential study omission yielded pooled estimates from 0.07% to 0.86%. The endpoint contained three explicitly attributed deaths across 11 studies.

**Supplementary Figure S23.**
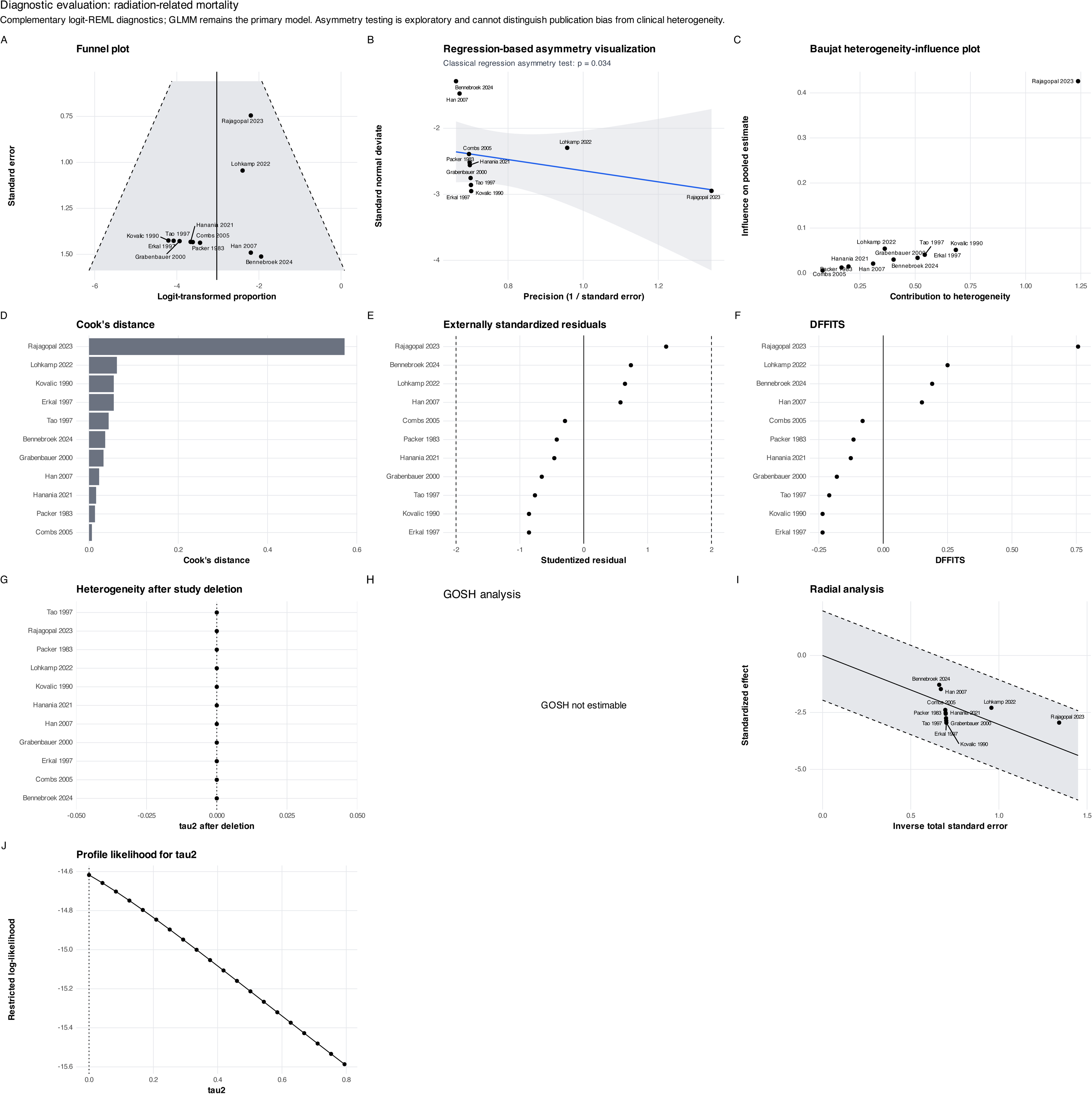
Diagnostic evaluation of the radiation-related-mortality meta-analysis. Funnel and regression-based asymmetry displays, Baujat plot, influence statistics, heterogeneity after deletion, radial analysis, and profile likelihood for τ^2^. Regression-based asymmetry was detected (p=0.034), but this sparse, zero-event-dominated pattern should not be interpreted as proof of publication bias; GOSH analysis was not estimable.

**Supplementary Figure S24.**
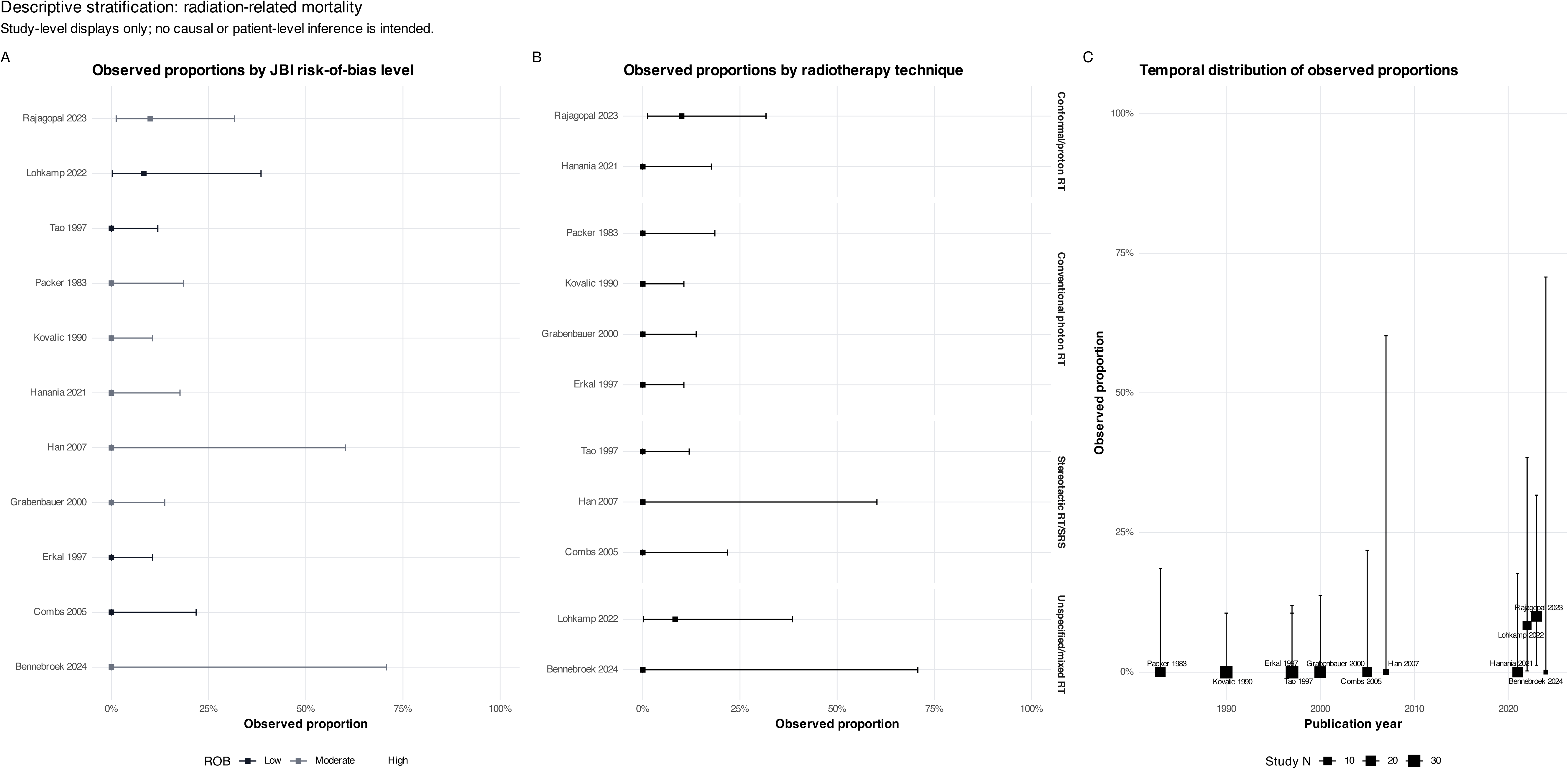
Descriptive stratification of radiation-related mortality. Observed study-level proportions displayed according to JBI risk-of-bias level, radiotherapy technique, and publication year. These plots are descriptive and do not support causal subgroup inference.

**Supplementary Figure S25.**
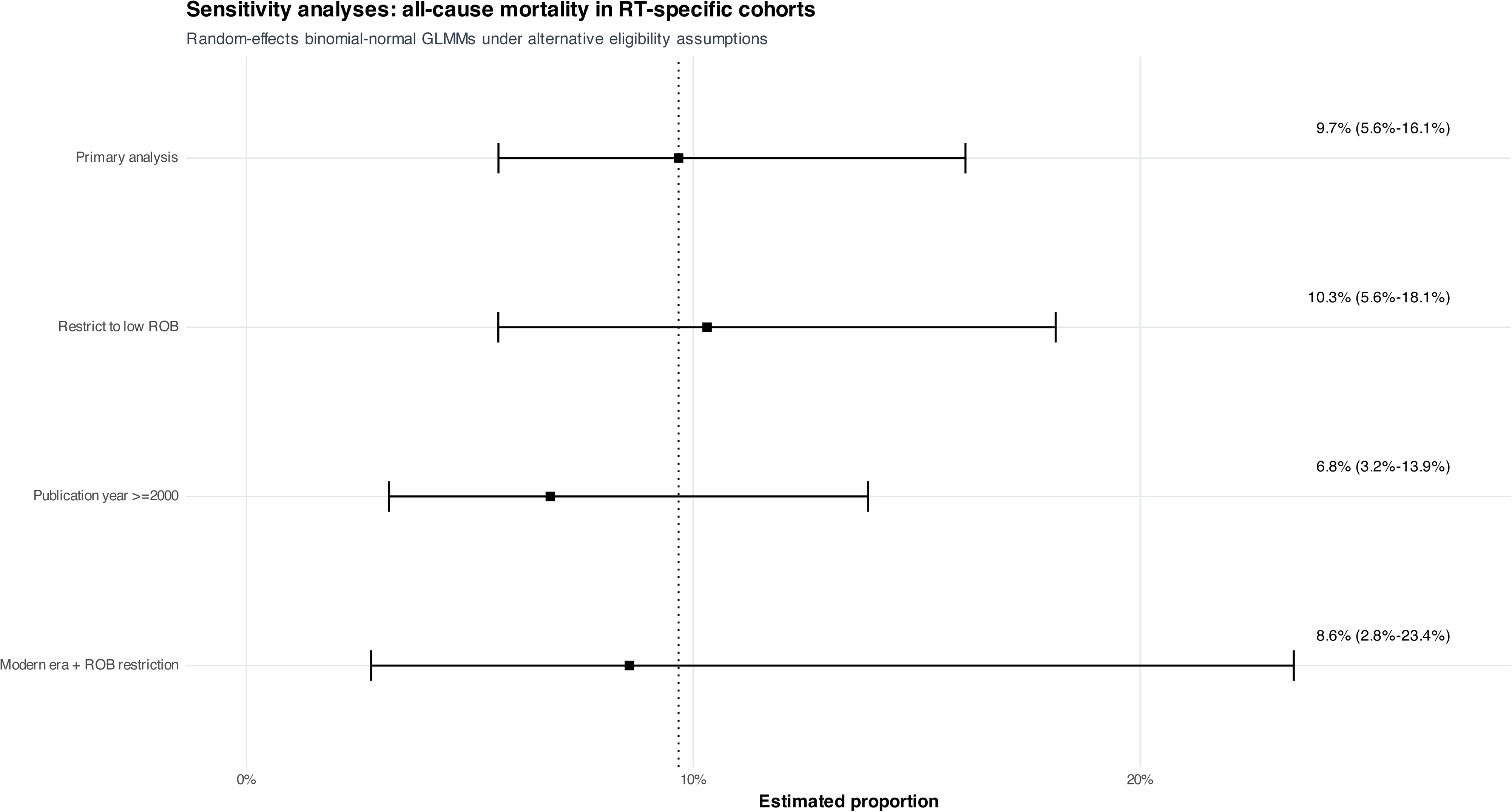
Sensitivity analyses for all-cause mortality in RT-specific cohorts. Comparison of the primary contextual GLMM with low-risk-of-bias, publication-from-2000, and combined modern-era/risk-of-bias restrictions. Pooled estimates ranged from 6.8% to 10.3% across these assumptions.

**Supplementary Figure S26.**
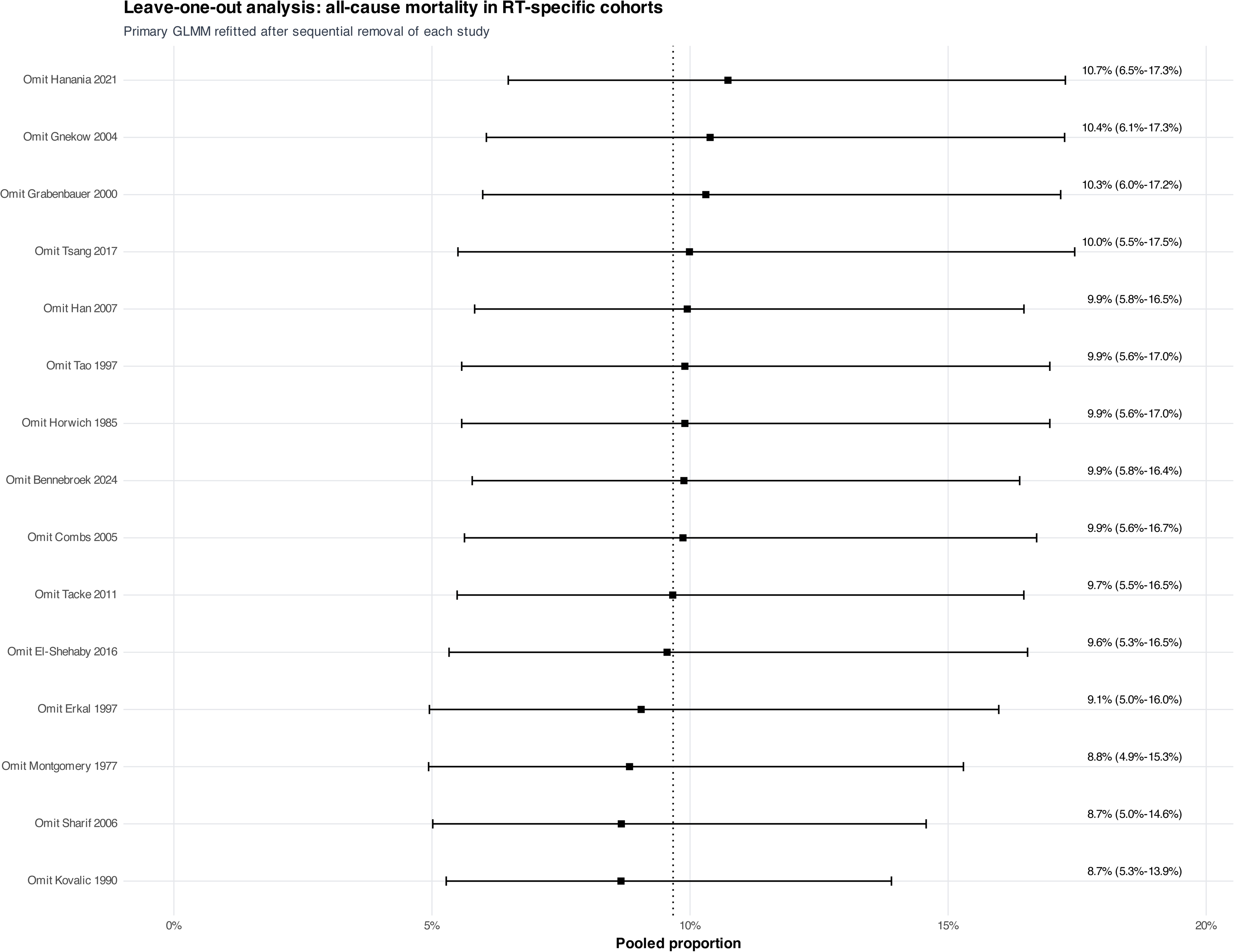
Leave-one-out analysis of all-cause mortality in RT-specific cohorts. Sequential study omission yielded pooled estimates from 8.7% to 10.7%, indicating comparatively limited dependence on any single cohort.

**Supplementary Figure S27.**
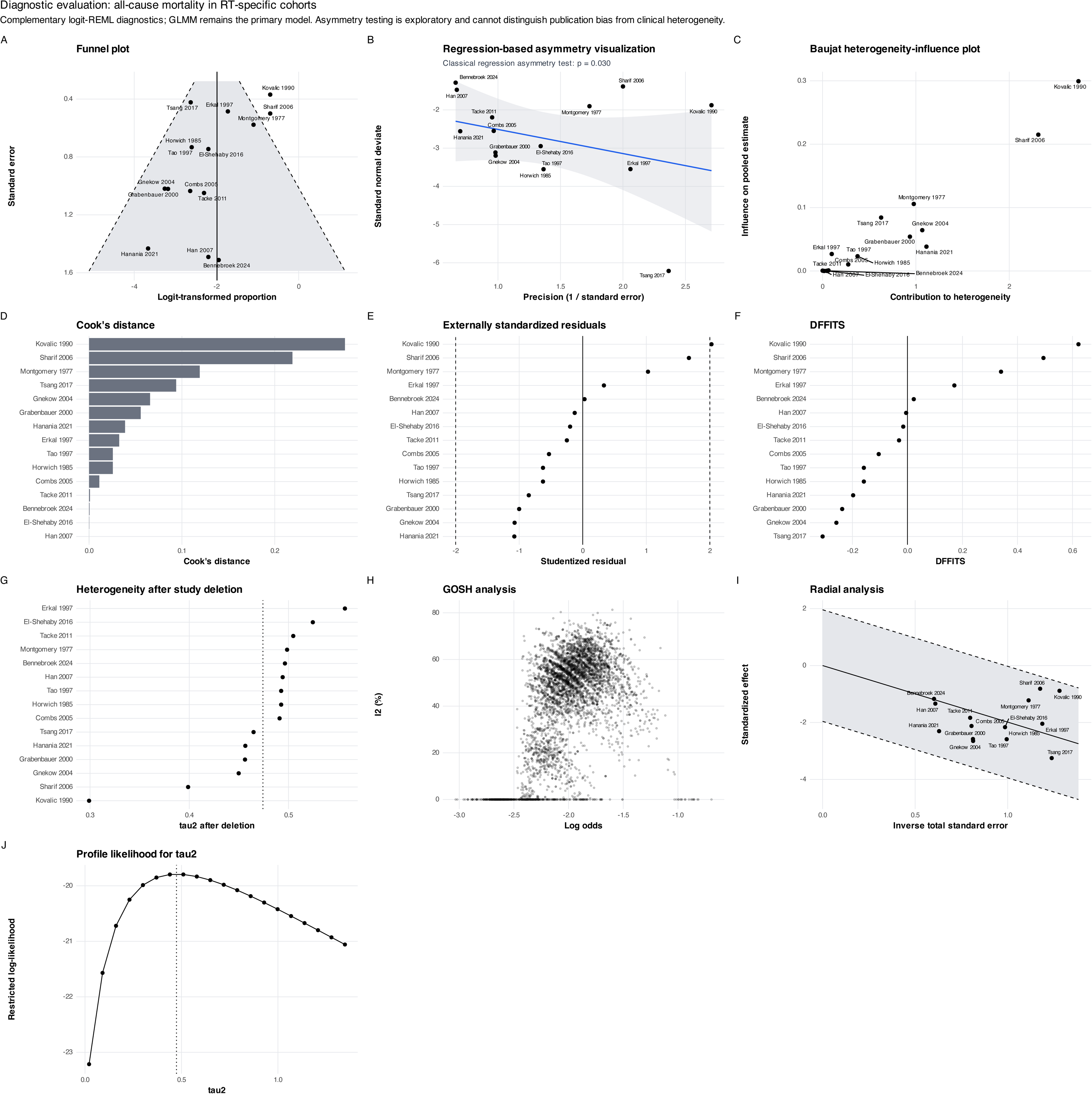
Diagnostic evaluation of all-cause mortality in RT-specific cohorts. Funnel and regression-based asymmetry displays, Baujat plot, influence statistics, heterogeneity after deletion, GOSH analysis, radial analysis, and profile likelihood for τ^2^. Regression-based asymmetry was detected (p=0.030), but may reflect differences in era, follow-up, and case mix rather than selective reporting alone.

**Supplementary Figure S28.**
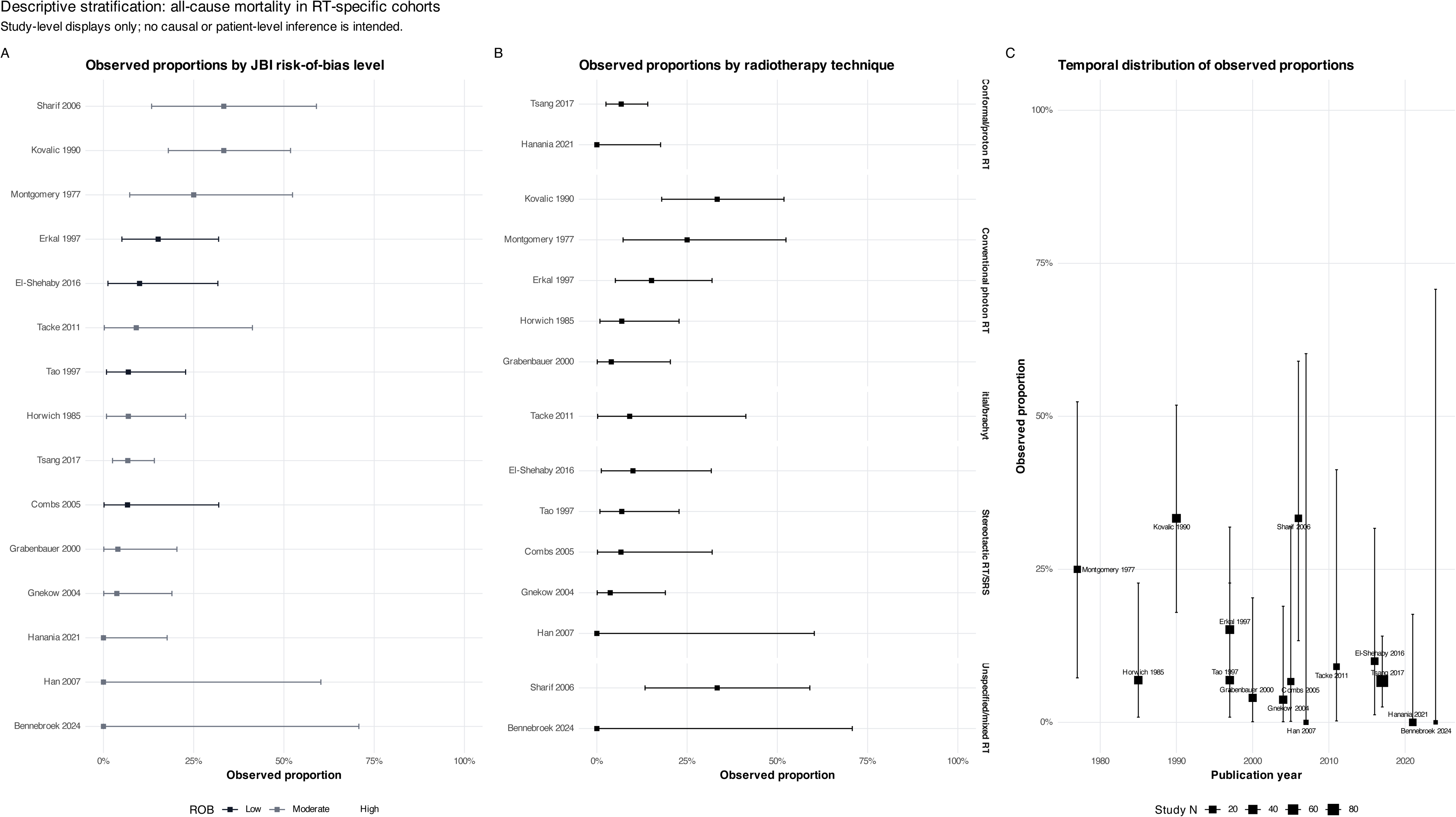
Descriptive stratification of all-cause mortality in RT-specific cohorts. Observed study-level proportions displayed according to JBI risk-of-bias level, radiotherapy technique, and publication year. All-cause mortality remains a contextual endpoint and should not be interpreted as treatment-attributable mortality.

## Supplementary Files

**Supplementary File 1. Completed PRISMA 2020 checklist.**

Completed Preferred Reporting Items for Systematic Reviews and Meta-Analyses 2020 checklist indicating where each applicable reporting item is addressed within the manuscript.

**Supplementary File 2. Complete search strategies.**

Full search strategies for PubMed, Scopus, Web of Science Core Collection, Embase, the Cochrane Library/CENTRAL, Google Scholar, and ClinicalTrials.gov.

**Supplementary File 3. Title and abstract screening workbook.**

Standardized workbook used during independent title and abstract screening, containing bibliographic identifiers, eligibility decisions, and prespecified reasons for exclusion.

**Supplementary File 4. PICOST eligibility and full-text inclusion workbook.**

Standardized eligibility framework documenting assessment according to population, intervention, comparator, outcomes, study design, and time frame, together with the study characteristics supporting final inclusion.

**Supplementary File 5. Full-text exclusion workbook.**

Complete record of reports excluded following full-text assessment, including bibliographic information and the specific prespecified reason for exclusion.

**Supplementary File 6. Complete study-level data extraction dataset.**

Standardized extraction workbook containing methodological characteristics, patient demographics, NF1 status, tumor characteristics, baseline morbidity, previous treatments, radiation modality and parameters, follow-up, toxicity ascertainment and attribution, outcome-specific event counts and denominators, comparative subgroup information, and meta-analysis-ready variables for all included studies.

**Supplementary File 7. Complete Joanna Briggs Institute risk-of-bias assessment.**

Item-level JBI critical appraisal assessments for all included cohort studies, case series, and quasi-experimental studies, together with the prespecified operational study-level risk-of-bias classifications used for graphical presentation.

## Declarations

## Acknowledgements

The authors have no additional acknowledgements to report.

## Preprint disclosure

No preprint version of this manuscript has been posted.

## Ethics approval

Not applicable. This systematic review and meta-analysis was based exclusively on aggregate data obtained from previously published studies. No new human participants were recruited, no intervention was performed, no identifiable personal information was accessed, and no new individual-level participant data or biological material were collected. Therefore, approval from an institutional review board or research ethics committee was not required.

The review was prospectively registered in PROSPERO under registration number **CRD420261474311** and was conducted and reported in accordance with the PRISMA 2020 statement.

## Clinical trial number

Not applicable.

## Consent to participate

Not applicable. No human participants were recruited, contacted, or enrolled, and no new individual-level participant data were collected for this systematic review and meta-analysis.

## Consent for publication

Not applicable. This manuscript does not contain identifiable personal information, individual participant data, or identifiable clinical images.

## Data availability

All data supporting the findings of this systematic review and meta-analysis are included in the article and its supplementary materials. The completed PRISMA 2020 checklist is provided in **Supplementary File 1**, and the complete database search strategies are provided in **Supplementary File 2**. Title and abstract screening decisions are provided in **Supplementary File 3**, while the PICOST eligibility and full-text inclusion workbook is provided in **Supplementary File 4**. Full-text exclusion decisions and corresponding reasons are provided in **Supplementary File 5**.

The complete study-level data-extraction dataset is provided in Supplementary File 6, and the complete Joanna Briggs Institute risk-of-bias assessments are provided in Supplementary File 7. Supplementary sensitivity, influence, subgroup, meta-regression, and diagnostic analyses are provided in the accompanying Supplementary Figures S1-S28.

## Author contributions

**Farzan Fahim (FF)** conceived the study and contributed to its design and methodology; developed the systematic-review framework and standardized screening, eligibility, data-extraction, and risk-of-bias workbooks; trained the reviewers involved in data extraction and methodological appraisal; supervised the systematic-review workflow, evidence synthesis, and manuscript development; contributed to interpretation of the clinical and methodological findings; and critically reviewed and revised the manuscript.

**Amirmahdi Mojtahedzadeh (AMM)** contributed to study conception, methodological development, and coordination of the systematic review and meta-analysis; adjudicated disagreements arising during title and abstract screening, full-text eligibility assessment, data extraction, and risk-of-bias assessment; contributed to quantitative synthesis, interpretation of the clinical and statistical findings, and integration of the safety evidence; supervised manuscript development; and critically reviewed and revised the manuscript for important intellectual content.

**Narin Biabangard (NB)** and **Hanane Sadat Hashemi (HSH)** independently performed title and abstract screening of the deduplicated search records and contributed to verification of study-selection decisions and review of the manuscript.

**Ava Khalili Dehkordi (AKD)** and **Borhan Rahimirad (BR)** independently performed full-text eligibility assessment according to the prespecified PICOST framework, documented inclusion and exclusion decisions, contributed to verification of the final study population, and participated in manuscript review and revision.

**Mobina Puraminaie (MP)** and **Omolbanin Soleymani Pour (OSP)** independently performed duplicate data extraction from the included studies using the standardized extraction framework, contributed to reconciliation and verification of the extracted dataset, and participated in interpretation and revision of the manuscript.

**Farnaz Mortezazade (FM)** and **Parvane Tayebzadeh (PT)** independently performed the design-specific Joanna Briggs Institute risk-of-bias assessments, contributed to methodological appraisal and verification of the included evidence, and participated in manuscript review and revision.

**Mahan Kamali (MK), Maryam Yaftian (MY), Kowsar Hariri (KH), Niloofar Sadeghi (NS), and Fatemeh Khazaei (FK)** contributed to review and interpretation of the relevant literature, synthesis of clinical and safety findings, manuscript drafting, and critical revision of the manuscript for intellectual content.

**Alireza Zali (AZ)** provided senior neurosurgical and academic supervision, contributed to interpretation of the clinical implications of radiotherapy and radiosurgery for optic pathway–hypothalamic glioma, guided refinement of the manuscript’s clinical perspective, and critically reviewed and revised the manuscript for important intellectual content.

**Farzan Fahim and Amirmahdi Mojtahedzadeh contributed equally to this work and share first authorship. Farzan Fahim and Amirmahdi Mojtahedzadeh share corresponding authorship.**

All authors reviewed and approved the final version of the manuscript and agree to be accountable for the work.

## Declaration of generative AI and AI-assisted technologies in the manuscript preparation process

During the preparation of this work, the authors used **ChatGPT by OpenAI** to assist with **grammar, and language refinement,**. The tool was not used to generate original research data, perform data extraction, conduct statistical analyses, create or alter figures, or replace the authors’ scientific judgment. After using this tool, the authors reviewed and edited the content as needed and take full responsibility for the content of the submitted manuscript.

